# Type 2 diabetes genetics in 125,000 admixed adults from Mexico City

**DOI:** 10.64898/2026.06.23.26356360

**Authors:** Jason M. Torres, Jaime Berumen, Diego Aguilar-Ramirez, Eirini Trichia, Francisco Barajas-Olmos, Humberto García-Ortiz, Sam Morris, Michael Turner, Elizabeth Barrera, Tianshu Liu, Consuelo D. Quinto-Cortés, Lauren E. Petty, Lourdes García-García, María Teresa Tusié-Luna, Carlos A. Aguilar-Salinas, Amanda Y. Chong, Paulina Baca, Fernando Rivas, Fiona Bragg, Louisa Gnatiuc Friedrichs, Michael R. Hill, Lisa Holland, Alejandra Vergara-Lope, Rachel Wade, Michael J. Gaziano, Rory Collins, Eleanor Wheeler, Alan R. Shuldiner, Jennifer E. Below, Kari E. North, Alexandre C. Pereira, Andrés Moreno-Estrada, Lorena Orozco, Jesús Alegre-Díaz, Pablo Kuri-Morales, Jonathan R. Emberson, Roberto Tapia-Conyer

## Abstract

Type 2 diabetes (T2D) is a highly heritable, polygenic disease with over 600 loci identified through genome-wide association studies (GWAS). However, despite possessing unique genetic variation shaped by demographic history and admixture, Latin American populations remain markedly underrepresented in global genomic research. To address this gap, we conducted genome- and exome-wide analyses of 19,431 T2D cases and 105,611 controls from the Mexico City Prospective Study (MCPS). We identified 86 independent GWAS associations, including 21 novel signals, 15 of which replicated in external cohorts. Risk alleles at novel loci were enriched in individuals with Indigenous American ancestry. Exome analyses revealed rare and ultra-rare missense variants with substantial risk effects at *HNF1A* and *GCK*, as well as a protein-damaging variant in *SLC30A8* that reduced T2D risk by 45% in carriers. Integrative analyses indicate that T2D genetic architecture in Mexico is predominantly driven by common regulatory variation acting in the endocrine pancreas. Polygenic risk scores strongly stratified T2D risk and transferred to Indigenous Mexican populations. These findings demonstrate the power of large-scale genetic discovery in diverse populations to refine disease architecture and identify loci with potential therapeutic relevance.

## Introduction

Genome-wide association studies (GWAS) have identified over 600 loci associated with type 2 diabetes (T2D)^1–3^, providing significant molecular insight into its etiology, but most such studies have been conducted in populations of European ancestry^4^. By contrast, individuals of Latin American descent remain underrepresented in genetic studies of T2D, thereby impeding the discovery of novel genetic risk variants that may segregate at higher frequencies in Latin America. Indeed, previous sequencing studies in Mexico—where the prevalence of adiposity and diabetes is nearly twice that observed in European populations—have identified novel T2D risk variants at the *SLC16A11*, *HNF1A*, and *IGF1* loci with moderate to large effects despite involving only a few thousand participants^5–7^. Moreover, genetic variation in Latin America, shaped by pre-Hispanic demographic events, admixture between Indigenous American and non-American ancestries, and post-colonial population growth, provides unique opportunities for resolving the genetic basis of T2D^8,9^. The comparative lack of non-European participants in GWAS also undermines the transferability of polygenic risk scores (PRS) across global populations, exacerbating existing health disparities^4^. In this study, we leverage the high prevalence of T2D in Mexico and the expansive genetic datasets (i.e. TOPMed-imputed genotypes, exome and whole-genome sequencing) from the Mexico City Prospective Study (MCPS)^10^—the largest blood-based prospective study in Latin America—to identify, replicate and fine-map novel loci for T2D. We further perform phenome-wide association analyses and integrate regulatory genomic maps to resolve biological processes mediating genetic effects on disease risk. Lastly, we evaluate polygenicity, selection and transferability of PRS in admixed and Indigenous American populations in Mexico. An overview of the study design is presented in Fig. 1a.

**Fig. 1.**
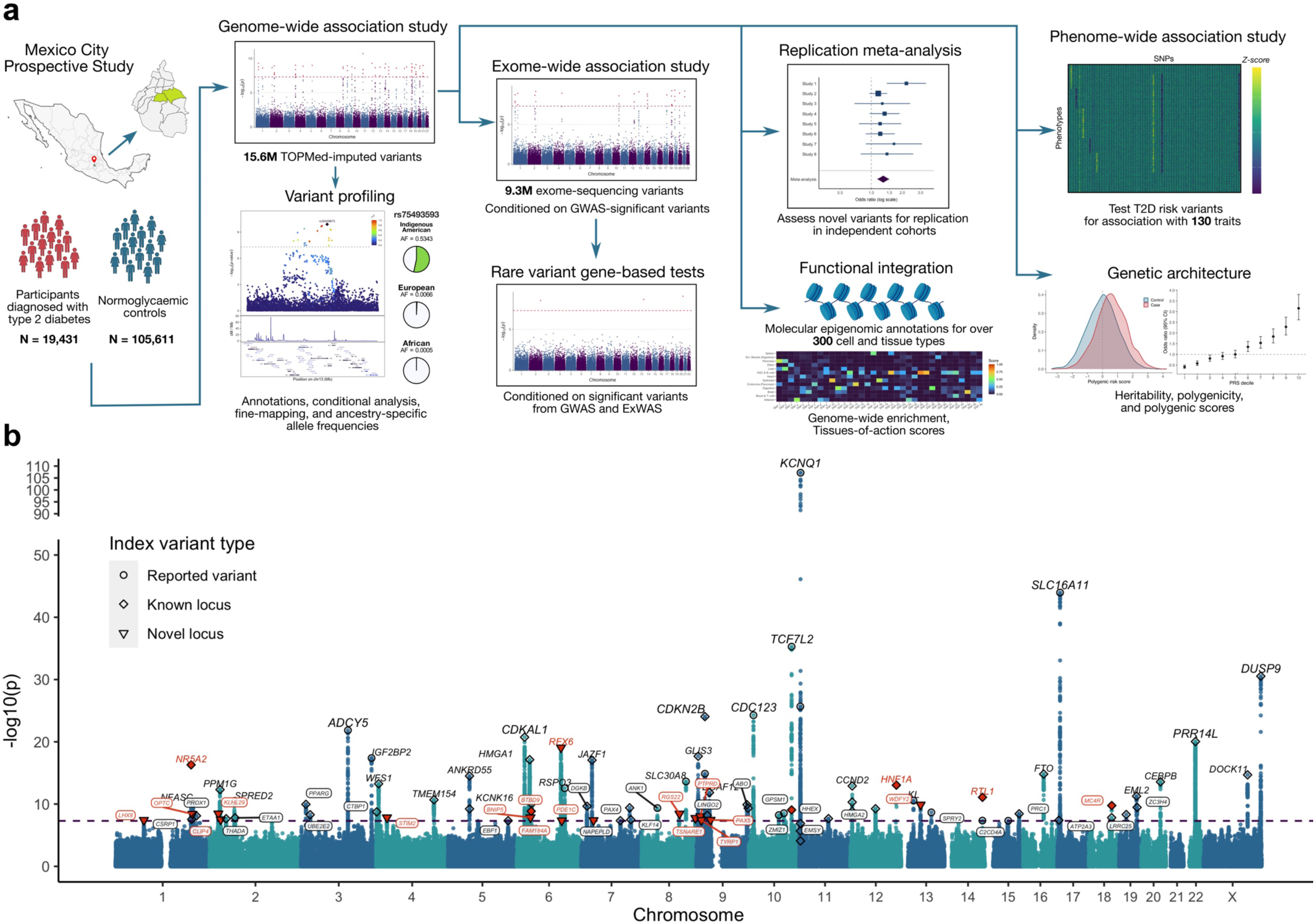
Genetic analysis of type 2 diabetes in the Mexico City Prospective Study. **a**, Overview of the genetic discovery analyses performed in this study. **b**, Manhattan plot from the genome-wide association study of T2D. The y-axis shows –log_10_(*p)* values from association statistics estimated using SUGEN. Classification of index variants are indicated by shape, and novel associations are highlighted in red. The nearest protein-coding gene to each index variant, based on Ensembl v105, is labelled.

## Results

### Genome-wide association study

We identified 19,431 individuals with previously-diagnosed T2D and 105,611 non-diabetic controls (see Methods) of admixed American ancestry in the MCPS cohort using data on diabetes status at recruitment, age at diagnosis, HbA1c levels, and use of glucose-lowering medication. The resulting effective sample (*N_eff_* = 65,646) is 2.4 times larger than that of the Hispanic/Latino (H/L) DIAMANTE meta-analysis^2^. After quality control and imputation to the TOPMed^11^ reference panel (r2), 15,578,291 autosomal and chromosome X variants were retained for GWAS.

To account for extensive relatedness in MCPS, where 71% of participants have at least one close relative within the third degree^10^, we identified 23,213 first-degree family networks and applied the generalised estimating equation-based method implemented in SUGEN (Methods). We identified 5,535 genome-wide significant variants (*p* < 5×10^-8^) and observed concordant effects at known T2D risk loci (Extended Data Fig. 1). GWAS results remained largely consistent after further adjustment for adiposity traits (see Supplementary Information).

The observed genomic control inflation factor (*λ*_!"_) was 1.25 (Extended Data Fig. 2a–b). Using covariate-adjusted linkage disequilibrium (LD) scores derived from whole-genome sequence data in 9,764 unrelated MCPS participants^12^ (see Methods), the LD score regression intercept was 1.064 (SE = 0.0026), indicating that most inflation reflected polygenicity rather than residual confounding.

Conditional analysis identified 86 independent associations (“signals”) at 75 loci (Fig. 1b, Supplementary Fig. 1 and Supplementary Table 1), including 65 previously reported T2D signals and 21 novel signals. Known signals included *SLC16A11* (rs113748381), a T2D locus first identified in Mexicans by the SIGMA consortium^5^. Of the novel signals, six mapped to established T2D loci but were independent of previously reported variants and were therefore considered *novel* associations at *known* T2D loci. These included predicted deleterious missense variants at *HNF1A* (rs483353044) and *MC4R* (rs79783591), together with non-coding variants at *TCF7L2* (rs148113496) and three additional loci (Fig. 2a, Supplementary Table 2). The remaining 15 signals were located more than 500 kb from any previously reported T2D variant in the NHGRI-EBI GWAS catalog and were therefore considered *novel* loci, including predicted deleterious missense variants in *WDFY2* (rs35409673) and *RFX6* (rs139343836) (Fig. 2a). We also observed allelic heterogeneity at seven loci, with *KCNQ1* harbouring five independent signals (Supplementary Table 1, Supplementary Fig. 1).

**Fig. 2.**
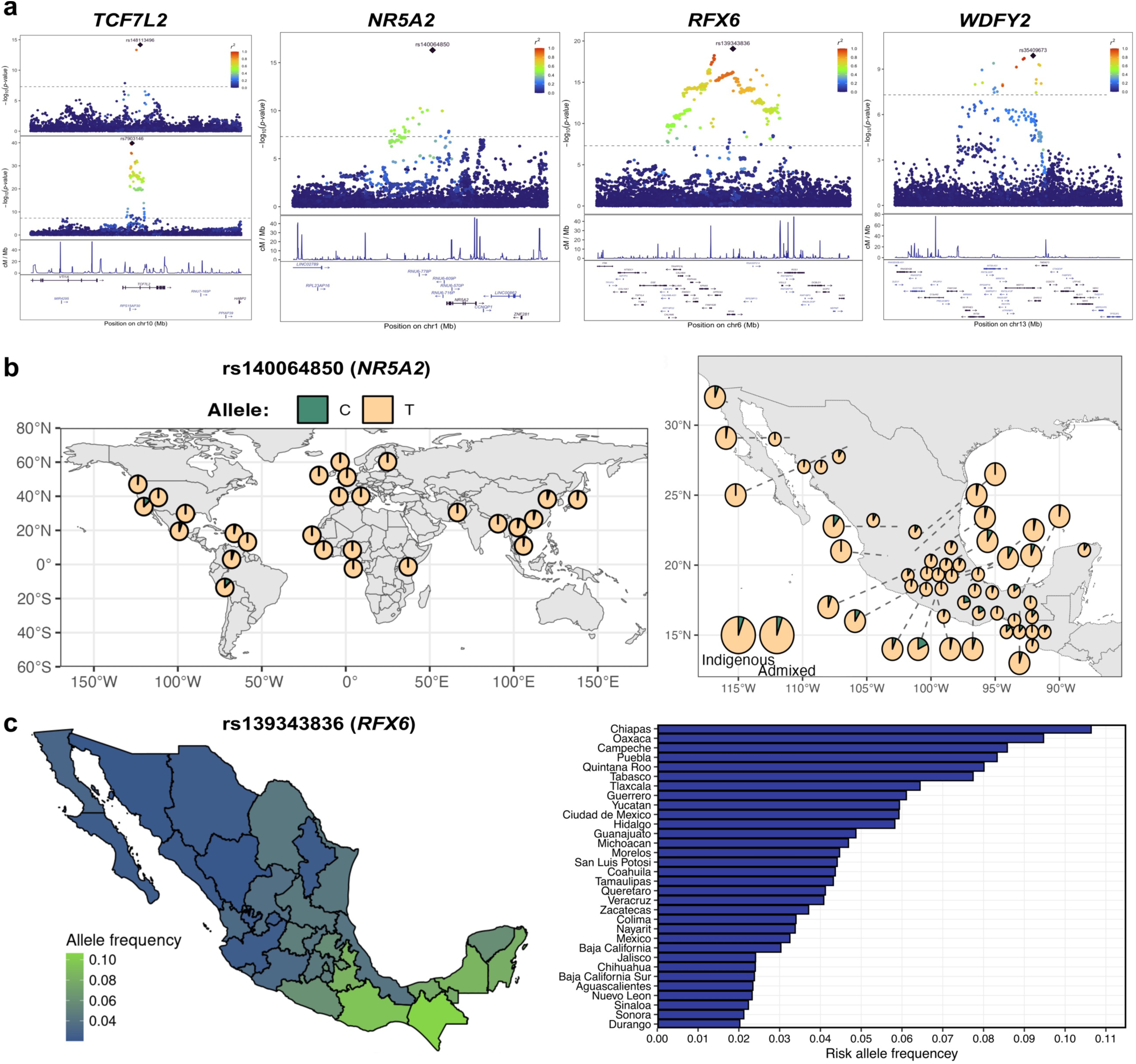
Novel T2D associations identified in MCPS. **a**, Locus plots for novel signals at *TCF7L2*, *NR5A2*, *RFX6* and *WDFY2*. The y-axis shows −log_10_(*p)* values from approximate conditional analyses. Recombination rate and GENCODE gene annotations are shown below, with protein-coding and non-coding genes displayed in dark purple and blue, respectively. The novel secondary signal at *TCF7L2* (rs148113496) is shown with the established primary signal (rs7903146). **b**, Frequency of the rs140064850 risk allele (C) at *NR5A2* across 1000 Genomes populations (left) and admixed and Indigenous Mexican populations from the MAIS study (right). **c**, Frequency of the rs139343836 risk allele (A) at *RFX6* across 31 Mexican states. Allele frequencies were estimated from the Mexico Biobank and visualized using the MexVar platform.

Lower frequency variants tended to have higher odds ratios (ORs). Among the four rare signals (MAF < 1%), ORs ranged from 1.45 to 2.48, whereas common variants (MAF > 5%) had a median OR of 1.10 (IQR 1.08–1.13) (Extended Data Fig. 2c). Several common variants nevertheless exhibited relatively large effects, including *SLC16A11* (rs113748381; MAF = 35.3%, OR = 1.19, 95% CI 1.17–1.22). The novel *RFX6* association (rs139343836; risk allele frequency [RAF] = 5.1%) also involved a common risk allele with OR = 1.26 (95% CI 1.19–1.33). At 43 of the 79 common signals, the major allele conferred increased risk in MCPS (Extended Data Fig. 2c, Supplementary Table 1). Genome-wide significant common variants explained 8.1% of T2D variance, compared with 0.48% for low-frequency and rare variants (see Methods), consistent with a genetic architecture largely driven by common variation^13^.

### Fine-mapping

Fine-mapping identified 23 signals at 18 loci with maximum posterior probability of association (PPA) > 0.5, indicating that a single variant accounted for most of the association signal (Supplementary Table 3). Despite a 3.5-fold larger effective sample size in the European DIAGRAM meta-analysis^14^ (*N_eff_* = 231,436), fine-mapping resolution was improved in MCPS for 11 shared signals (Extended Data Fig. 3, Supplementary Table 4). The greatest improvement was observed at the *KCNQ1* locus, where the PPA for rs4929965 increased from 0.20 in DIAGRAM to 0.72 in MCPS. Two resolved to a single variant (i.e. PPA ≥ 0.99), including the novel association at *NR5A2* (rs140064850).

### Exome variant and gene burden tests

Exome sequencing in over 141,000 MCPS participants enabled a comprehensive analysis of rare and ultra-rare coding variants. An exome-wide association study (ExWAS) identified 290 variants associated with T2D at *p* < 5×10^-8^, almost all of which (97.9%) mapped to loci implicated in the GWAS (Extended Data Fig. 4a, Supplementary Table 5).

Missense index variants from the GWAS were also the strongest ExWAS associations at their respective loci, including the novel signals in *RFX6*, *WDFY2*, *HNF1A* and *MC4R*, with each predicted to be deleterious by one or more pathogenicity tools (Supplementary Table 5).

As most of the risk alleles were common (99.0% of ExWAS-significant variants had RAF > 1%), we conditioned on significant GWAS variants and found that only 11 exome variants remained significant (Extended Data Fig. 4b, Supplementary Table 6). The strongest associations involved two rare missense variants (rs754729248 and rs371717826) in *HNF1A*, a gene in which protein-damaging variants can cause maturity onset diabetes of the young (MODY)^15^. Both variants were rare in MCPS (RAFs = 0.0018 and 0.0007, respectively) and virtually absent outside of Mexico (e.g., RAFs = 3.2×10^-4^ and 3.0×10^-6^ in RGC Million Exome resource [URL]). Their effects were large (OR = 2.56 95% CI 2.05–3.19 and OR = 2.86, 95% CI 2.04–4.01, respectively) and both were predicted to be highly damaging (Supplementary Table 6). Notably, the variants altered the same proline residue at position 379, substituting alanine and histidine, respectively. These findings provide genetic evidence supporting a role for both variants in T2D susceptibility, despite previous conflicting clinical interpretations^16^.

Another notable ExWAS association involved a missense variant (rs145677283) in the established diabetes gene *SLC30A8*, which encodes a zinc transporter involved in insulin secretion. This variant was extremely rare globally (e.g., MAF = 3.0×10^-6^ in the RGC 1 Million Exomes study), but was enriched on Indigenous American (IAM) local ancestry segments in MCPS (MAF = 0.005). The minor “A” allele conferred a protective effect (OR = 0.52, 95% CI 0.41–0.66), corresponding to an estimated ∼45% reduction in T2D risk among carriers, and its corresponding arginine-to-histidine substitution at position 165 was predicted to be damaging by multiple pathogenicity metrics (Supplementary Table 6). Although this variant has not previously been implicated in diabetes, other rare protein-damaging variants in *SLC30A8* are known to protect against T2D^17,18^.

Gene-based testing of ultra-rare predicted loss-of-function and damaging missense variants across 18,156 protein-coding genes identified a single association after conditioning on GWAS and ExWAS signals: *GCK* (*p* = 2.2×10^-10^; Extended Data Fig. 4c). This gene encodes glucokinase—an enzyme that acts as a glucose sensor within pancreatic ß-cells—and loss-of-function mutations in *GCK* are known to cause MODY type 2^15^. Although the signal reflected the aggregate effect of 35 qualifying variants, it was largely driven by rs1131691598 (p.Leu45Pro), which strongly associated with T2D at Bonferroni significance (0.05 / 35) (OR = 52.4, 95% CI 6.4–427.0, *p* = 2.2×10^-4^). Carriers of the risk allele had significantly higher HbA1c levels than non-carriers (Fig. 3; Supplementary Fig. 2). The variant was extremely rare (RAF = 1.01×10^-4^) and was observed exclusively on IAM local ancestry segments.

**Fig. 3.**
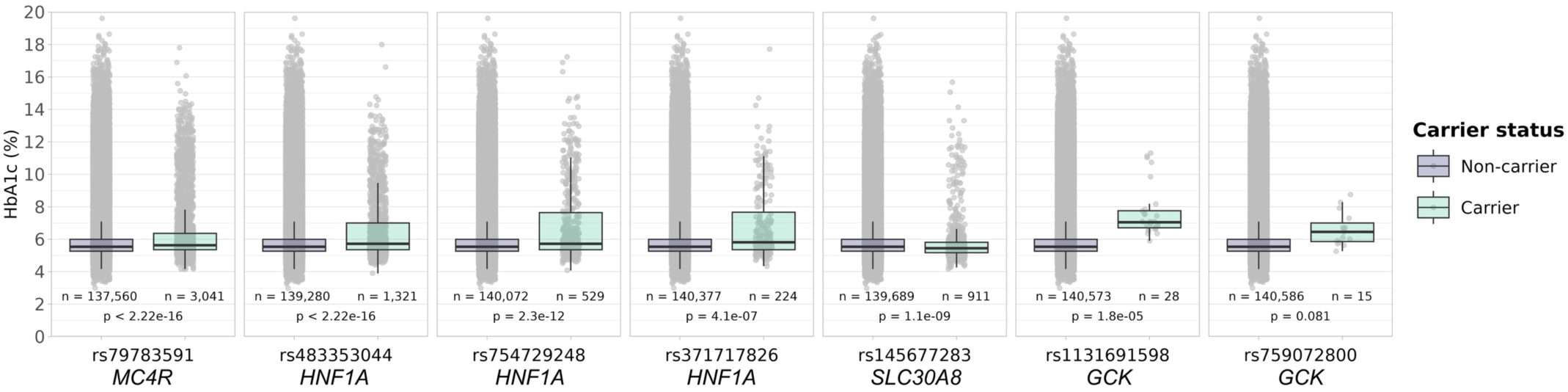
Effects of rare protein-damaging variants on HbA1c levels in MCPS. Box-and-whisker plots show the distribution of HbA1c (%) among carriers and non-carriers of predicted damaging missense variants. Variants include GWAS signals (*MC4R* rs79783591 and *HNF1A* rs483353044), ExWAS signals (*HNF1A* rs754729248 and rs371717826; *SLC30A8* rs145677283), and rare *GCK* variants included in the gene-level analyses (rs1131691598 and rs759072800). The variant rs1131691598 (p.Leu45Pro) represents a novel *GCK* association in MCPS, whereas rs759072800 (p.Asp124Asn) is annotated as pathogenic for MODY2 in ClinVar. Grey points indicate individual participant values; *p* values correspond to two-sided *t*-tests comparing mean HbA1c levels between carriers and non-carriers.

### Replication meta-analyses

We assessed replication of novel GWAS signals using summary statistics from external studies (Supplementary Table 7), including Hispanic/Latino (H/L) GWAS meta-analyses from T2DGGI^3^ and DIAMANTE^2^, H/L participants from the Million Veteran Program (MVP)^1^, admixed participants from the Mexico Biobank (MXB)^19^, and Indigenous Mexican participants from the MAIS study^9^.

We performed inverse-variance-weighted meta-analysis across all non-overlapping studies and separately across H/L and non-H/L cohorts (see Methods). Fourteen of the 21 signals (67%) met replication criteria in the combined meta-analysis (Table 1, Supplementary Fig. 3). All but one of the novel signals at a known locus replicated, including the *NR5A2* signal rs140064850, which had an OR of 1.14 (95% CI 1.10–1.18) in MCPS and 1.10 (95% CI 1.06–1.14) in the replication meta-analysis (Extended Data Fig. 5a). Two replicated signals, *HNF1A* (rs483353044) and *MC4R* (rs79783591), were rare in MCPS and absent from all non-Hispanic/Latino studies, but nevertheless replicated in the Hispanic/Latino meta-analysis. Eight of the 15 novel loci also showed evidence of replication (Table 1).

**Table 1.**
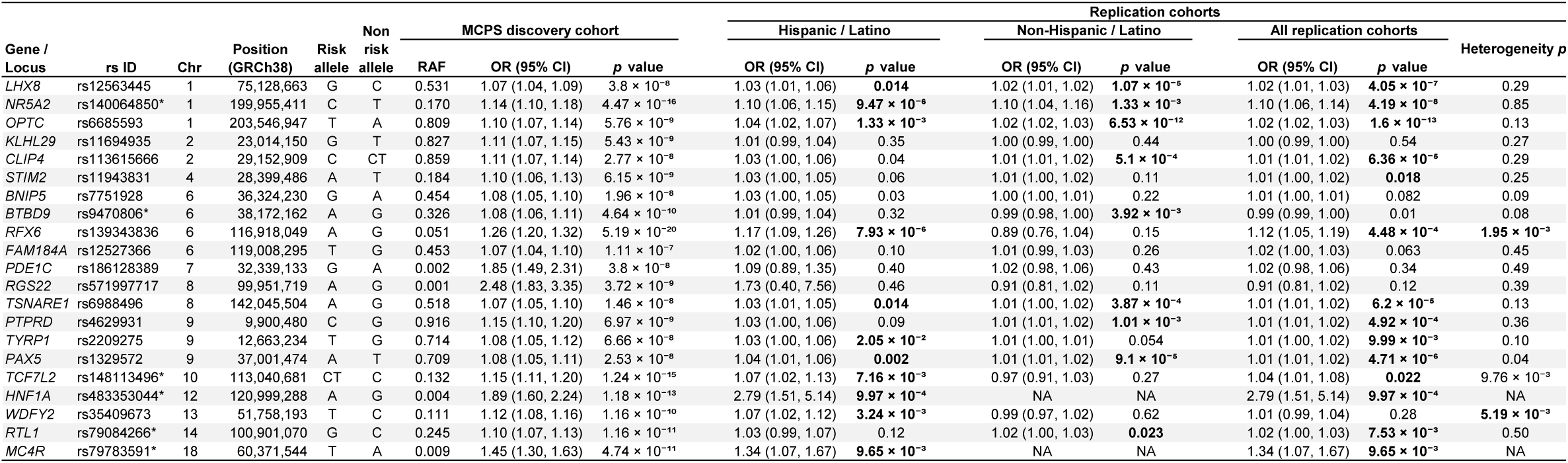
Effects of novel T2D signals in replication cohorts. RAF, risk allele frequency. *p* values are reported to two significant figures or, where *p* < 0.001, in scientific notation with two decimal places. An asterisk (*) denotes novel associations at previously reported T2D loci. Bold text for replication-cohort *p* values indicates associations meeting replication criteria, defined as directional consistency with MCPS and a Benjamini–Hochberg-adjusted *p* < 0.05. Bold text for heterogeneity *p* values indicates a Benjamini–Hochberg-adjusted *p* < 0.05.

We examined allele frequencies from 26 populations in the 1000 Genomes Project, 34 Indigenous, and 20 admixed populations across Mexico, and local ancestry-inferred frequencies in MCPS^10^. Risk alleles for most replicated variants (12 of 15) were enriched on Indigenous American ancestry backgrounds (Supplementary Tables 8–10), including signals at *WDFY2*, *PAX5*, *RTL1*, *TCF7L2*, *HNF1A*, and *MC4R*. Notably, the risk allele (“C”) at rs140064850 (*NR5A2*) had an IAM allele frequency of 26.5%, compared to just 0.95% and 0.56% in African and European ancestries, respectively (Fig. 2b, Supplementary Table 8). Among global populations, this allele was most frequent in the Matlatzinca (RAF = 0.214), an Indigenous community from central Mexico belonging to the Oto-Mangue linguistic family (Fig 2b, Supplementary Tables 9–10). Similarly, the “A” risk allele at rs139343836 (*RFX6*) had a frequency of ∼8% in IAM populations, but was present at frequencies below 0.01% in non-American populations (Supplementary Tables 8–10). Its frequency was highest in the Kaqchiquel (RAF = 0.256), a Maya-speaking Mesoamerican population from southeastern Mexico and the Guatemalan highlands, and was most prevalent in the Mexican states of Chiapas, Campeche and Oaxaca (RAFs = 0.106, 0.095 and 0.086, respectively; Fig. 2c), according to the MexVar platform^20^.

We tested for heterogeneity in effect size estimates between H/L and non-H/L cohorts and observed FDR-significant differences for rs139343836 (*RFX6*; *p_het_* = 1.95×10^-3^) and rs35409673 (*WDFY2*; *p_het_* = 5.19×10^-3^), together with suggestive evidence at rs148113496 (*TCF7L2*; *p_het_* = 9.76×10^-3^) (Table 1). Risk alleles at all three signals were enriched on IAM ancestry backgrounds (Supplementary Tables 8-10). The risk alleles at rs35409673 and rs148113496 were associated with T2D in H/L cohorts (OR = 1.07 for both variants) but not in non-H/L cohorts (Table 1). The difference was most pronounced for rs139343836 (*RFX6*), for which the “A” allele increased T2D risk in H/L cohorts (OR = 1.17, 95% CI 1.09–1.26) but was not associated in non-H/L cohorts (OR = 0.89, 95% CI 0.76–1.04), where it was substantially less frequent (maximum allele frequency 0.071 versus 0.0039; Extended Data Fig. 5b).

We further investigated rs139343836 (*RFX6*) and identified a significant interaction with global Indigenous American ancestry, such that the effect size attenuated with increasing IAM ancestry proportion (OR_interaction_ = 0.63, 95% CI [0.47, 0.85]; *p* = 0.0023; Supplementary Fig. 4). In silico analysis further suggested that the missense substitution encoded by rs139343836 (p.Val329Ile) affects a conserved residue within a functional domain of the protein (Supplementary Fig. 5). Additional analyses of this variant are described in the Supplementary Information.

### Phenome-wide association study

To further characterize GWAS signals, we performed a phenome-wide association study (PheWAS) of 130 traits present in MCPS, including 110 NMR-based metabolite features, among participants without diabetes or other chronic diseases (see Methods). At FDR < 5%, we identified 1,436 genotype-trait associations involving 82 variants (Extended Data Fig. 6, Supplementary Table 11). Over half of the index variants (44 out of the 86 GWAS signals) had risk alleles associated with elevated HbA1c (e.g., signals at *ADCY5* and *SLC30A8*), while 31 variants were associated with ≥10 metabolite traits, including eight associated with more than 50 traits (Extended Data Fig. 6, Supplementary Tables 11–12).

Thirteen variants associated with adiposity measures (BMI, hip and waist circumference), including known signals at *FTO* and *MC4R*, and novel signals at *MC4R* (rs79783591), *TSNARE1* (rs6988496), *PAX5* (rs1329572), and *LHX8* (rs12563445). The risk allele at the novel *MC4R* missense variant was also associated with increased waist-to-hip ratio (*β*=0.131, 95% CI 0.084–0.178, *p* = 2.8×10^-8^) (Extended Data Fig. 7a, Supplementary Table 11).

To contextualize biological mechanisms, we integrated MCPS PheWAS results with external trait-association data using a “physiological clustering” framework^21^ (see Supplementary Information). Replicated signals were assigned to clusters reflecting impaired insulin secretion, insulin action or lipid metabolism (Extended Data Fig. 7b, Supplementary Tables 13–14). *HNF1A* (rs483353044) clustered with impaired insulin secretion signals, whereas *SLC16A11* (rs113748381) clustered with signals associated with impaired lipid metabolism. Most remaining replicated signals, including *NR5A2*, *WDFY2*, *TCF7L2*, *RTL1* and *RFX6*, mapped to a cluster with mixed effects on insulin secretion and action. Notably, despite its adiposity associations, the rare *MC4R* variant rs79783591 did not cluster with established BMI-associated variants at *FTO* or *MC4R*, reflecting its distinct metabolomic profile characterised by negative associations with ApoA1, omega-6 fatty acids, and HDL cholesterol, and no positive associations with triglycerides.

### Functional integration

To further investigate biological processes influencing T2D risk in this population, we integrated GWAS results with functional genomic data. Enrichment analysis across 1,202 chromatin accessibility annotations from the Common Metabolic Diseases Genome Atlas, spanning 192 tissues and cell types, identified 293 enriched annotations from 34 tissues or cell types at FDR < 5% (Supplementary Table 15, Supplementary Fig. 6). The strongest enrichment was observed in pancreatic ß-cell chromatin accessibility regions (OR = 4.1, 95% CI 2.6–6.4, BH-adjusted *p* = 1.93×10^-7^), while endocrine pancreas annotations were among the most enriched overall.

We next integrated fine-mapped credible sets with chromatin state maps from 223 biosamples from EpiMap^22^, and 218 single-cell chromatin accessibility annotations from CATlas^23^, including 107 brain-specific maps^24^, using TACTICAL^25^ to estimate tissue-of-action (TOA) scores for each GWAS signal. TOA scores broadly recapitulated known biology at established loci (Extended Data Fig. 7c–d, Supplementary Table 16). Among novel signals, the rare missense variant at *MC4R* (rs79783591) showed highest TOA scores brain tissues and neuronal cell types (glutamatergic neurons and GABAergic neurons), whereas *RFX6* (rs139343836) and *NR5A2* (rs140064850) showed highest TOA scores in pancreatic tissue and cell types. The *HNF1A* signal (rs483353044, PPA = 0.96) showed highest TOA scores in pancreatic acinar cells and stomach chief cells, while the *OPTC* signal (rs6685593) showed highest TOA scores in adipose tissue.

### Heritability and Polygenicity

We estimated SNP heritability 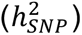 using several methods that account for genetic relatedness, admixture, and LD between imputed variants across the allele frequency spectrum (see Methods). In MCPS, 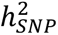 ranged from 0.20 to 0.31 (Fig. 4a, Supplementary Figs. 7–8)—exceeding estimates from European-descent and trans-ancestry studies (0.18-0.19^1,14^), even at the lower bound, and aligning closely with the estimate from the SIGMA Mexican cohort (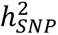 = 0.26, SD = 0.07^12^).

**Fig. 4.**
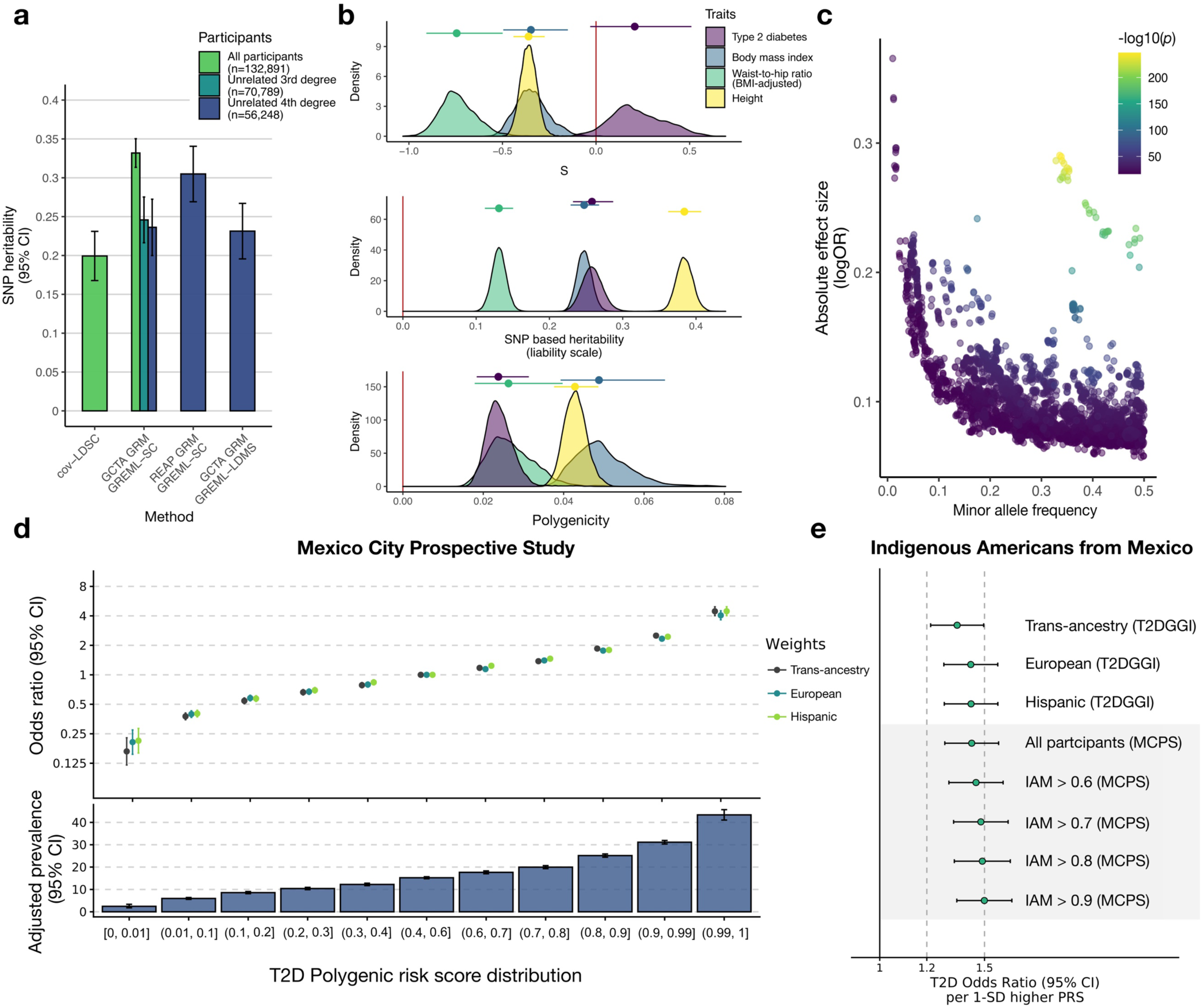
Heritability estimation and polygenic risk score evaluation. **a**, SNP-heritability 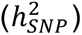 of T2D in MCPS. Columns show liability-scale heritability estimates by method and degree of relatedness (prevalence = 0.12). 95% confidence intervals are shown. **b,** Posterior distributions of genetic architecture parameters (polygenicity and *S*) estimated using GCTB-BayesS. Points indicate posterior medians and lines indicate 95% credible intervals. **c,** Relationship between minor allele frequency (MAF) and absolute effect size for T2D-associated variants (MAF > 1%; *p* < 5×10^-8^). Points are coloured by –log10(*p*). **d,** Association of polygenic risk scores (PRSs) from T2DGGI with T2D in MCPS. Odds ratios (ORs) were estimated using logistic regression adjusted for age, sex, BMI, district of residence and genetic principal components. The upper panel shows risk by PRS percentile and the lower panel shows adjusted prevalence for the trans-ancestry PRS. **e,** Association of PRSs with T2D in the MAIS cohort using weights from T2DGGI and MCPS-derived weights estimated in subsets of participants stratified by proportion of Indigenous American ancestry. ORs correspond to a one standard deviation increase in PRS. Abbreviations: cov-LDSC, covariate-adjusted LD score regression; GREML-SC, single-component GREML; GREML-LDMS, LD- and MAF-stratified GREML; REAP, relatedness estimation in admixed populations; IAM, Indigenous American.

To further characterise genetic architecture, we applied GCTB-BayesS^26^, which jointly estimates 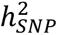, polygenicity (the proportion of SNPs with non-zero effects), and the relationship between effect size and MAF (*S*). Estimates of 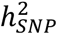 were consistent with other methods (median posterior 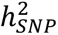 = 0.26, 95% CI 0.23–0.29), and the median posterior polygenicity was 0.02 (95% CI 0.02–0.03), consistent with previous studies^27,28^ (Fig. 4b). We observed a positive median posterior *S* estimate (0.21, 95% CI −0.03–0.51) together with a depletion of 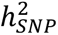 from low frequency variants (0.01 < MAF ≤ 0.1) (Extended Data Fig. 8a–b). Including variants with MAF > 0.001 strengthened this signal (*S* = 0.29; 95% CI 0.07–0.55), which remained positive after excluding GWAS index variants and their LD proxies (Extended Data Fig. 8c), indicating that effect sizes tend to increase with MAF. By contrast, *S* was negative for height (−0.36; 95% CI −0.44 to −0.27), BMI (−0.35; 95% CI −0.50 to −0.15), and BMI-adjusted waist-to-hip ratio (−0.74; 95% CI −0.91 to −0.50), indicating the opposite relationship between effect size and allele frequency for these traits (Fig. 4b, Supplementary Table 17). Together with our estimates of phenotypic variance explained by GWAS-significant variants, these results suggest that the detectable genetic architecture of T2D in MCPS is driven primarily by common variants with disproportionately large effects on disease risk (Fig. 4c).

### Polygenic risk scores

To assess polygenic risk score (PRS) performance in MCPS, we calculated scores using weights from a trans-ancestry GWAS meta-analysis comprising up to 2.5 million participants (∼40% non-European^3^) (Extended Data Fig. 9a–b). PRSs were strongly associated with T2D independent of age, sex, BMI and district of residence. Individuals in the top 1% of the PRS distribution had more than fourfold increased risk of T2D compared with those in the middle fifth (OR = 4.46, 95% CI 4.02–4.95 for the trans-ancestry PRS), whereas those in the bottom 1% had approximately one-sixth the risk (OR = 0.17, 95% CI 0.12–0.23) (Fig. 4d, Supplementary Table 18). The adjusted prevalence of T2D ranged from 2% (95% CI 0.02–0.03) in the bottom 1% of the PRS distribution to 43% (95% CI 0.41–0.45) in the top 1% (Fig. 4d, Supplementary Table 19). Predictive performance was similar across PRS weight sources, T2D definitions and relatedness thresholds (AUC 0.741–0.755; Supplementary Table 20, Supplementary Fig. 9), but was stronger at younger ages, particularly among women (Extended Data Fig. 10a–b, Supplementary Table 21). Adding the trans-ancestry PRS to a model including age, sex, BMI and district increased the Nagelkerke *R^2^* from 0.071 to 0.119 (Δ*R^2^*= 0.048; likelihood ratio test *χ*^2^(1) = 6382.1, *p* < 1×10^-16^), corresponding to an additional 8.7% of variance explained on the liability scale (assuming a population prevalence of 0.12^29^).

We next evaluated PRS performance in an independent sample from the MAIS study, comprising 661 cases and 1,292 controls from Indigenous communities across Mexico^9^. In addition to PRSs based on T2DGGI weights, we evaluated scores using weights derived from MCPS, including subsets of participants with high Indigenous American ancestry (Extended Data Fig. 9c–d). All PRSs were significantly associated with T2D in MAIS, with effect sizes ranging from OR = 1.35 (95% CI 1.22–1.49 per s.d. increase) for the T2DGGI trans-ancestry PRS to OR = 1.50 (95% CI 1.35–1.67) for the PRS derived from MCPS participants with >90% IAM ancestry (Fig. 4e, Supplementary Table 22).

## Discussion

Our study of the MCPS cohort significantly enhances Latin American representation in genomic mapping studies of T2D, addressing a critical gap in global genetic research. The effective sample size of T2D in our GWAS was more than double that for H/L cohorts from the trans-ancestry GWAS meta-analysis by the DIAMANTE consortium^2^, and alone is ∼90% that of the combined sample size of 17 H/L cohorts within the T2DGGI meta-analysis of 2.5M participants^3^. Moreover, MCPS contains a high degree of Mesoamerican admixture from central and southern Mexico and is enriched for genetic variation poorly represented in other globally diverse sequencing studies^10^. Following deep imputation to the TOPMed reference panel and stringent quality control, we identified 86 independent GWAS signals. Despite the relatively modest sample size compared to studies of European ancestry, we identified 21 novel associations, 15 of which replicated in meta-analyses of external cohorts, and the majority harboured risk alleles that were more frequent on IAM ancestral backgrounds. Furthermore, PheWAS and integrative analyses offered biological insight into locus-specific pathophysiology and reinforced the central role of genetic regulatory effects in the endocrine pancreas in shaping T2D susceptibility in Mexico.

Novel signals discovered in MCPS corroborate the diabetes relevance of various cellular pathways. For example, the deleterious missense variant at *WDFY2* further implicates an endosomal protein—WD repeat and FYVE domain-containing 2—involved in differentiation and insulin-stimulated glucose uptake within adipocytes^30,31^, and regulation of insulin sensitivity, gluconeogenesis and glycogen accumulation in hepatocytes^32^. To our knowledge, this is the first GWAS study providing human genetic evidence linking variation in *WDFY2* to T2D risk. Moreover, the T2D association with the missense variant at *RFX6* reinforces the emerging importance of the transcription factor Regulatory Factor X, 6, to T2D pathogenesis. Notably, a recent multimodal study of single-cell transcriptomics in purified pancreatic α and β cells, chromatin accessibility, and *ex vivo* and *in vivo* functional assays identified *RFX6 as* a hub transcription factor with a central role in the early stages of T2D, characterised by intrinsic β-cell dysfunction without β-cell loss^33^. Moreover, knockdown of *RFX6* in human pseudoislets impaired glucose-stimulated insulin secretion (GSIS) and altered β-cell chromatin structure. A separate knockdown study in purified α-cells indicated that *RFX6* also plays a role in glucagon secretion and exocytosis^34^. Our integrative functional analysis suggested that the most likely “tissue-of-action” for the novel *RFX6* signal in MCPS was endocrine pancreas. Furthermore, *in silico* analysis indicated that this variant alters a highly conserved residue within a domain important for RFX6 dimerization.

We referred to the NHGRI-EBI GWAS Catalog to characterise signals, which introduces caveats to the interpretation of putatively novel variants. Of note, we classified rs139343836 at *RFX6* as a *novel* signal at a *novel* locus. Although it is indeed the case that this variant has not been previously reported in the literature, a recent study of half a million Finnish individuals reported a rare frameshift mutation associated with T2D at *RFX6* (p.His293LeufsTer7; AF = 0.15%, OR = 3.7, *P* = 1.2×10^−10^)^35^. A subsequent study found that haploinsufficiency of this variant in CRISPR-engineered isogenic human stem cell-derived islets impaired GSIS and reduced calcium signaling^36^. Moreover, we reported missense variants at *MC4R* (rs79783591) and *HNF1A* (rs483353044) as *novel* signals at *known* loci as both were not listed as significant T2D variants in the catalog. However, the missense rs79783591 variant (p.Ile269Asn) in *MC4R*—encoding the appetite-regulating Melanocortin 4 Receptor—was previously found to associate with both childhood and adult obesity in a Mexican cohort 1,613 children and 3,932 adults^37^. Moreover, this variant was found to associate with T2D in a study of 3,754 Mexican adults (OR = 2.00, 95% CI 1.35–2.97, *P* = 0.00057) through both obesity-dependent and - independent routes^38^. This latter finding is notable as this variant did not map closely with known obesity-associated variants at *MC4R* (rs71336381) and *FTO* (rs1558902) in our physiological clustering analysis based on its distinct metabolomic profile. This variant was also recently reported as associated with youth-onset T2D (OR = 2.93, *P* = 2×10^-6^) in a trans-ancestry exome study^39^. Similarly, the rs483353044 missense variant (p.Glu508Lys) at *HNF1A*—the gene responsible for MODY3 which encodes Hepatocyte Nuclear Factor 1-α—was found to be associated with T2D in an exome study of 3,756 H/L participants from Mexico and the US (OR = 5.48; 95% CI 2.83–10.61; *P* = 4.4×10^-7^)^7^. Our analysis in MCPS is the first study where these consequential, rare variants attain genome-wide significance.

Diabetes is a major contributor to morbidity throughout the Americas, and is a leading cause of premature death in Mexico^40^. Here, we resolve important aspects of the genetic basis of T2D in this population. The proportion of inter-individual differences in T2D status due to genetic markers (i.e., SNP heritability) is substantial in MCPS, in line with a previous study in Mexico^12^ but exceeding that reported in European-descent cohorts^1,14^.

Moreover, we show that the “genetic architecture” of T2D in this cohort is predominantly driven by variants segregating in Mexico at higher frequencies. This is evidenced by: 1) a higher liability explained by common SNPs among genome-wide significant variants; 2) a large effect of polygenic burden due to variants shared across global populations on T2D risk; and 3) a positive estimate for the *S* parameter. This is contrary to the relationship observed for T2D in the UK Biobank^26^, and for other complex traits evaluated in MCPS. However, this may not be too unexpected given the presence of common variants with outsized risk effects, such as the lead variant at *SLC16A11* (rs113748381, OR = 1.19, 95% CI 1.16-1.22, RAF = 0.35) and the novel variant at *RFX6* (OR = 1.26, 95% CI 1.20-1.32)—the latter of which having an RAF of 5% in MCPS and 8% among IAM ancestry.

Despite the pronounced contribution of common risk variants, our exome analyses identified novel rare variants at established “diabetes” genes, including two additional deleterious missense variants at *HNF1A* and a damaging missense variant at *SLC30A8* that is highly protective against T2D. This latter finding reinforces previous sequencing studies^17,18^ that reported similar protective associations at *SLC30A8*, which are thought to confer protection via enhanced insulin secretion through destabilized insulin granules in the setting of haploinsufficiency. Moreover, we found an ultra-rare, protein damaging variant in the known MODY2 gene *GCK*. Although such variants tend to cause MODY2, a relatively benign condition characterised by mildly elevated HbA1c but otherwise well-tolerated^15^, the missense variant rs1131691598 (p.Leu45Pro) strongly associated with T2D risk in MCPS and coincided with pathological levels of HbA1c (mean 7.7%, SD 1.6%) among carriers.

This comprehensive study underscores the value of genetic exploration in globally diverse yet underrepresented populations. The discovery of novel risk variants enriched in IAM ancestry provides insights into T2D biology, while also highlighting opportunities for precision diabetes medicine in Latino and Indigenous American populations. Rare, IAM-enriched T2D risk variants in MODY genes may inform clinical management, whereas polygenic scores validated in MCPS could more precisely identify individuals for targeted prevention. These findings further illustrate how broadening inclusion in genetic research can expand the impact of discovery and help ensure more equal benefits.

## Methods

Detailed descriptions of the MCPS cohort and analytical procedures, including admixture estimation, are provided in the Supplementary Methods.

### Ethics statement

The MCPS was approved by the Mexican Ministry of Health, the Mexican National Council of Science and Technology (0595 P-M), the Central Oxford Research Ethics Committee (C99.260), and the Ethics and Research Commissions of the Faculty of Medicine at the National Autonomous University of Mexico (FMED/CI/SPLR/067/2015). Transport and long-term storage of blood samples (plasma and buffy coat) at the Clinical Trial Service Unit sample archive, University of Oxford, were approved by the Mexican Ministry of Health. All participants provided written informed consent.

### Phenotype selection

Case and control sets of MCPS participants were defined using information recorded at baseline. T2D cases were participants with a previous diagnosis of diabetes and/or taking glucose-lowering medication. Participants with an estimated age at diagnosis <35 years who were taking insulin at recruitment were presumed to have type 1 diabetes and excluded. Normoglycemic controls were defined as participants with HbA1c < 6.0% recorded at baseline. Individuals with prediabetes (HbA1c 6.1–6.4%) or undiagnosed diabetes (HbA1c ≥ 6.5%) were excluded. After restricting analysis to the 140,829 participants included in the quality-controlled array genotyping dataset (see Supplementary Methods), there were 19,431 T2D cases and 105,611 normoglycemic controls remained for genetic association analyses.

### Genotyping array quality control

Genotyping was performed using the Illumina Infinium Global Screening Array (GSA) v2.0 at the Regeneron Genetics Center, generating genotype data for 140,831 participants and of 650,381 variants^10^. Quality control followed a previously described framework^10^ with minor modifications that increased sample retention by 2,318 participants (Supplementary Table 23). The resulting dataset comprised 140,829 participants, 539,448 autosomal variants and 18,560 chromosome X variants.

### Relatedness

Pairwise relatedness was inferred from shared identical-by-descent (IBD) segments using KING v2.2.7. Relationships were identified up to the third degree (IBD proportion > 0.0884). To generate a maximum set of individuals unrelated up to the third degree, independent vertex set (IVS) analysis was applied to pairwise relatedness networks (Supplementary Methods). Participants selected by IVS analysis were then combined with the 40,695 participants without any relatives in the cohort up to the third degree. This procedure yielded a set of 79,612 individuals, hereafter referred to as the maximum unrelated set.

### Principal component analysis

Principal component analysis (PCA) was performed using the workflow of Privé et al.^41^ implemented in the R package *bigsnpr*. Principal component scores and loadings were estimated from the scaled genotype matrix of 40,695 unrelated individuals using truncated singular value decomposition. PCA models were evaluated across combinations of LD-clumping (*r*^2^ = 0.2, 0.01 and 0.005) and MAF (0.01 and 0.05) thresholds, and the model using LD-clumping *r*^2^ = 0.005 and MAF = 0.05 (12,927 variants) was selected because it maximized the number of principal components exhibiting loading distributions consistent with population structure rather than local LD structure (Supplementary Figs. 10–11). No sample outliers were identified, and the remaining 100,134 related individuals were projected into the resulting PC space using the Online Augmentation, Decomposition and Procrustes (OADP) algorithm^41^. Based on their loading distributions, the first seven PCs were selected as covariates in genetic association analyses

### Imputation and variant selection

Imputation was performed using the TOPMed Imputation Server^11^ with the TOPMed reference panel (version r2), comprising 97,256 whole-genome sequenced samples and 308,107,085 autosomal and chromosome X variants. Haplotype phasing and genotype imputation were performed with Eagle v2.4 and Minimac4, respectively. Variants were retained for association analyses if they met two criteria: (1) imputation INFO *r^2^* > 0.4 and (2) effective sample size (*N_eff_*) > 50, where

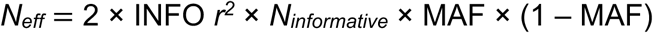

and *N_informative_* is the number of T2D cases. This *Neff* threshold was also applied to binary traits in the PAGE multi-ancestry GWAS study^42^. A total of 15,578,291 variants (15,054,525 autosomal and 523,766 chromosome X variants) met these criteria.

### Genome-wide association studies

Genetic association analyses of autosomal and chromosome X variants were performed using SUGEN^43^. To account for extensive relatedness in MCPS, family networks were delineated using pairwise IBD estimates (Supplementary Methods) and incorporated into the association model. A logistic regression model with T2D status as the outcome was fitted using the “model-based” variance specification. Age, sex and the first seven genetic principal components were included as covariates. Additional analyses were performed with further adjustment for BMI or waist-to-hip ratio.

### Covariate-adjusted LD score regression

To distinguish polygenicity from inflation arising from population structure or relatedness, we applied covariate-adjusted LD score regression (cov-LDSC)^12^. Covariate-adjusted LD scores were estimated using whole-genome sequence data from 9,764 MCPS participants^10^. Principal components were incorporated during LD score estimation as described previously^12^. Cov-LD scores were calculated for 18,789,983 variants with MAF ≥ 0.1% using 1-Mb windows.

### Dissection of independent GWAS signals

T2D-associated loci were defined as non-overlapping genomic windows extending 500 kb around variants attaining genome-wide significance (*p* < 5×10^-8^). The index variant for each locus was defined as the variant with the lowest *p* value, and loci were annotated using the nearest protein-coding ENSEMBL v105 gene. Independent association signals were identified using approximate conditional analysis implemented in GCTA-COJO (v1.94). Linkage disequilibrium was estimated using TOPMed-imputed genotypes from the 79,162 MCPS participants comprising the “maximum unrelated set” (see Supplementary Information). Stepwise selection was performed until no additional variants attained conditional genome-wide significance and all retained variants remained significant in the joint model.

### Variant annotation

Trait-associated variants were annotated using the Ensembl Variant Effect Predictor (VEP) (URL) with Ensembl version 105 annotations for GRCh38. Annotations corresponded to Ensembl canonical genes and predicted functional impact scores (SIFT, Polyphen, PrimateAI, CADD, REVEL and SpliceAI) were extracted. Effect allele frequencies were estimated from the maximum unrelated set of MCPS participants. Ancestry-specific allele frequencies were obtained from the MCPS Variant Browser (URL), while population frequencies from gnomAD exome and whole-genome sequencing datasets were obtained through VEP.

### Identification of novel GWAS signals

Novelty was assessed using previously reported T2D-associated variants obtained from the NHGRI-EBI Catalog (accessed August 21, 2022) and supplemented with genome-wide significant variants from the DIAMANTE trans-ancestry meta-analysis of 1.3M individuals^2^. GWAS signals were classified as: (1) reported, if the index variant had been previously associated with T2D; (2) known locus, if the index variant was located within 500 kb of a previously reported T2D variant; (3) novel locus, if the index variant was located more than 500 kb from any previously reported variant and at least two other variants within the locus attained a *p* < 1×10^-5^; or (4) undetermined.

To identify novel associations at known loci, approximate conditional analyses were performed using GCTA-COJO. Index variants were assessed for genome-wide significant associations after conditioning on previously reported T2D-associated variants located within 500 kb. Signals that remained significant after conditioning (*p* < 5×10^-8^) were considered novel associations at known loci. Following release of the GWAS meta-analysis of 2.5M individuals by T2DGGI^3^, variants classified as novel in MCPS were confirmed not to be reported as index variants in that study.

### Genetic fine-mapping

Independent GWAS signals were fine-mapped using an approximate Bayesian approach^14,44^. Signals were included if the index variant had MAF > 0.0025 and was located outside the MHC region. For loci harbouring multiple independent signals, conditional association statistics were obtained for each signal using GCTA-COJO by conditioning on all remaining signals at the locus. Approximate Bayes factors, posterior probabilities of association (PPAs), and 99% credible sets were then estimated as described previously^14,44^. Fine-mapping resolution was assessed using both the number of variants in each credible set and the maximum PPA.

To compare fine-mapping resolution between MCPS and DIAGRAM^14^, 99% credible sets were obtained from DIAGRAM and mapped to GRCh38 coordinates. Shared signals were identified using LD between index variants estimated in the maximum unrelated set of MCPS participants, with signals considered shared at LD *r^2^* ≥ 0.4.

### Phenotypic variance explained by GWAS signals

The phenotypic variance explained by GWAS-significant variants was estimated within the maximum unrelated set of MCPS participants. Separate estimates were obtained for common (MAF ≥ 0.05) and low-frequency or rare (MAF < 0.05) index variants. T2D status was modelled using logistic regression with age, sex and the first seven genetic principal components as covariates, with and without inclusion of the corresponding index variants. Nagelkerke’s *R^2^* was calculated from the resulting models and converted to variance explained on the liability scale using the method of Lee et al.^45^, assuming a case fraction of 0.155 and a population prevalence of 0.12 for T2D in Mexico^29^.

### Exome-wide association study

Exome-wide association analysis was performed in 141,045 MCPS participants using 9,325,897 variants that passed previous quality-control procedures^10^. The single-variant test was conducted using REGENIE v3.1.3. A whole-genome regression model was fitted using quality-controlled array variants (MAC ≥100; 560,015 variants) and incorporated into association testing using the leave-one-chromosome-out (LOCO) framework. Association testing was restricted to variants with MAC ≥ 25, and an approximate Firth correction was applied to variants with *p* < 0.01. Analysis was performed conditional on genome-wide significant GWAS variants present in the exome variant set (249 overlapping variants). The logistic regression model was adjusted for age, sex, and the first seven genetic PCs.

### Gene-based rare variant testing

Exome variants were annotated according to the BRaVa Consortium framework, which integrates VEP annotations, CADD scores, and SpliceAI predictions. Gene-based analyses considered two classes of potentially deleterious variants: (1) putative loss-of-function (pLoF) variants and (2) damaging missense or protein-altering variants (Supplementary Methods).

Gene-based tests were performed using REGENIE v3.1.3 with the same genome-wide predictors and covariates used for the ExWAS. Four variant masks were evaluated per gene, corresponding to pLoF-only or pLoF-plus-damaging variants restricted to either singletons or variants with MAF < 0.1%. Associations with T2D were assessed using the “Gene P” test^46^, which combines burden, SKAT-O and ACAT-based test statistics into a single gene-level *p* value. Genes with *p* ≤ 2.73 × 10^-6^ (Bonferroni correction for 18,262 protein-coding genes) were considered significant.

### Replication meta-analyses

To assess replication of novel T2D-associated variants identified in MCPS, association summary statistics were obtained from publicly available resources and collaborating studies, including Mexico Biobank, MAIS, MVP, T2DGGI, DIAMANTE, PAGE, AGEN and Pan-UK Biobank (Supplementary Methods and Supplementary Table 7). Summary statistics were harmonised across studies with respect to effect and non-effect alleles in MCPS. Trans-ancestry effect estimates for MVP were derived using inverse-variance-weighted (IVW) meta-analysis across ancestry-specific GWAS results. Per-variant replication meta-analyses were performed using IVW meta-analysis and restricted to the largest available non-overlapping studies. For example, DIAMANTE, MVP, PAGE, AGEN and Pan-UK Biobank results were excluded when variants were available in T2DGGI because of cohort overlap. Three meta-analyses were performed for each variant corresponding to: (1) Hispanic/Latino cohorts, (2) non-Hispanic/Latino cohorts and (3) all non-overlapping cohorts combined.

Heterogeneity between Hispanic/Latino and non-Hispanic/Latino effect estimates was assessed using Cochran’s Q test implemented in the *meta* R package and by estimating between-group difference *p* values as described previously^47^. Meta-analysis and heterogeneity *p* values were adjusted using the Benjamini–Hochberg procedure. Novel MCPS signals were considered replicated if effect estimates were directionally consistent and had an adjusted *p* ≤ 0.05 in the combined replication meta-analysis.

### Phenome-wide association study

Each of the 86 T2D-associated index variants was tested for association with 130 traits in MCPS, including HbA1c, blood pressure, anthropometric traits, baseline disease outcomes, all-cause mortality, and 110 metabolite features measured on the Nightingale platform. Quantitative traits were transformed by rank-based inverse normalisation separately in men and women and adjusted for age and the first seven genetic principal components. Metabolite traits were additionally adjusted for fasting duration.

Analysis of quantitative traits were restricted to participants free of major chronic disease at baseline and excluded individuals with undiagnosed diabetes, missing phenotype data or implausible anthropometric measurements (Supplementary Methods). Sample sizes ranged from 5,764 to 107,296 participants, depending on trait availability. Linear and logistic regression were used for quantitative and disease traits, respectively, while all-cause mortality was analysed using Cox proportional hazards regression. All association analyses were performed using SUGEN.

### Functional integration at T2D loci

Tissue-of-action (TOA) scores were estimated using TACTICAL^25^, which integrates genetic credible sets with tissue- and cell-type-specific epigenomic annotations. We incorporated chromatin state maps from 223 adult biosamples (28 tissues) generated by EpiMap and 218 single-nucleus ATAC-seq maps spanning 26 unique tissues and 107 brain cell types the *Cis* Element Atlas. TOA scores were calculated by partitioning posterior probabilities of association from 99% credible sets across weighted epigenomic annotations, as described previously^25^. Analysis were performed at the level of both individual biosamples or cell types and across whole tissues.

### Heritability

SNP heritability 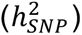 attributable to variants genotyped on the GSAv2 array was estimated using restricted maximum likelihood as implemented in GCTA v1.94.0. Analyses were performed in the full GWAS sample and in subsets of increasingly unrelated participants. Models included age, sex and the seven genetic principal components as covariates. To account for recent admixture, heritability was also estimated using admixture-adjusted genetic relatedness matrices calculated with REAP^48^.

Multicomponent SNP heritability was estimated using GREML-LDMS^49^ applied to variants imputed from the TOPMed reference panel and stratified by MAF and LD-score bins. A disease prevalence of 0.12 was assumed for all liability-scale estimates.

### Polygenicity

Polygenicity analyses were performed using GCTB-BayesS^26^, which models SNP effects using a mixture prior comprising a normal distribution and a point mass at zero. The unknown mixing probability, *Pi*, provides an estimate of polygenicity, while the *S* parameter captures the relationship between effect size and MAF. Negative values of S indicate larger effects among lower-frequency variants, consistent with negative selection, whereas positive values indicate the opposite relationship. Posterior distributions were estimated using Markov chain Monte Carlo with 50,000 iterations and 2,500 burn-ins. The posterior median was used as the point estimate and the highest posterior density was used to construct 95% credible intervals.

Analyses were performed in the set of 56,248 MCPS participants unrelated to the fourth degree (8,223 cases and 48,025 controls). A total of 333,195 genotyped variants (GSAv2 array) were retained after excluding variants with MAF < 1%, HWE *p* < 10^-6^, or genotype missingness > 5%. Models included age, sex, and the first seven genetic principal components as covariates. Comparable analyses were also performed for height, BMI, and BMI-adjusted waist-to-hip ratio.

### Polygenic risk scores

Polygenic risk scores (PRSs) were evaluated in 138,026 MCPS participants with complete covariate data, including 26,119 T2D cases and 111,907 controls. Cases included participants with either previously diagnosed diabetes or undiagnosed diabetes defined by HbA1c > 6.5%, whereas controls were participants without T2D. PRSs were calculated using the PGS Catalog^50^ calculator and conditionally independent variants from the trans-ancestry GWAS meta-analysis of 2.5M individuals by the T2D Global Genomics Initiative (T2DGGI)^3^. Trans-ancestry, European and Hispanic effect estimates from T2DGGI were used to generate ancestry-specific PRSs. Associations between PRSs and T2D were evaluated using logistic regression models adjusted for age, sex, BMI, district of residence and the first seven genetic principal components. Predictive performance was assessed using the area under the receiver operating characteristic curve (AUC) and compared across PRS weight sources, T2D definitions and relatedness thresholds. Analyses were additionally stratified by sex and age.

PRS transferability was evaluated in 1,953 participants from the Metabolic Analysis in an Indigenous Sample (MAIS) study (661 cases and 1,292 controls). In addition to T2DGGI-derived weights, PRSs were generated using effect estimates derived from MCPS and from subsets of MCPS participants with progressively higher Indigenous American ancestry proportions (>60%, >70%, >80% and >90%). MCPS-derived weights were estimated using SUGEN with adjustment for age, sex and the first seven genetic PCs. Associations with T2D in MAIS were evaluated using logistic regression models adjusted for age, sex, BMI and the first ten genetic PCs.

## URLs

AGEN: https://blog.nus.edu.sg/agen/

*Cis* Element Atlas: https://catlas.org

CMDGA: https://cmdga.org/

DIAGRAM Consortium website (source of DIAGRAM, DIAMANTE and T2DGGI summary statistics): https://diagram-consortium.org/

Ensembl Variant Effect Predictor (VEP): https://www.ensembl.org/Tools/VEP

EpiMap: https://compbio.mit.edu/epimap/

MCPS Variant Browser: https://rgc-mcps.regeneron.com/

MexVar platform: https://morenolab.shinyapps.io/mexvar/

NHGRI-EBI GWAS Catalog: https://www.ebi.ac.uk/gwas/

Pan-UK Biobank: https://pan.ukbb.broadinstitute.org/

RGC Million Exome Variant Browser: https://rgc-research.regeneron.com/me/

TOPMed Imputation Server: https://imputation.biodatacatalyst.nhlbi.nih.gov/

## Data availability

GWAS summary statistics will be made available upon publication through a dedicated project website. Covariate-adjusted LD scores derived in 9,764 whole-genome-sequenced MCPS participants will be made publicly available for download via a Zenodo repository upon publication. Data from the Mexico City Prospective Study are available to bona fide researchers. For more details, the study’s Data and Sample Sharing policy is available in English or Spanish from https://www.ctsu.ox.ac.uk/research/mcps. Available study data can be examined in detail through the study’s Data Showcase, available at https://datashare.ndph.ox.ac.uk/mexico/.

## Acknowledgments

We thank the participants and staff of the Mexico City Prospective Study (MCPS). MCPS has received funding from the Mexican Health Ministry, the National Council of Science and Technology for Mexico, Wellcome Trust (058299/Z/99), Cancer Research UK, British Heart Foundation (RE/13/1/30181), Kidney Research UK, and the UK Medical Research Council (MC_UU_00017/2, MR/Z504543/1). We also thank the members of the ENSA Genomics Consortium, who enabled the collection and processing of samples from the 2000 National Health Survey conducted by the National Institute of Public Health and contributed to the establishment of the Mexican Biobank (MXB), whose data were used in this work. J.E.B., K.E.N. and L.E.P. were partially supported by R01HL142302. F.B. was supported by Health Data Research UK which is funded by UK Research and Innovation, the Medical Research Council, the British Heart Foundation, Cancer Research UK, the National Institute for Health and Care Research, the Economic and Social Research Council, the Engineering and Physical Sciences Research Council, Health and Care Research Wales, Health and Social Care Research and Development Division (Public Health Agency, Northern Ireland), Chief Scientist Office of the Scottish Government Health and Social Care Directorates. This research was also supported by the Wellcome Trust Core Award Grant Number 203141/Z/16/Z with additional support from the NIHR Oxford BRC. The views expressed are those of the authors and not necessarily those of the NHS, the NIHR or the Department of Health.

## Author Contributions

Conceptualization: J.M.T., J.B., J.A.-D., J.R.E., P.K.-M., and R.T.-C. Software: J.M.T., S.M., and T.L. Formal analysis: J.M.T., D.A.-R., E.T., F.B.-O., S.M., M.T., and E.B. Validation: F.B.-O, H.G.-O., C.D.Q.-C., L.E.P., L.G.-G., C.A.A.-S., M.T.-L., A.Y.C., J.E.B., K.E.N., M.J.G., A.C.P., A.M.-E., and L.O. Investigation: J.M.T., J.B., D.A.-R., E.T., and S.M., Resources: J.R.E. Data curation: J.M.T. and M.T. Writing, original draft: J.M.T., D.A.-R., E.T., F.B.-O., and S.M. Writing, review and editing: all authors. Visualization: J.M.T., D.A.-R., E.T., and S.M. Supervision: J.M.T., J.B., J.A.-D., P.K.-M., J.R.E., and R.T.-C. Project administration: J.M.T. Funding acquisition: R.C., J.B., J.A.-D., P.K.-M., J.R.E., and R.T.-C.

## Competing interests

J.R.E. and R.C. report grants to the University of Oxford from AstraZeneca and Regeneron Pharmaceuticals. R.C. reports having a patent for a statin-related myopathy genetic test licensed to the University of Oxford from Boston Heart Diagnostics (R.C. has waived any personal reward with any share in royalty and other payments waived in favour of the Nuffield Department of Population Health, University of Oxford) and being chair of not-for-profit clinical trial company PROTAS, chief executive of UK Biobank, and chair of the steering committee of the ORION-4 clinical trial of inclisiran. A.R.S. is an employee of Regeneron Pharmaceuticals, Inc. and receives salary, stock and stock options as compensation for his employment. E.W. is a current employee and stockholder of AstraZeneca. All other authors declare no competing interests.

---

**Extended Data Fig. 1.**
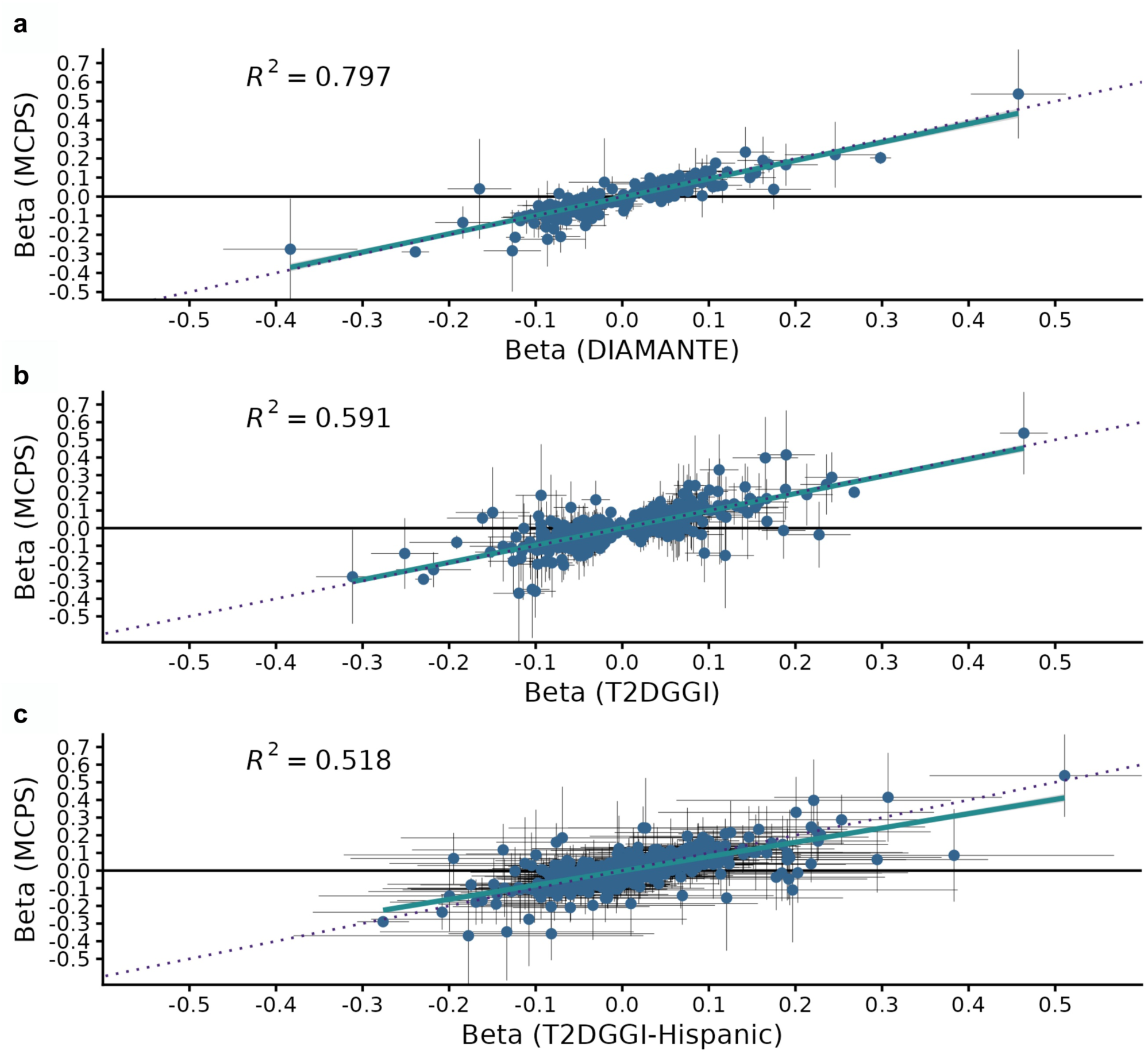
Effect size comparison at known T2D loci. **a**, Beta coefficients for index variants reported in a trans-ancestry meta-analysis of 1.3 million individuals by the DIAMANTE consortium (Mahajan et al., 2022) are shown on the x-axis, with corresponding estimates from MCPS on the y-axis. Error bars represent 95% confidence intervals. Of the 338 reported variants, 327 (96.7%) were available in the MCPS GWAS of T2D, and 217 (64.2%) replicated (directionally consistent with p < 0.05). Effect sizes were highly correlated (Pearson’s *r* = 0.89; *p* < 2.2×10^-16^). **b**, Comparison with index variants from a trans-ancestry meta-analysis of 2.5 million individuals by the T2DGGI consortium (Suzuki et al., 2024). Of the 1,289 reported variants, 1,229 (95.3%) were included in the MCPS GWAS, and 519 (40.3%) replicated. Effect sizes were strongly correlated (Pearson’s *r* = 0.77; *p* < 2.2×10^-16^). **c**, Comparison with beta coefficients from the meta-analysis of Hispanic participants within T2DGGI (N_cases_ = 29,375; N_controls_ = 59,368). Among the 1,289 index variants, 511 (38.6%) replicated and effect sizes were correlated between studies (Pearson’s *r* = 0.72; *p* < 2.2×10^-16^).

**Extended Data Fig. 2.**
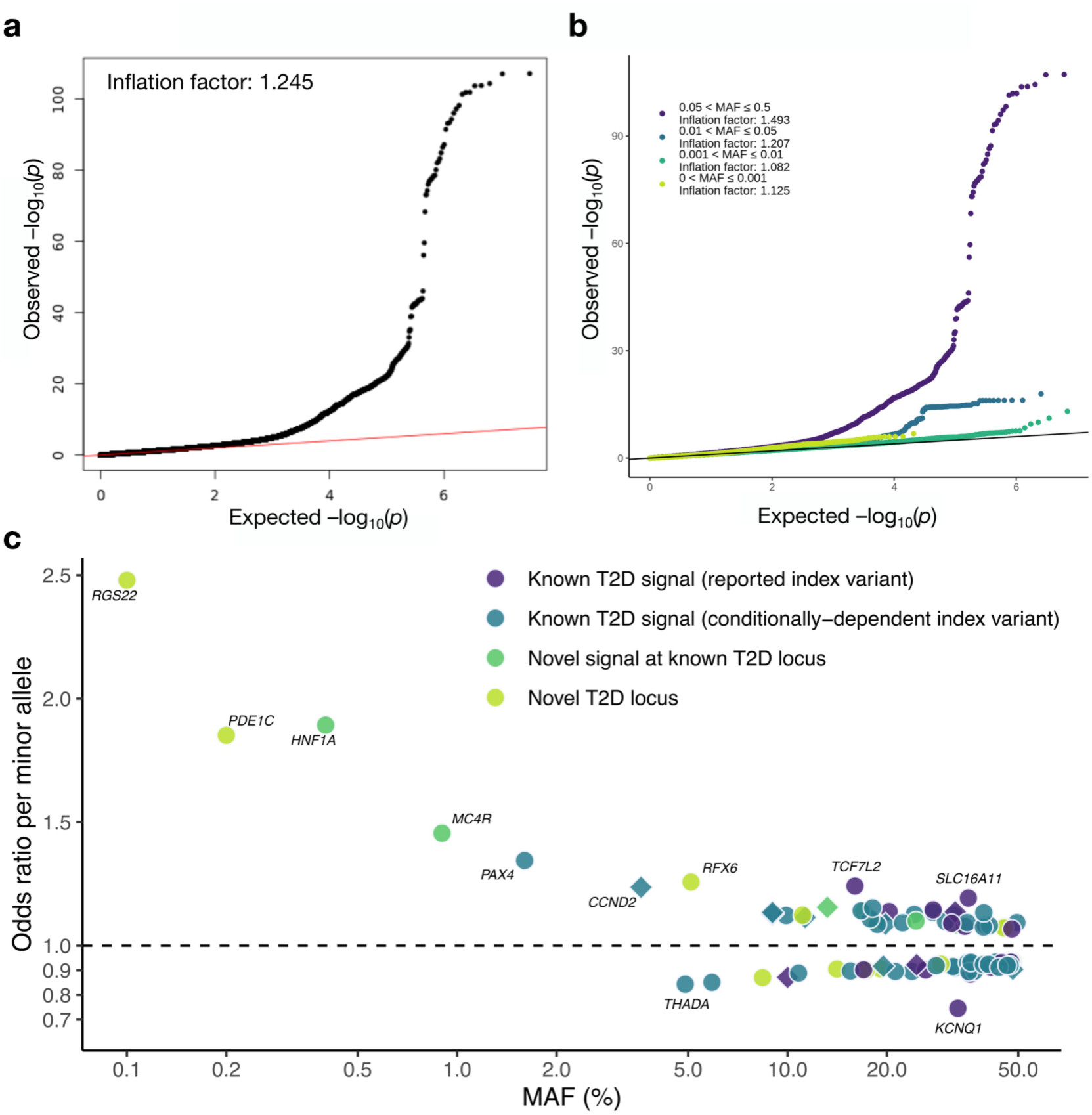
Distribution and effect sizes of GWAS associations for T2D. **a**, Quantile–quantile (QQ) plot of observed versus expected association *p* values for all variants. **b**, QQ plots stratified by minor allele frequency (MAF). **c**, Relationship between MAF and effect size for the 86 GWAS index variants. MAF (%) is shown on the x-axis and the per-minor-allele odds ratio (OR) on the y-axis. Points are coloured according to GWAS signal classification: previously reported T2D associations in the NHGRI-EBI GWAS Catalog (purple), known signals at known loci (blue), novel signals at known loci (green), and novel signals at novel loci (yellow). Both axes in **c** are shown on a log10 scale.

**Extended Data Fig. 3.**
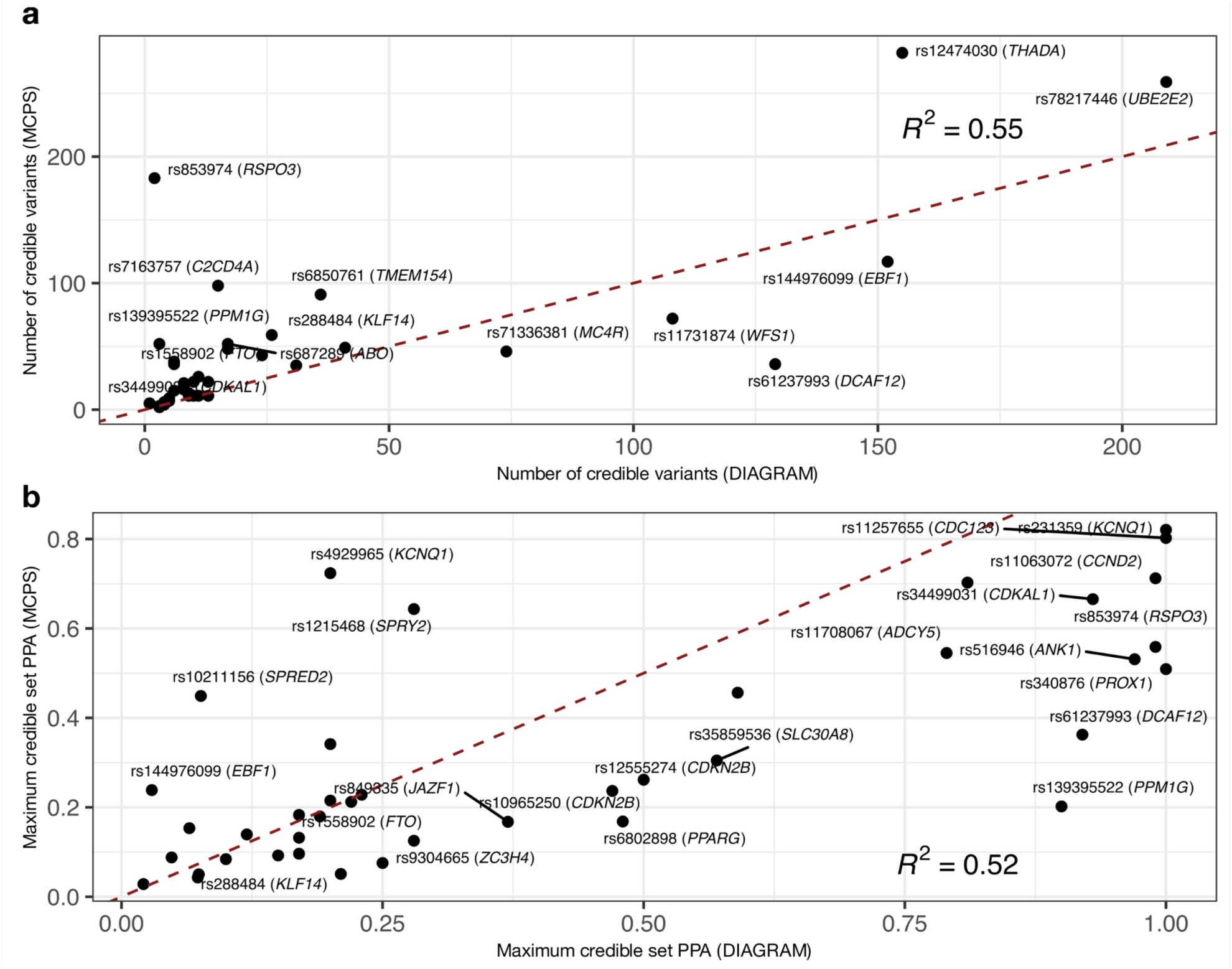
Comparison of fine-mapping resolution between MCPS and DIAGRAM. Forty GWAS signals from MCPS were matched to corresponding signals in DIAGRAM (LD *r^2^* ≥ 0.4 between index variants). **a**, Comparison of the number of variants in the 99% credible set. b, Comparison of the maximum posterior probability of association (PPA) within each credible set. Signals differing by more than 25 credible variants (**a**) or 0.15 PPA (**b**) are labelled with the index variant rsID and nearest protein-coding gene.

**Extended Data Fig. 4.**
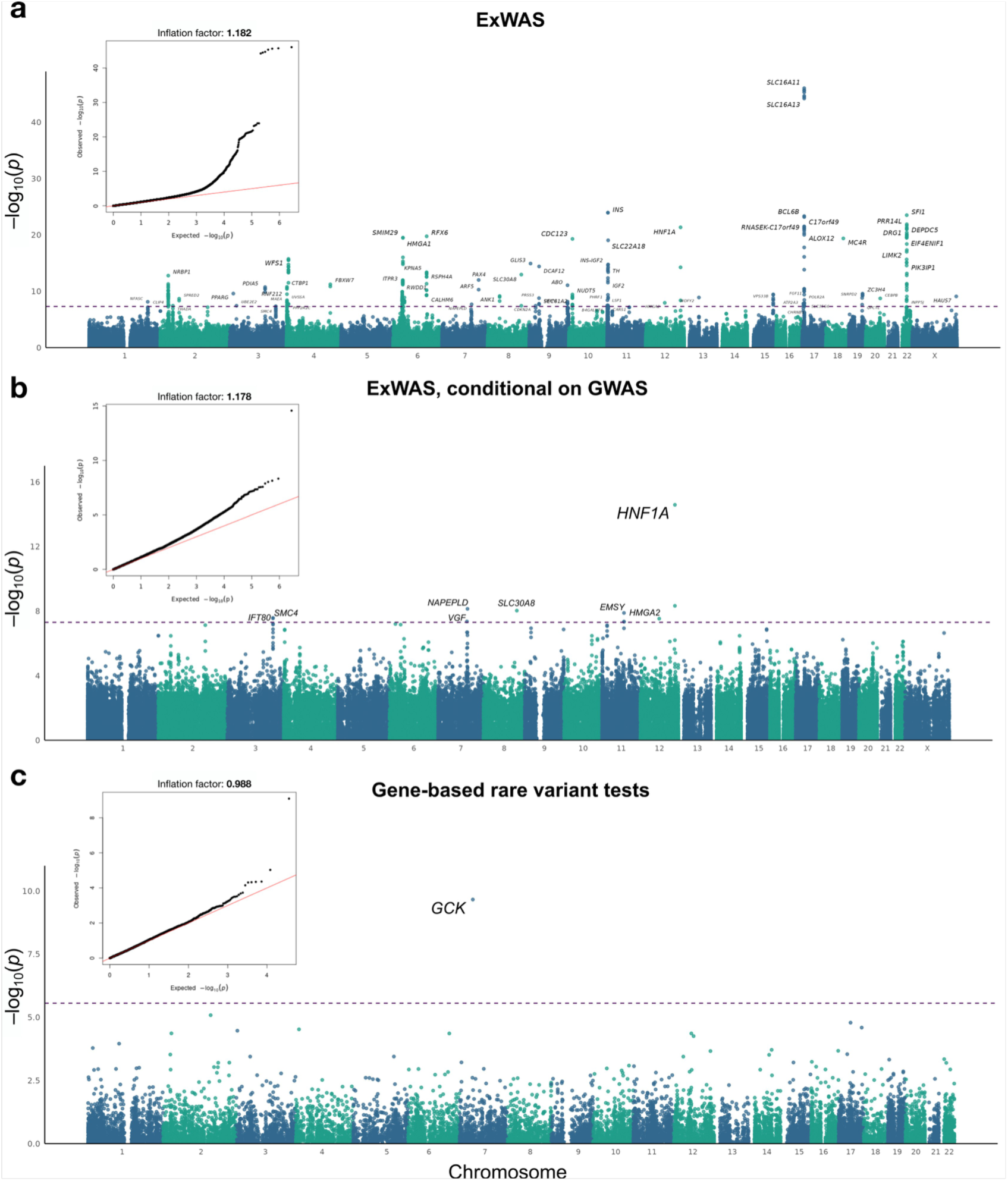
Exome-wide association analyses of T2D diabetes in MCPS. **a**, Manhattan plot of exome-wide association results for individual variants. **b**, Conditional exome-wide association results after adjustment for genome-wide significant GWAS variants. **c**, Gene-level associations from ultra-rare predicted loss-of-function (pLOF) or damaging missense variants. The y-axis shows –log_10_(*p*) values from single-variant association tests **(a, b)** or the "Gene P" test **(c)**. The standard genome-wide significance threshold (*p* < 5×10^-8^) was applied to exome-wide analyses of single variants, whereas a Bonferroni-adjusted threshold (*p* < 2.74×10^-6^), correcting for 18,262 genes, was applied to gene-level tests. Quantile-quantile plots are shown in the insets.

**Extended Data Fig. 5.**
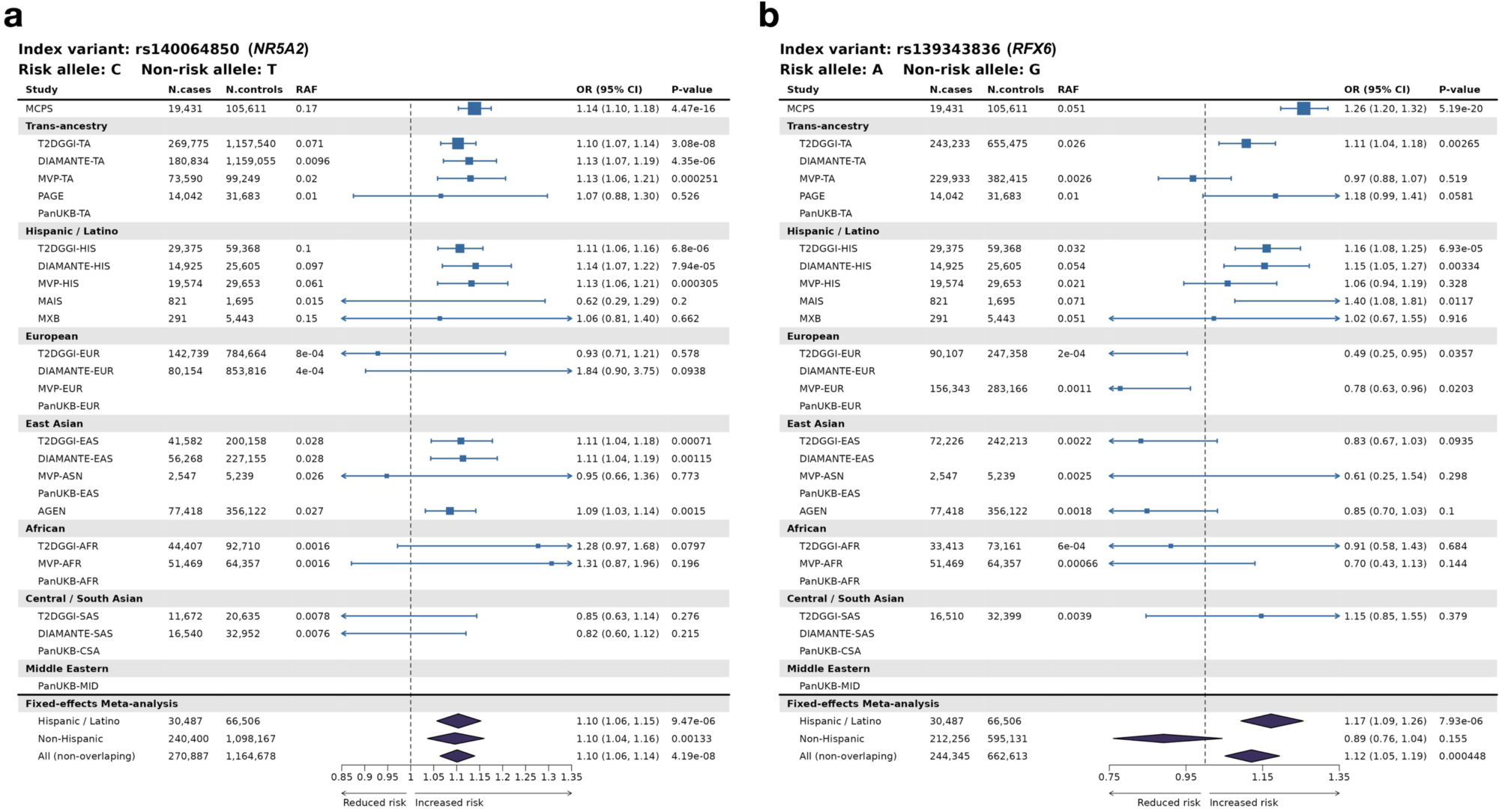
Replication of novel T2D associations at *NR5A2* and *RFX6*. Forest plots show effect estimates for the risk alleles at **a**, rs140064850 (*NR5A2*) and **b**, rs139343836 (*RFX6*) in MCPS and external studies, including T2DGGI, DIAMANTE, Pan-UK Biobank, PAGE, AGEN, MVP, Mexico Biobank (MXB), and Metabolic Analysis of an Indigenous Sample (MAIS). Fixed-effects inverse-variance-weighted meta-analysis results are shown for Hispanic/Latino, non-Hispanic/Latino and all non-overlapping cohorts. RAF, risk allele frequency; TA, trans-ancestry; HIS, Hispanic/Latino; EUR, European; EAS/ASN, East Asian; AFR, African/African American; CSA/SAS, Central or South Asian; MID, Middle Eastern.

**Extended Data Fig. 6.**
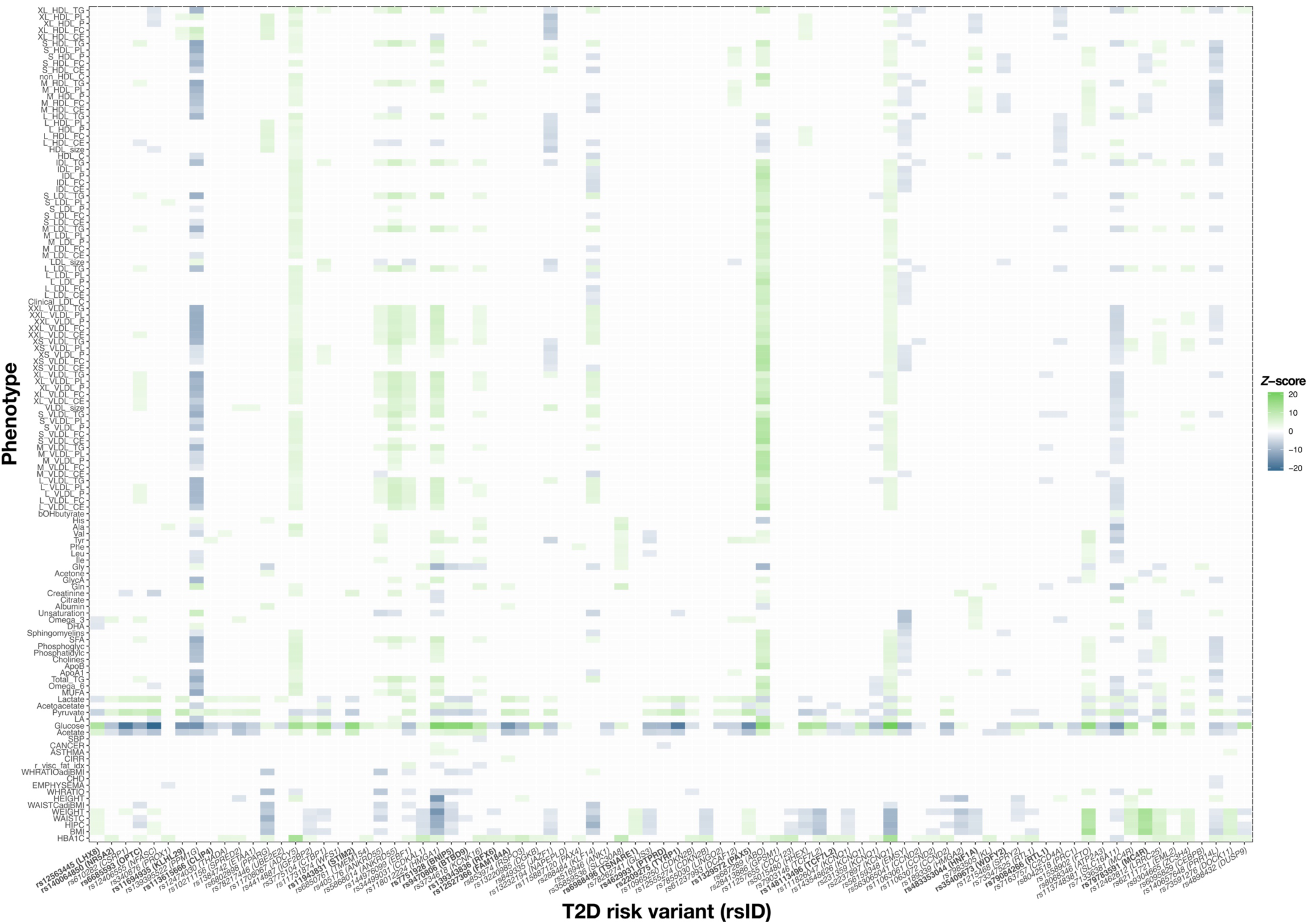
Phenome-wide association study of T2D-associated variants in MCPS. Associations were tested across 130 traits. Shown are all variant–trait associations with a Benjamini–Hochberg-adjusted *p* < 0.05. *Z* scores correspond to effects of the T2D risk allele. Bold text indicates rsIDs for novel T2D signals. Metabolite concentrations are reported in mmol l^-1^. XXL, extremely large particle; XL, very large particle; L, large particle; M, medium particle; S, small particle; XS, very small particle; HDL, high-density lipoprotein; LDL, low-density lipoprotein; IDL, intermediate-density lipoprotein; VLDL, very-low-density lipoprotein; P, particle concentration; PL, phospholipids; C, cholesterol; CE, cholesteryl esters; FC, free cholesterol; TG, triglycerides; DHA, docosahexaenoic acid; LA, linoleic acid; SFA, saturated fatty acids; MUFA, monounsaturated fatty acids; PUFA, polyunsaturated fatty acids; Unsaturation, degree of unsaturation; ApoB, apolipoprotein B; ApoA1, apolipoprotein A1; Ala, alanine; Gln, glutamine; Gly, glycine; His, histidine; Total_BCAA, total branched-chain amino acids; Ile, isoleucine; Leu, leucine; Val, valine; Phe, phenylalanine; Tyr, tyrosine; GlycA, glycoprotein acetyls; BMI, body mass index; WHR, waist-to-hip ratio; HIPC, hip circumference; WAISTC, waist circumference; adjBMI, adjusted for BMI; HbA1c, haemoglobin A1c.

**Extended Data Fig. 7.**
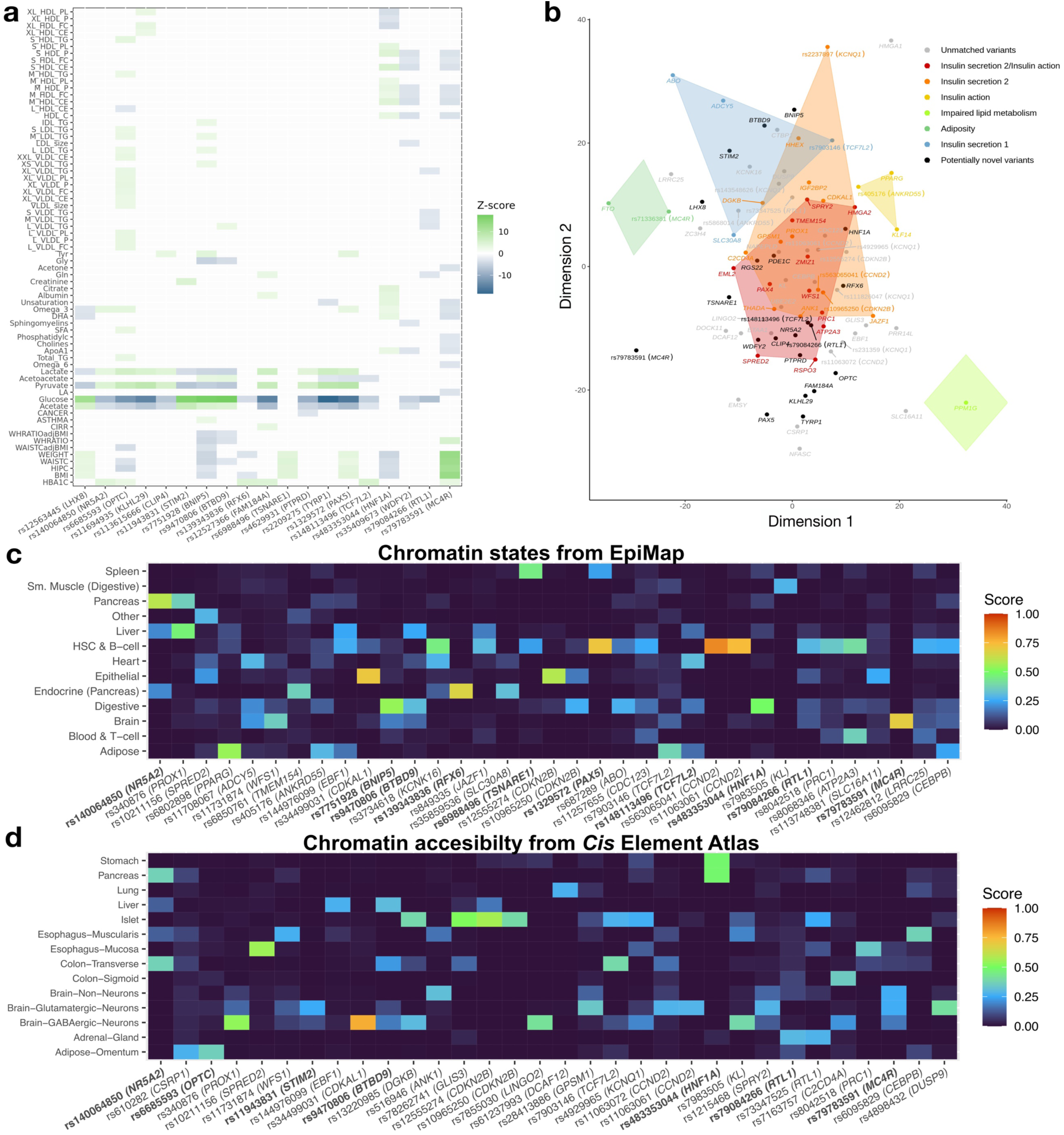
Physiological and epigenomic profiling of T2D GWAS signals in MCPS. **a**, Phenome-wide association study (PheWAS) results for the 21 novel GWAS signals. Associations were tested across 130 traits, with results shown for variant–trait pairs passing a Benjamini–Hochberg adjusted *p* < 0.05. *Z*-scores correspond to the T2D risk allele; metabolite concentrations are in mmol/L. **b**, Multidimensional scaling of PheWAS *Z*-scores for 51 continuous traits in MCPS. Shown are 86 conditionally independent variants; bold labels indicate novel variants. Colors represent physiology-based clusters of known T2D variants as classified in Mahajan et al., *Nat. Genet.* 2018. Novel variants are shown in black; all other variants are shown in gray. **c,d**, Tissue-of-action (TOA) profiles for fine-mapped GWAS signals in MCPS, based on active chromatin states in 223 adult biosamples from EpiMap **c,** or single-cell chromatin accessibility annotations in 218 adult cell types from the *Cis* Element Atlas (CATlas) **d**. TOA scores reflect the tissue specificity of each signal, based on the weighted partitioning of posterior probabilities from credible variants mapped to functional annotations. Results are collapsed into tissue groups and limited to tissues where ≥1 signal had TOA ≥ 0.25, and to signals with TOA ≥ 0.25 in at least one tissue.

**Extended Data Fig. 8.**
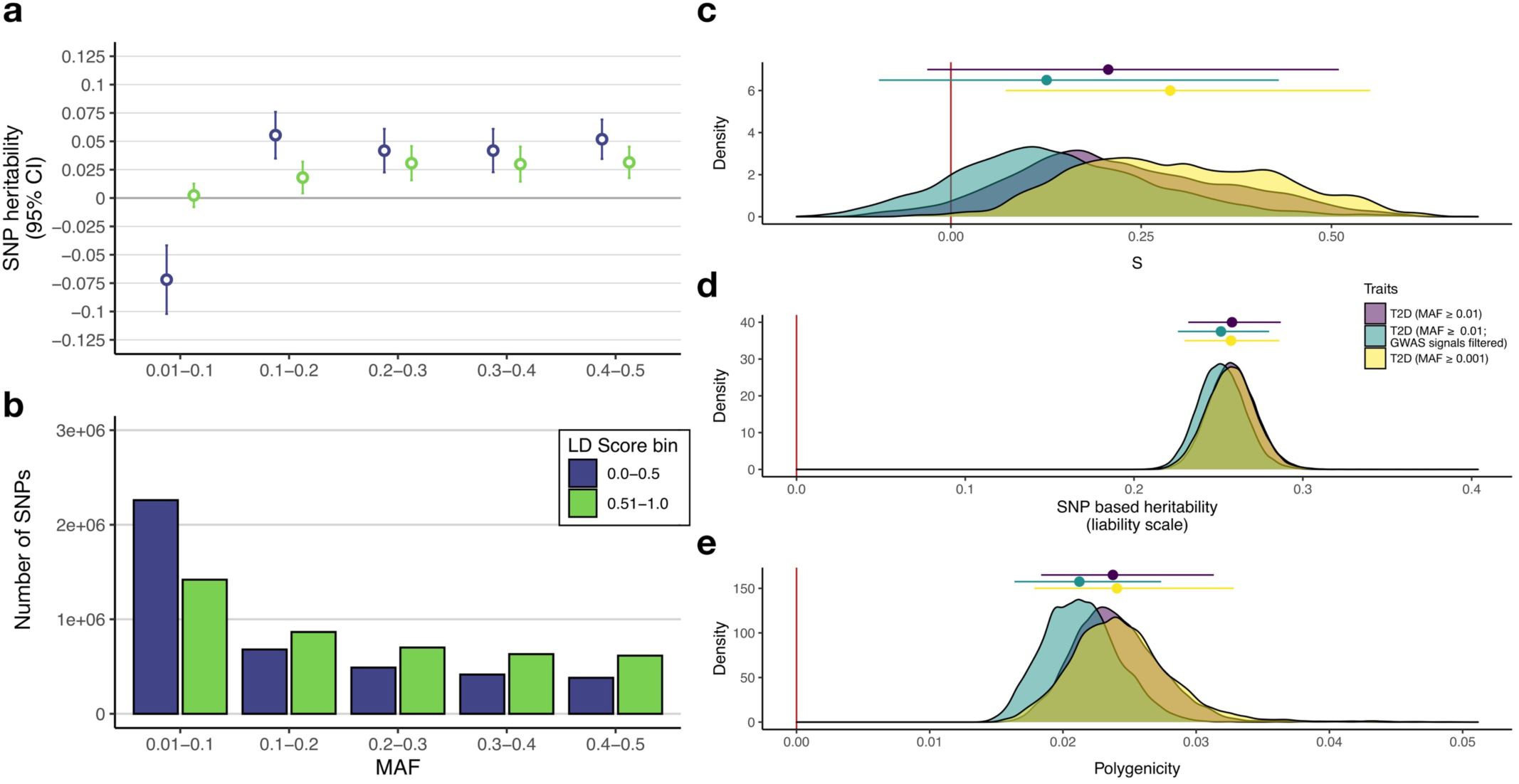
Heritability and genetic architecture of T2D in MCPS. **a**, SNP heritability 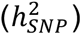 partitioned by minor allele frequency (MAF) and linkage disequilibrium (LD) score using GREML-LDMS applied to TOPMed-imputed variants. Estimates are shown on the liability scale for 56,248 participants unrelated up to the fourth degree (8,223 cases and 48,025 controls). Variants were stratified into five MAF bins and two LD-score bins and jointly fitted in a multicomponent model. Points and error bars indicate heritability estimates and 95% confidence intervals for each component. **b**, Number of variants assigned to each MAF–LD bin used in the GREML-LDMS analysis. **c–e**, Posterior distributions of genetic architecture parameters estimated using GCTB-BayesS. Points indicate posterior medians and horizontal lines indicate 95% credible intervals. **c**, S parameter, which captures the relationship between effect size and MAF. **d**, SNP heritability 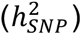. **e**, Polygenicity (*π*), defined as the proportion of SNPs with non-zero effects. Colours indicate alternative analysis specifications: purple, primary T2D analysis restricted to variants with MAF ≥ 0.01; yellow, sensitivity analysis including variants with MAF ≥ 0.001; green, sensitivity analysis excluding GWAS index variants and variants in LD (*r^2^* > 0.2) with them. All analyses were performed in the set of 56,248 participants unrelated up to the fourth degree and adjusted for age, sex and the first seven genetic principal components.

**Extended Data Fig. 9.**
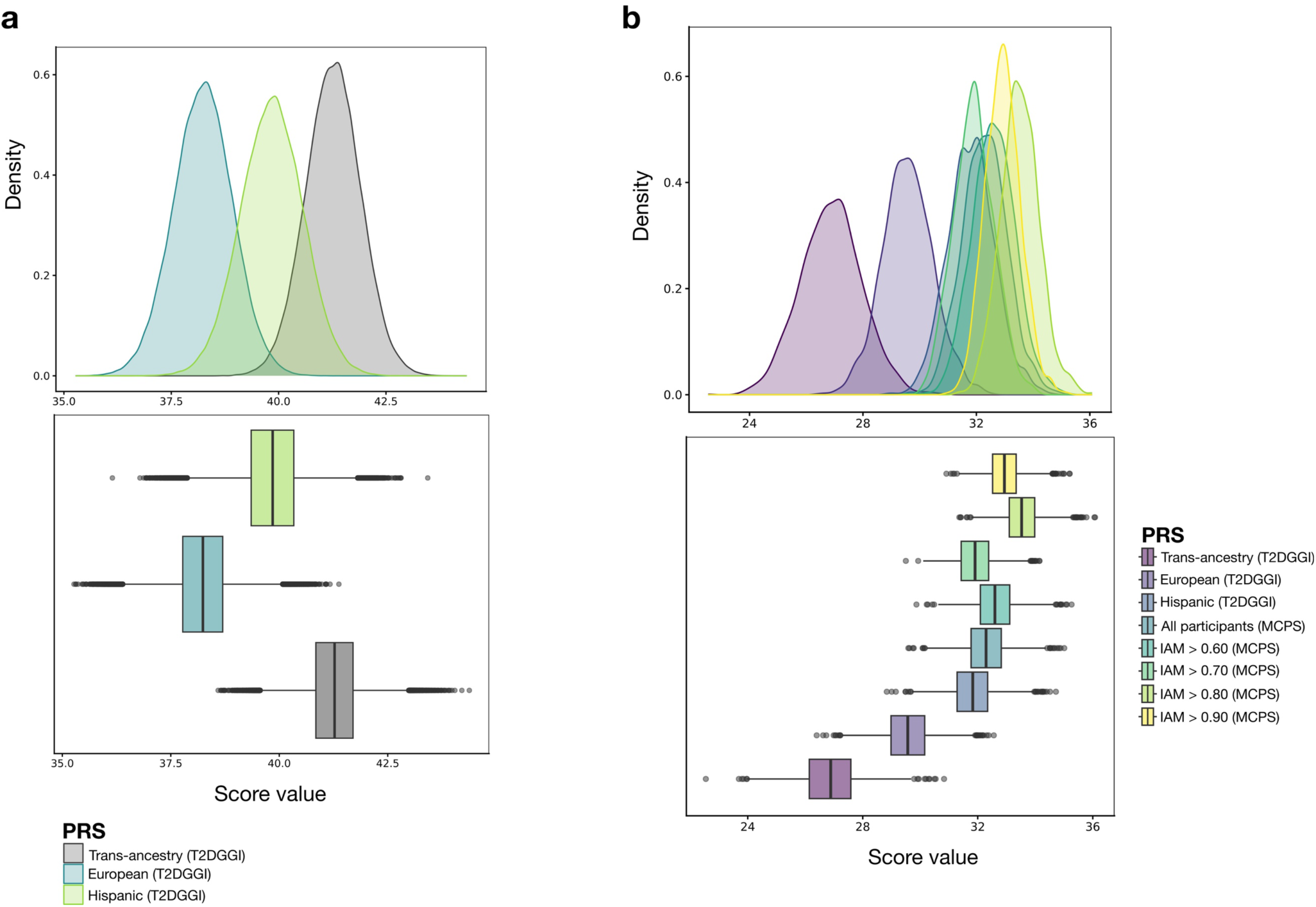
Distributions of T2D polygenic risk scores in MCPS and MAIS. **a**, Distributions of trans-ancestry (TA), European (EUR) and Hispanic (HIS) T2D polygenic risk scores (PRSs) among 138,025 MCPS participants with complete covariate data. PRSs were calculated using conditionally independent variants and effect estimates from Suzuki et al.3. A total of 1,055 of 1,289 variants (82%) were matched in MCPS. **b**, Distributions of T2D PRSs among 1,953 participants from the Metabolic Analysis in an Indigenous Sample (MAIS) study. In addition to TA, EUR and HIS weights from T2DGGI, PRSs were generated using effect estimates derived from all MCPS participants and from subsets of MCPS participants with progressively higher Indigenous American ancestry proportions (>60%, >70%, >80% and >90%). Between 1,059 and 1,061 variants (∼82%) were matched in MAIS.

**Extended Data Fig. 10.**
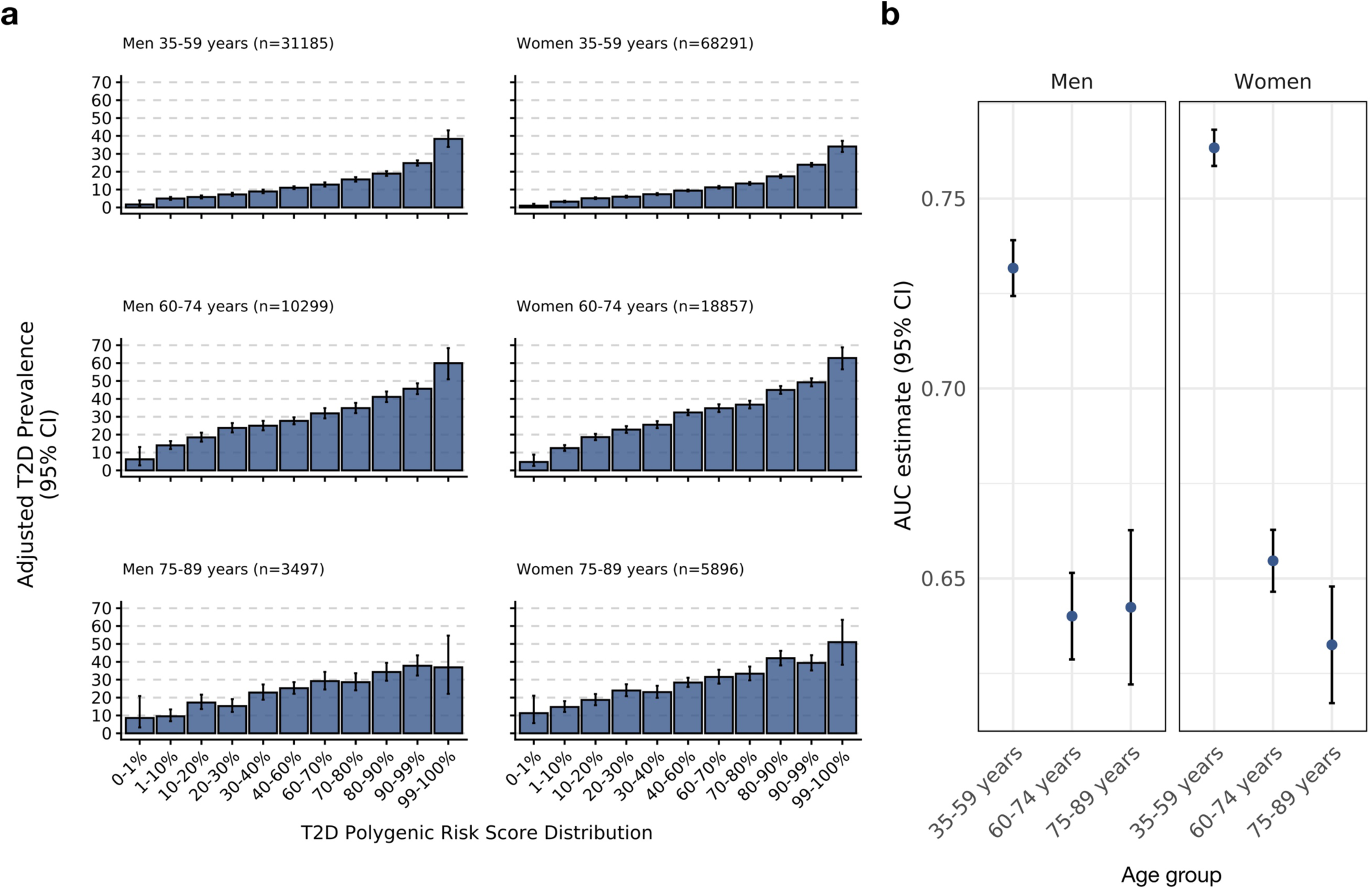
Association between a trans-ancestry T2D polygenic risk score (PRS) and T2D in MCPS, stratified by age and sex. **a**, Adjusted prevalence of T2D across increasing levels of the trans-ancestry PRS derived from T2DGGI. Participants were grouped according to the PRS distribution among controls. Prevalence estimates were adjusted for age, district of residence and body-mass index within each age- and sex-specific stratum. **b**, Predictive performance of the trans-ancestry PRS across age- and sex-specific groups. Area under the receiver operating characteristic curve (AUC) estimates were obtained from logistic regression models adjusted for age, district of residence and body-mass index within each stratum, with the PRS modelled as a continuous variable (per one standard deviation increase).

## Notes

### Competing Interest Statement

Jonathan R. Emberson and Rory Collins report grants to the University of Oxford from AstraZeneca and Regeneron Pharmaceuticals. Rory Collins reports having a patent for a statin-related myopathy genetic test licensed to the University of Oxford from Boston Heart Diagnostics (Rory Collins has waived any personal reward with any share in royalty and other payments waived in favour of the Nuffield Department of Population Health, University of Oxford) and being chair of not-for-profit clinical trial company PROTAS, chief executive of UK Biobank, and chair of the steering committee of the ORION-4 clinical trial of inclisiran. Alan R. Shuldiner is an employee of Regeneron Pharmaceuticals, Inc. and receives salary, stock and stock options as compensation for his employment. Eleanor Wheeler is a current employee and stockholder of AstraZeneca. All other authors declare no competing interests.

### Author Declarations

The Mexico City Prospective Study (MCPS) was approved by the Mexican Ministry of Health, the Mexican National Council of Science and Technology (0595 P-M), the Central Oxford Research Ethics Committee (C99.260), and the Ethics and Research Commissions of the Faculty of Medicine at the National Autonomous University of Mexico (FMED/CI/SPLR/067/2015). Transport and long-term storage of blood samples (plasma and buffy coat) at the Clinical Trial Service Unit sample archive, University of Oxford, were approved by the Mexican Ministry of Health. All participants provided written informed consent.

