## Supplementary Information for "Type 2 diabetes genetics in 125,000 admixed adults from Mexico City"

### Supplementary Results

#### Adiposity-adjusted GWAS

Mexico has among the highest rates of overweight and obesity globally. To assess the impact of adiposity in the GWAS, we performed genetic analyses adjusted for body mass index (BMI) and waist-to-hip ratio (WHR). Overall, adjusting for adiposity did not substantially alter the strength of association for most GWAS signals (Supplementary Fig. 12). However, significant differences in effect sizes (FDR < 0.05) were observed for three signals in the BMI-adjusted analysis and three in the WHR-adjusted analysis. Adjustment for BMI significantly increased the strength of association for rs118012224 at *HMGA1* and the lead signal rs2237897 at *KCNQ1*. In contrast, BMI adjustment significantly attenuated the association of the missense variant rs79783591 in *MC4R* (unadjusted $Z = 6.38$; adjusted $Z = 5.68$). Although BMI adjustment also weakened the associations at two other T2D-associated variants known to act via adiposity—rs71336381 in *MC4R* and rs1558902 in *FTO*—these differences were smaller and not statistically significant, and less pronounced than those observed in the DIAGRAM study[1]. While BMI is a well-established risk factor for T2D, this finding is consistent with previous MCPS results showing little difference in BMI between participants with and without a diabetes diagnosis, likely due to weight loss from poorly controlled disease prior to recruitment[2]. WHR adjustment increased the strength of association for signals at *PPARG* (rs6802898), *ANKRD55* (rs405176), and *SLC16A11* (rs113748381). No signals showed significantly weaker associations with WHR adjustment.

#### Additional analysis of rs139343836 at *RFX6*

We further characterised the rs139343836 (*RFX6*) association by evaluating alternative genetic models, ancestry effects, environmental interactions and epistatic interactions. Although the variant was associated with T2D under dominant and recessive models, tests assuming an additive effect (allelic and trend) yielded the lowest *p*-values, consistent with an additive effect on T2D risk (Supplementary Table 24). We next examined whether the effect of rs139343836 varied according to Indigenous American (IAM) ancestry proportion. Although the variant remained positively associated with T2D risk across the ancestry spectrum, effect sizes attenuated with increasing IAM ancestry proportion (Supplementary Fig. 4b). This pattern was supported by a significant SNP-by-ancestry interaction ($OR_{interaction} = 0.63$, 95% CI [0.47, 0.85]; $p = 0.0023$; Supplementary Fig. 4d), indicating that a shift from 0% to 100% IAM ancestry reduced the SNP's odds ratio by approximately 37%. No significant interactions were observed with IAM local ancestry dosage, BMI, physical activity, or sugary drink consumption. Additionally, no genome-wide epistatic interactions passed the FDR < 0.05 threshold.

The missense variant rs139343836 (c.985G>A) variant results in a valine to isoleucine substitution at position 329 of RFX6, a transcription factor involved in pancreatic islet development and function[3,4]. We performed a set of *in silico* analyses and found that the valine 329 residue is highly conserved (Supplementary Fig. 5a–b) and maps to the C domain which, along with two other domains (B and Dimerization), is crucial in RFX6 dimer formation[5]. Protein modelling predicted a slight destabilising effect for p.Val329Ile ($\Delta\Delta G \approx -0.24$ kcal·mol$^{-1}$); however, given the small magnitude and the uncertainty inherent to homology modelling, we refrain from drawing strong mechanistic conclusions (Supplementary Fig. 5c-e).

**Physiological clustering**

To contextualize biological mechanisms, we performed a "physiological clustering" analysis[6,7] that integrated MCPS PheWAS results with previously classified variants based on glycemic, anthropometric and lipid trait associations in external studies (Supplementary Tables 13–14). Two signals—at *PPM1G* (previously classified) and *SLC16A11* (classified here)—were clustered with variants associated with impaired lipid metabolism, including signals at *GCKR*, *TM6SF2*, and *TOMM40-APOE*[6]. These signals showed predominantly negative metabolite associations, including reductions in VLDL particle lipid features (free cholesterol, esterified cholesterol, phospholipids) and triglycerides, together with modest positive associations with docosahexaenoic acid and pyruvate (Extended Data Fig. 6).

Five variants at *ADCY5*, *ABO*, *KCNQ1* (rs2237897), *HMGA1*, and *ANKRD55* (rs5868014) showed predominantly positive associations with circulating lipid traits, with more than seven-fold as many positive as negative NMR trait associations. Together, these variants shared positive associations with VLDL particle features, circulating glucose, total triglycerides, and triglycerides across LDL, IDL, and HDL particles (Extended Data Fig. 6, Supplementary Table 12). These variants also overlapped with signals associated with lower fasting insulin and HOMA-B, and with altered proinsulin levels[6].

The *KLF14* signal was previously classified as impaired insulin action, characterised by lower BMI, adiponectin and HDL cholesterol, and higher proinsulin and TG levels. In our analysis, this variant showed a similar number of positive (26) and negative (24) associations. Positive associations included total triglycerides and triglycerides across VLDL, LDL, IDL, and HDL particles, as well as free cholesterol, phospholipids, and esterified cholesterol in VLDL particles; negative associations included ApoA1, LDL cholesterol, LDL size, and free cholesterol and esterified cholesterol in HDL particles. The *KLF14* risk allele was also negatively associated with adiposity traits and was the only T2D signal associated with increased BMI-adjusted waist-to-hip ratio in this cohort ($\beta = 0.013$, 95% CI 0.003–0.023, $p = 5.0\times10^{-3}$).

**Elucidating tissue-of-action scores with TACTICAL**

We integrated fine-mapped credible sets with chromatin state maps from 223 adult tissue and primary-cell biosamples from EpiMap[8], and 218 single-cell chromatin accessibility annotations from CATlas[9], including 107 brain-specific maps[10], and used TACTICAL[11] to estimate tissue-of-action (TOA) scores for each GWAS signal. TOA scores recapitulated established biology at known T2D loci. The obesity-associated signal at *FTO* had the highest TOA scores in EpiMap brain tissues (0.19) and CATlas brain GABAergic neurons (0.17). TOA scores also supported expected tissues for several known T2D risk variants: endocrine pancreas for signals at *SLC30A8*, *GLIS3*, *CDKN2B*, and *DGKB*; adipose tissue for *PPARG*; liver for *PPM1G* and skeletal muscle for *ANK1* (Supplementary Table 16).

TOA scores also provided insight into novel GWAS signals. The rare missense variant at *MC4R* (rs79783591) showed strong scores for brain tissue (0.69, EpiMap) and brain cells (e.g., glutamatergic neurons: 0.26, GABAergic neurons: 0.17) (Extended Data Fig. 7c–d). The highest scores for the *OPTC* signal (rs6685593) were observed in adipose tissue (0.22, EpiMap) and adipocytes (0.33, CATlas). The signal at *RFX6* (rs139343836) was most enriched in endocrine pancreas (0.67, EpiMap), while the signal at *NR5A2* (rs140064850) showed a high score in pancreas overall (0.60, EpiMap) but was most enriched in pancreatic ductal cells (0.26, CATlas). For *HNF1A* (rs483353044, PPA = 0.96), the highest scores were in non-endocrine acinar cells of the pancreas and stomach chief cells (both 0.48, CATlas) (Extended Data Fig. 7c–d, Supplementary Table 16).

### Supplementary Methods

**Cohort description**
The Mexico City Prospective Study (MCPS) was established by epidemiologists at the National Autonomous University of Mexico (Universidad Nacional Autónoma de México; UNAM) in collaboration with epidemiologists at the University of Oxford with the aim of identifying major risk factors for disease and premature death in Mexico. As described previously[2,12], 159,755 adults aged ≥ 35 years were recruited from 106,059 households located in two districts of Mexico City (Coyoacán and Iztapalapa) between 1998 and 2004. A questionnaire was completed by each participant that included age, sex, socioeconomic factors, lifestyle factors (e.g. smoking status, alcohol consumption, and physical activity), medication usage, and medical history. Height, weight, and hip and waist circumferences were measured using standard protocols. Systolic and diastolic blood pressure were recorded with participants in a seated position. Deaths of study participants are tracked through electronic linkage to a national death registry[2]. A 10-ml blood sample was obtained and used for extraction of plasma and buffy coat samples from each participant[12]. HbA1c was measured by the University of Oxford Clinical Trial Service Unit's Wolfson laboratories using buffy coat samples and a high-performance liquid chromatography method on HA-8180 analysers as previously described[13]. The laboratory is ISO17025-accredited as a testing laboratory and the method is on their scope of accreditation. Metabolomic profiling on the Nightingale Health Ltd targeted NMR platform was performed using the same validated high-throughput protocol across all samples, including those newly analysed for this study[14].

**Independent vertex set analysis of relatedness networks**
Pairwise relatedness inference based on shared identical-by-descent (IBD) segments was performed using KING v2.2.7 with the --ibdseg option on the full set of 140,829 genotyped participants. KING determines all segments with one or two copies shared IBD between individuals (i.e., IBD1 and IBD2), from which relatedness can be inferred. The --degree 3 option was used to report all pairs of inferred relatives within the third degree (i.e., IBD proportion > 0.0884).

To obtain a maximum set of unrelated individuals, the 40,695 participants without any third-degree relatives in the cohort were retained. Independent vertex set (IVS) analysis was then performed on the remaining set of 100,134 participants. Clusters were delineated using the *clusters* function within the *igraph* package in R v3.6.2 with edges corresponding to relationship pairs inferred from IBD segment analysis. For each cluster with ≤50 participants, a single largest maximum IVS set was selected from the output of the *largest_ivs* function in the R *igraph* package. For clusters with >50 participants, a greedy algorithm[15] was applied wherein the vertex (i.e., participant) with the smallest degree was selected with removal of neighbors from consideration. This procedure was iterated until no vertices from the cluster remained. The combination of the resulting set of IVS-selected individuals with the set of 40,695 unrelated individuals yielded a set of 79,612 maximally unrelated individuals up to the third degree, heretofore referred as the *maximum unrelated set*.

**Admixture**

WGS variant calls from globally diverse samples in the 1000 Genomes Project and the Human Genome Diversity Project (HGDP) were previously merged[16] with TOPMed-imputed genotypes of Indigenous Mexican samples from the Metabolic Analysis of an Indigenous Sample (MAIS) study[17]. This dataset was then merged with TOPMed-imputed genotypes from MCPS participants at variants with imputation INFO scores ≥ 0.99 using the procedure described by Ziyatdinov, Torres, and Alegre-Díaz et al. 2023[16]. This produced a merged dataset of 459,622 autosomal variants. PLINK v1.9 was then used to perform LD clumping using allele frequencies as pseudo *p*-values and the options --clump-p1 1, --clump-p2 1, and --clump-r2 0.95, resulting in a clumped dataset of 132,891 autosomal variants. From this dataset, a subset of reference samples representing five global "superpopulations" were selected: Africa (n = 765), East Asia (n = 727), Europe (n = 658), Middle East (n = 161), and America (n = 814). The America samples included 64 Mexican Americans from Los Angeles (MXL) from 1KG, and 21 Maya and 13 Pima samples from HGDP. 1,000 random MCPS participants unrelated up to the fourth degree were also selected, resulting in a total of 1,814 American samples (and an overall total of 4,125 reference samples).

Ancestry-specific allele frequencies and per-individual ancestry proportions were estimated with ADMIXTURE v1.3.0. An admixture model was fit for each *K* parameter (i.e. the number of inferred ancestral populations) in the set $K = \{4,...,40\}$ and the --cv option was specified in each analysis to perform a five-fold cross validation procedure. The *K* parameter that yielded the lowest cross-validated error was $K = 20$. Learned clusters and ancestry proportions from the $K = 4$ and $K = 20$ models estimated with the *unsupervised* mode of ADMIXTURE were then used to project the remaining set of MCPS participants and estimate ancestry proportions with the -P option. Each of the estimated *K* ancestries was assigned to a global "superpopulation" and the cumulative *K* ancestry proportion for each superpopulation was used to estimate total ancestry in each individual from that population.

**Imputation workflow**

Prior to imputation, the quality-controlled genotyping array dataset was converted from PLINK format to unphased VCF files using PLINK 2.0, as described previously[16]. To accommodate TOPMed Imputation Server constraints, samples were randomly assigned to six tranches of approximately 23,500 individuals and split into per-chromosome bgzipped VCF files. Imputation jobs were submitted via the NIH BioData Catalyst API of the TOPMed Imputation Server.

Following imputation, dosage VCF files were merged across all samples in 5-Mb genomic regions using bcftools. Imputed variants were then converted into three separate genotype representations for downstream analyses: (1) BGEN files generated from genotype probabilities (GP) using QCTOOL v2; (2) PGEN files generated from haploid dosages (HDS) using PLINK 2.0; and (3) PGEN files generated from hard-called genotypes (GT) using PLINK 2.0.

**GWAS relatedness adjustment**

Genetic association analyses were performed using SUGEN[18,19], which implements a generalized estimating equation framework for genetic association testing in related samples. The model assumes that phenotypes are correlated within but not between families. Family networks were defined using pairwise IBD estimates calculated from array genotype data. Following recommendations from the SUGEN framework[19], networks were constructed by connecting individuals sharing first-degree relationships. This procedure resolved 23,213 family networks (median size = 2; maximum size = 48), which were supplied to SUGEN to account for within-family correlation.

**Adiposity-adjusted GWAS**

GWAS analyses were performed with SUGEN as described above (see *Genome-wide association studies* in Methods) with additional adjustment for BMI or WHR as covariates in the logistic regression model. After excluding individuals with missing adiposity measures, 18,957 cases and 104,583 controls were retained in the BMI-adjusted analysis and 18,956 cases and 104,593 controls were retained in the WHR-adjusted analysis. Stepwise-approximate conditional analysis was then performed for each trait as described above (see *Dissection of independent GWAS signals* in Methods). The impact of adiposity adjustment was assessed by comparing marginal effect size estimates between the unadjusted and adjusted analysis for the set of index variants corresponding to conditionally-independent association signals. The significance of difference between beta coefficients for the unadjusted and adjusted analyses for each variant was tested using the *t* statistic:

$$t = \frac{\beta_{unadj.} - \beta_{adj.}}{\sqrt{SE_{unadj.}^2 + SE_{adj.}^2 - 2rSE_{unadj.} \cdot SE_{adj.}}}$$

where *r* corresponds to the correlation between the unadjusted and adiposity-adjusted beta coefficients, calculated across all genome-wide variants with the Spearman rank correlation coefficient (estimated to be 0.982 and 0.975 for BMI and WHR, respectively). *p* values derived from this statistic ($P_{diff}$) were adjusted using the Benjamini-Hochberg procedure to control the false discovery rate.

**Covariate-adjusted LD score estimation**

Covariate-adjusted LD (cov-LD) scores were estimated from a previously quality controlled set of 9,764 whole-genome sequenced MCPS participants[16] unrelated at a KING kinship coefficient threshold of 0.0442[20]. Principal components (PCs) were calculated using the bigsnpr workflow[21] applied to variants with MAF ≥ 0.05 and an LD-clumping threshold of $r^2 = 0.005$, resulting in 24,217 retained variants. To further reduce the influence of relatedness, 266 individuals with inferred relationships within the third degree based on IBD segment sharing were excluded prior to PCA and subsequently projected into the resulting PC space. No outlier individuals were detected. As required by cov-LDSC[22], PCs were centred to mean zero and standard deviation 0.01. Eight PCs were selected for covariate adjustment based on inspection of loading plots and PC by PC plots, which indicated population structure rather than local LD structure[21] (Supplementary Fig. 13). Cov-LD scores were calculated for 18,789,983 variants with MAF ≥ 0.1% using 1 Mb windows of 1 Mb, consistent with previous large-scale analyses (e.g., Pan-UK Biobank study).

**Approximate conditional analysis**

Approximate conditional analyses were performed using GCTA-COJO (v1.94). Linkage disequilibrium was estimated from TOPMed-imputed genotypes corresponding to the 79,162 MCPS participants in the maximum unrelated set. The --cojo-collinear threshold was set to 0.9. At each locus, forward selection was performed using the --cojo-cond function, starting with the index variant and continuing until no additional variant attained a conditional $p < 5 \times 10^{-8}$. For loci with multiple selected variants, iterative backward elimination and forward selection were subsequently performed until no additional variants met the significance threshold and all retained variants remained genome-wide significant in the joint model estimated with the --cojo-joint function.

**Identification of novel GWAS signals**

Reference variant identifiers corresponded to dbSNP version 153. Previously reported T2D-associated variants were obtained from the NHGRI-EBI Catalog (version 1.0.2; accessed August 21, 2022). Phenotype designations used for extracting reported T2D variants included: “Type 2 diabetes”; “Type 2 diabetes (adjusted for BMI)”; “Type ii diabetes”; “Childhood onset type 2 diabetes”; “Type 2 diabetes (age of onset)”; “Severe insulin-deficient type 2 diabetes”; “Severe insulin-resistant type 2 diabetes”; “Mild obesity-related type 2 diabetes”; “Mild age-related type 2 diabetes”; “Severe autoimmune type 2 diabetes”. Index variants from the trans-ancestry meta-analysis of 1.3M individuals by DIAMANTE[23] were also included to ensure completeness.

In signals classified as occurring at known loci, all previously reported T2D-associated variants within 500 kb of the index variant were collated. Of 481 previously reported variants identified in these regions, 453 variants (94.2%) were present among imputed variants passing quality control thresholds and were retained for analysis. Pairwise LD ($r^2$) between index GWAS variants and previously reported variants was estimated using PLINK v2.0 within the maximum unrelated set of MCPS participants. To minimise collinearity during conditional analysis, previously reported variants were LD-pruned at $r^2$ thresholds of 0.9 and 0.5 (using arguments --r2, --ld-window-r2 0, --window 100, and --ld-window-kb 2000), yielding reference sets of 404 and 282 variants, respectively. Approximate conditional analyses were performed in GCTA-COJO with the maximum unrelated set as the LD reference panel (using arguments --cojo-cond and --cojo-collinear). Signals were first conditioned on variants from the $r^2 < 0.9$ reference set. Where analyses failed because of excessive collinearity, conditioning was repeated using the $r^2 < 0.5$ reference set. Signals whose index variant remained genome-wide significant after conditioning ($p < 5\times10^{-8}$) were considered novel associations at known loci.

**Exome-wide association study**

Analyses of exome sequencing data from 141,046 MCPS participants were restricted to 9,325,897 variants that passed previous quality-control procedures involving machine-learning based detection of low-quality variants and hard filters[16]. Single-variant association test was performed using REGENIE v3.1.3. Quality-controlled genotyping array variants were further restricted to those with MAC ≥ 100 (560,015 variants in 140,829 individuals) and used to fit the whole-genome regression model (step 1) with a block size of 1000. Association testing was performed using a logistic regression score test with LOCO predictors from step 1. For step 2, a block size of 400 was used and analyses were restricted to exome variants with MAC ≥ 25. An approximate Firth correction was applied to variants with $p < 0.01$. Association tests were performed conditional on all genome-wide significant GWAS variants present in the exome variant set (249 overlapping variants) using the --condition-list argument. The logistic regression models was adjusted for age, sex, and the first seven genetic PCs as covariates. Significant variants were annotated using the Ensembl VEP web interface. Annotation outputs included MANE Select transcript consequences, population allele frequencies from the 1000 Genomes Project and gnomAD, and pathogenicity predictions from SIFT, PolyPhen, PrimateAI, REVEL and CADD.

**Gene-based rare variant testing**

Exome sequence variants were annotated per guidelines from the Biobank Rare Variant Analysis (BRaVa) Consortium guidelines, which integrate VEP predictions, CADD scores, and splice-site disruption predictions from SpliceAI. Annotations were restricted to canonical protein-coding transcripts and variants were classified into five non-overlapping categories:

(1) Putative loss-of-function (pLoF) variants with high-confidence Loftee calls (181,675 variants)
(2) Damaging missense or protein altering variants meeting one or more of the following criteria; REVEL ≥ 0.773, $CADD_{PHRED}$ ≥ 28.1, SpliceAI max Δ-score ≥ 0.20, or low-confidence Loftee calls (520,998 variants)
(3) Other missense or protein-altering variants (i.e., missense, start-loss, stop-loss, or in-frame indel variants) not in category 2 (2,118,452 variants)
(4) Synonymous variants with SpliceAI DS score < 0.2 (1,195,125 variants)
(5) Non-coding variants impacting UTRs, introns, or intergenic regions (6,897,946 variants)

Variants not assigned to any category were excluded from further analyses.

Gene-based analysis used either pLoF variants alone (category 1) or pLoF plus damaging missense/protein-altering variants (categories 1 and 2). Two maximum allele-frequency thresholds were applied (singletons and MAF < 0.1%), resulting in four variant masks per gene. Gene-based tests were performed in REGENIE v3.1.3 using the same genome-wide predictors and logistic regression covariates as the ExWAS. Associations were evaluated using the "Gene P" framework[24], which combines SBAT, SKAT-O, ACAT-V, and BURDENT-ACAT statistics using the Cauchy combination method. Genes with "Gene P" $p \leq 2.73\times10^{-6}$ were considered significant after Bonferroni correction for 18,262 protein-coding genes.

**Genetic fine-mapping**

Independent GWAS signals were fine-mapped using the approximate Bayesian framework described by Wakefield[25]. Signals were included if the index variant had MAF > 0.0025 and did not map to the MHC region. For loci containing multiple independent signals, conditional association statistics were obtained using the GCTA-COJO --cojo-cond procedure by conditioning on all remaining signals at the locus. Otherwise, marginal association statistics was used.

Approximate Bayes factors ($\Lambda_j$) were estimated for each variant *j* at a locus using:

$$\Lambda_j = \sqrt{\frac{V_j}{V_j + \omega}}\, exp\left[\frac{\omega\beta_j^2}{2V_j(V_j\omega)}\right]$$

Where $\beta_j$ is the estimated allelic effect on the log-odds ratio scale, $V_j$ is the corresponding variance estimate, and $\omega$ is the prior variance in allelic effects. Consistent with previous T2D fine-mapping in the DIAGRAM study, $\omega$ was set to 0.04[1,25].

For each variant *j* within the set of variants at a locus, a posterior probability of association (PPA) value was obtained by:

$$\pi_j = \frac{\Lambda_j}{\sum_k \Lambda_j}$$

Variants were ranked according to descending order of PPA and included until cumulative PPA reached 0.99, yielding a 99% credible set. Fine-mapping resolution was assessed using both credible-set size and the maximum PPA value.

To compare fine-mapping resolution between MCPS and DIAGRAM, 99% credible sets were downloaded from the DIAGRAM and lifted from hg19 to GRCh38 coordinates with liftOver. Pairwise LD between index variants from MCPS and

DIAGRAM was estimated within the maximum unrelated set of MCPS participants using PLINK v1 (with arguments --ld-snp-list, --r2, --ld-window-kb 1000, and --ld-window-r2). Signals whose index variants were correlated at LD $r^2 \geq 0.4$ were considered to represent the same underlying association signal, resulting in 40 shared signals between studies.

**Replication cohorts and meta-analysis procedures**

Genome-wide summary statistics from trans-ancestry (TA) meta-analyses and ancestry-specific meta-analyses performed by the Diabetes Meta-analysis of Trans-Ethnic association studies (DIAMANTE)[23] and Type 2 Diabetes Genomics Initiative (T2DGGI)[26] consortia were downloaded from the DIAGRAM Consortium website. GWAS summary statistics from a multi-ancestry analysis of T2D conducted by the Population Architecture using Genomics and Epidemiology (PAGE) study[18] were downloaded from the NHGRI-EBI GWAS Catalog. Summary statistics from a BMI-unadjusted GWAS meta-analysis of T2D in East Asian cohorts performed by the Asian Genetic Epidemiology Network (AGEN)[27] were obtained from the AGEN website.

Results from a pan-ancestry genetic analysis of the UK Biobank were obtained from the Pan-UK Biobank website. Downloaded pan-UK Biobank results included summary statistics from a sex-combined analysis for phecode 250.2 (T2D) that corresponded to a 'fixed-effect' inverse-variance-weighted meta-analysis across all available ancestry groups for the trait. Ancestry-specific results for EUR, AFR, Middle Eastern (ME), and Central or South Asian (CSA) ancestries were also extracted from the Pan-UK Biobank website.

Studies that provided summary statistics for novel GWAS variants which were obtained through collaboration included: a Hispanic/Latino(H/L)-specific meta-analysis by the DIAMANTE Consortium; Ancestry-specific GWAS analyses of EUR, AFR, EAS, and H/L ancestries in the Million Veterans Program (MVP) cohort[28,29]; a GWAS of T2D involving 821 case and 1,695 control individuals of Indigenous Mexican ancestry from the Metabolic Analysis of an Indigenous Sample (MAIS) study[30,31]; a GWAS analysis of T2D in the Mexico Biobank[32].

Summary statistics were harmonised across all studies with respect to the designation of effect and non-effect alleles in MCPS. Effect allele frequencies (EAF) were estimated in the DIAMANTE-TA dataset by calculating allele counts from the reported EAF and sample counts in each of the ancestry-specific DIAMANTE datasets (i.e. EAF $\times$ 2 $\times$ $N_{cases} + N_{controls}$) and dividing allele counts by the total number of alleles across the constituent cohorts. An important caveat in these EAF estimates is that although African ancestry cohorts were included in the DIAMANTE-TA meta-analysis, African ancestry-specific summary statistics are not currently available and hence African ancestry allele and sample counts were not represented in these estimates.

To obtain trans-ancestry effect size estimates in the MVP study, an inverse-variance-weighted (IVW) meta-analysis of the log odds ratios (ORs) was performed using results from each ancestry-specific GWAS. To assess per-variant evidence of significance across studies, IVW meta-analyses were performed and were limited to the largest available non-overlapping studies.

**Protein modeling analysis of RFX6**

The wild-type RFX6 amino acid sequence (UniProt accession Q8HWS3) was modelled using I-TASSER. Evolutionary conservation was assessed using ConSurf. The impact of the p.Val329Ile substitution on protein stability was evaluated using DynaMut, which estimated the predicted change in folding free energy (ΔΔG). MutationMapper was used to generate the gene schematic shown in Supplementary Fig. 5.

**Phenome-wide association study exclusions**

Traits included HbA1c, systolic blood pressure, 10 anthropometric traits, seven baseline disease outcomes, all-cause mortality and 110 NMR metabolic features.

Quantitative trait analysis excluded participants with self-reported chronic disease at baseline, undiagnosed diabetes (HbA1c ≥ 6.5%), missing phenotype data or implausible anthropometric measurements (height <120 or >200 cm, weight <35 or >250 kg, BMI <15 or >60 kg/$m^2$, waist circumference <60 or >180 cm, hip circumference <70 or >180 cm, and WHR <0.5 or >1.5). Sample sizes ranged from 107,296 participants for most anthropometric and glycaemic traits, to 61,910–81,257 participants for NMR metabolite features, and approximately 5,800 participants for body fat percentage and visceral fat index measurements collected during follow-up resurveys. Analyses were restricted to directly measured NMR metabolite features, excluding metabolite ratios.

**Physiological clustering**

Of the 86 index variants from the GWAS of T2D in MCPS, 65 were either previously-reported or conditionally-dependent with respect to known T2D risk variants (Supplementary Table 1). These 65 variants were referenced against a set of T2D-associated variants that were previously assigned to distinct physiological clusters in a C-means "soft" clustering analysis[6]. Of these variants, 34 were linked to a physiologically-clustered variant either through direct matching (n = 6), through an LD proxy variant ($r^2 \geq 0.2$) (n = 21), or physical proximity (within 500 kb) (n = 7) (Supplementary Table 13). At least one variant was matched to each of the six clusters: Insulin secretion 1 (risk allele associated with lower fasting insulin [FI] and HOMA-B and higher proinsulin [PI]), Insulin secretion 2 (risk allele associated lower FI, HOMA-B, and PI, and higher HDL-C), Insulin secretion 2/Insulin action (risk allele associated with lower FI, HOMA-B, PI and HDL-C; or risk allele associated with lower BMI and HDL-C and higher PI), Insulin action (risk allele associated with lower HDL-C, BMI, and adiponectin and higher triglycerides [TG] and PI), Adiposity (risk allele associated with higher BMI and WHR), and Impaired lipid metabolism (risk allele associated with lower TG) (Supplementary Table 13).

Multidimensional scaling (MDS) was used to map index variants onto a two-dimensional space through decomposition a matrix of Z-scores for 86 variants from genetic association analyses performed in MCPS for 51 continuous traits (anthropometric, HbA1c, SBP and 39 NMR-based metabolite features pruned for collinearity). Variants identified in the GWAS of T2D in MCPS that had not been previously clustered by Mahajan et al.[6] were assigned to the pre-specified physiological clusters via a "hard" clustering based on a K-means algorithm using medoids and a "soft" probabilistic clustering based on a Gaussian mixture model using prespecified medoids and 1,000 iterations. The assigned mean cluster probabilities for each variant were estimated from the mean of 500 bootstrapped samples whereas 95% confidence intervals were derived from the 2.5$^{th}$ and 97.5$^{th}$ percentiles of the distribution of bootstrapped samples.

**Enrichment of functional annotations**

BED files of functional genomic annotations on build GRCh38 were accessed from the Common Metabolic Disease Genome Atlas (CMDGA). Annotations corresponding to DNAse hypersensitivity sites (DHS), assay for transposase-accessible chromatin using sequencing (ATAC-seq) peaks, single nuclear (snATAC-seq) peaks, and chromatin states were obtained from available bulk tissue and single cell samples. Genome-wide enrichment of functional annotations was assessed using GARFIELD[33] which implements a logistic model that regresses variant significance (i.e. a binary variable that indicates if a variant meets a specified significance threshold in the GWAS) on distance to nearest transcription start site (TSS), number of LD proxies, and annotation membership. LD tag variants for the requisite LD input file were identified from WGS data for 9,674 unrelated MCPS participants (using a KING kinship coefficient threshold of 0.0442) for variants with minor allele count (MAC) ≥10 using the PLINK1 command --show-tags all and specifying --tag-r2 0.8 and 0.01 for clumping and pruning tags, respectively. Requisite MAF input values were estimated from TOPMED imputed genotypes for the maximum set of unrelated individuals up to the third degree (n=79,123) using the --freq command in PLINK1. TSS annotations collated by Abugessaisa et al.[34] were downloaded and used to calculate per-variant distance to nearest TSS. Variants were annotated to each functional annotation using the *countOverlaps()* function in the R package GenomicRanges and converting the resulting Boolean output to integers. GARIELD analysis was restricted to bi-allelic variants.

**Functional integration at T2D loci**

Tissue-of-action (TOA) scores were estimated using TACTICAL, which integrates posterior probabilities of association (PPAs) from genetic credible sets with epigenomic annotations. Chromatin state maps corresponding to the 18-state Roadmap model were obtained from EpiMap for 223 adult biosamples representing 28 tissues. Chromatin states were derived from ChIP-seq profiles of six histone modifications (H3K27ac, H3K4me1, H3K4me3, H3K36me3, H3K9me3 and H3K27me3). Ten chromatin states were used for TOA estimation and weighted according to regulatory activity. Strong enhancer and promoter states (TssA, TssFlnkU, EnhG2 and EnhA1) were assigned a weight of 3, EnhA2 was assigned a weight of 2, and weaker enhancer/promoter states (TssFlnk, TssFlnkD, EnhG1 and EnhWk) together with transcribed regions (Tx) were assigned a weight of 1.

Single-nucleus ATAC-seq annotations were obtained from the *Cis* Element Atlas and comprised 218 adult cell-type maps spanning 26 tissues, including 107 brain cell types. Peaks were ranked by chromatin accessibility and assigned decile-based weights ranging from 1 to 10. To capture nearby regulatory variants, peak coordinates were extended by 500 bp.

For each GWAS signal, PPAs from the corresponding 99% credible set were partitioned across weighted chromatin-state and chromatin-accessibility annotations to generate TOA scores. Analyses were performed at the level of both tissues and individual biosamples or cell types. An "unclassified" score corresponded to the cumulative PPA of credible variants that did not overlap any epigenomic annotation.

**Heritability analyses**

SNP heritability attributable to variants genotyped on the GSAv2 array was estimated using single-component genetic restricted maximum likelihood (REML-SC) as implemented in GCTA v1.94.0. Estimates were obtained in three sets of MCPS participants with decreasing levels of relatedness: 1) all 125,042 GWAS participants; 2) 70,789 participants unrelated up to the third degree (10,796 cases and 59,993 controls); 3) 56,248 participants unrelated individuals up to the fourth degree (8,223 cases and 48,025 controls). For each set, a genetic relatedness matrix (GRM) was calculated from 539,448 autosomal GSAv2 variants passing quality control, and models included age, sex, and the first seven genetic principal components (and fit using the --reml function).

To obtain estimates robust to recent admixture, the analyses was repeated using admixture-adjusted GRMs with REAP[35]. Per-individual ancestry proportions and the 132,891 SNPs used to estimate them (see *Admixture*) were supplied to REAP. For computational tractability, genotypes were divided into 2,000-SNP chunks, from which 67 kinship matrices were generated and combined into a genome-wide GRM compatible with GCTA using the R package *plinkFile.* REML-SC was then performed in the subset of 56,248 participants unrelated to the fourth degree, using the same covariates as above.

Multicomponent SNP heritability was estimated using GREML-LDMS[36] applied to 8,455,702 TOPMed-imputed variants with MAF $\geq$ 0.01. Variants were partitioned into five MAF bins (0.5–0.4, 0.4–0.3, 0.3–0.2, 0.2–0.1, and 0.1–0.01) and two LD-score bins, and GRMs were jointly fitted in the 56,248 participants unrelated up to the fourth degree with age, sex, and the first seven genetic PCs as covariates. A disease prevalence of 0.12 was assumed for liability-scale conversion of all T2D heritability estimates, based on epidemiological data[37].

### Supplementary Figures

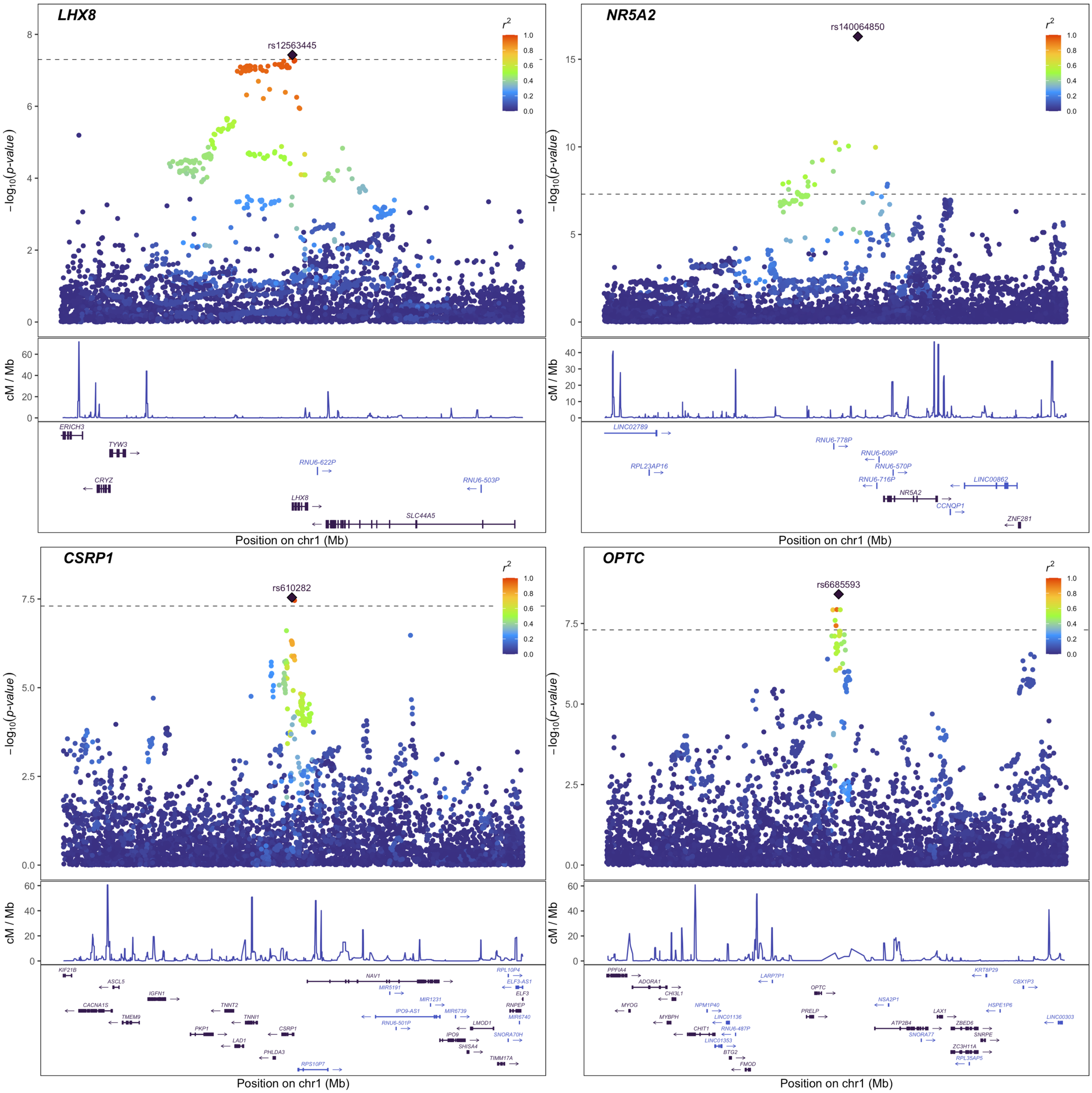


**Supplementary Fig. 1 | Locus plots for the 86 independent GWAS signals associated with T2D.** Negative log10($p$) values from approximate conditional analyses are shown for each signal, conditioning on all other independent signals within the locus. Index variants are indicated by diamonds, and colours correspond to pairwise LD ($r^2$) with the index variant, estimated in the maximum unrelated set of MCPS participants. Genomic positions are shown on the GRCh38 reference genome and gene annotations correspond to Ensembl release 105.

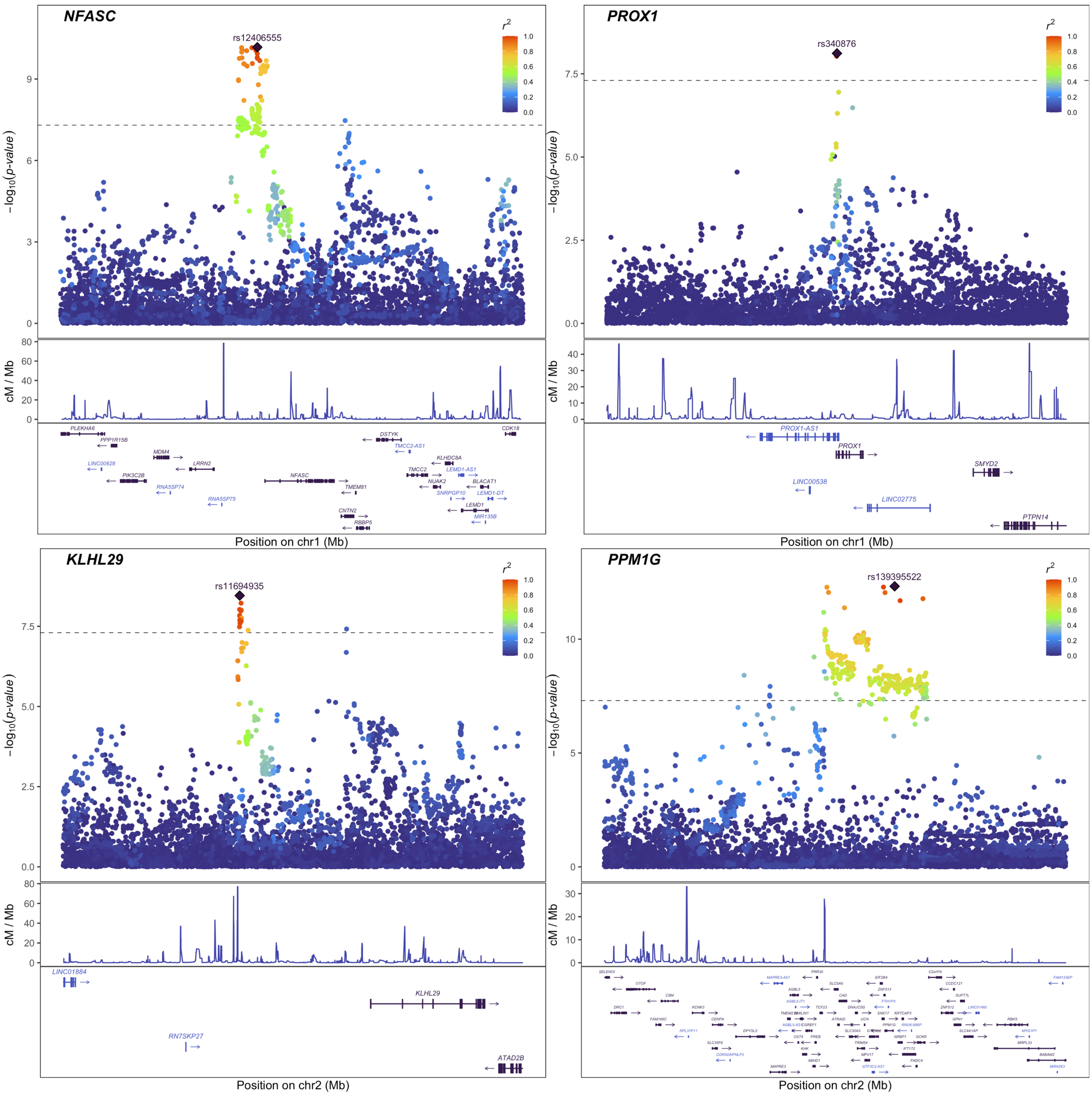


**Supplementary Fig. 1 (continued).**

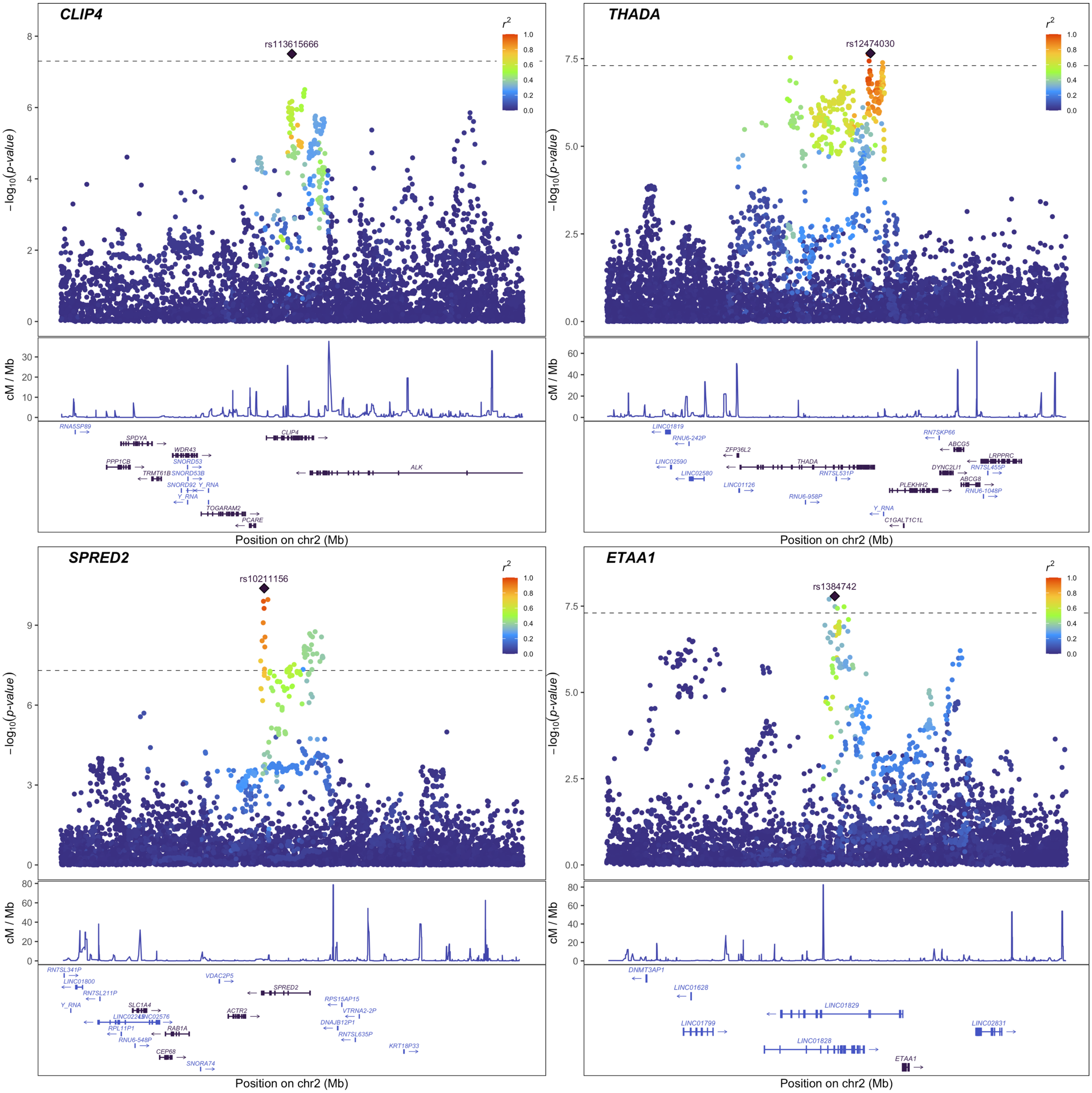


**Supplementary Fig. 1 (continued).**

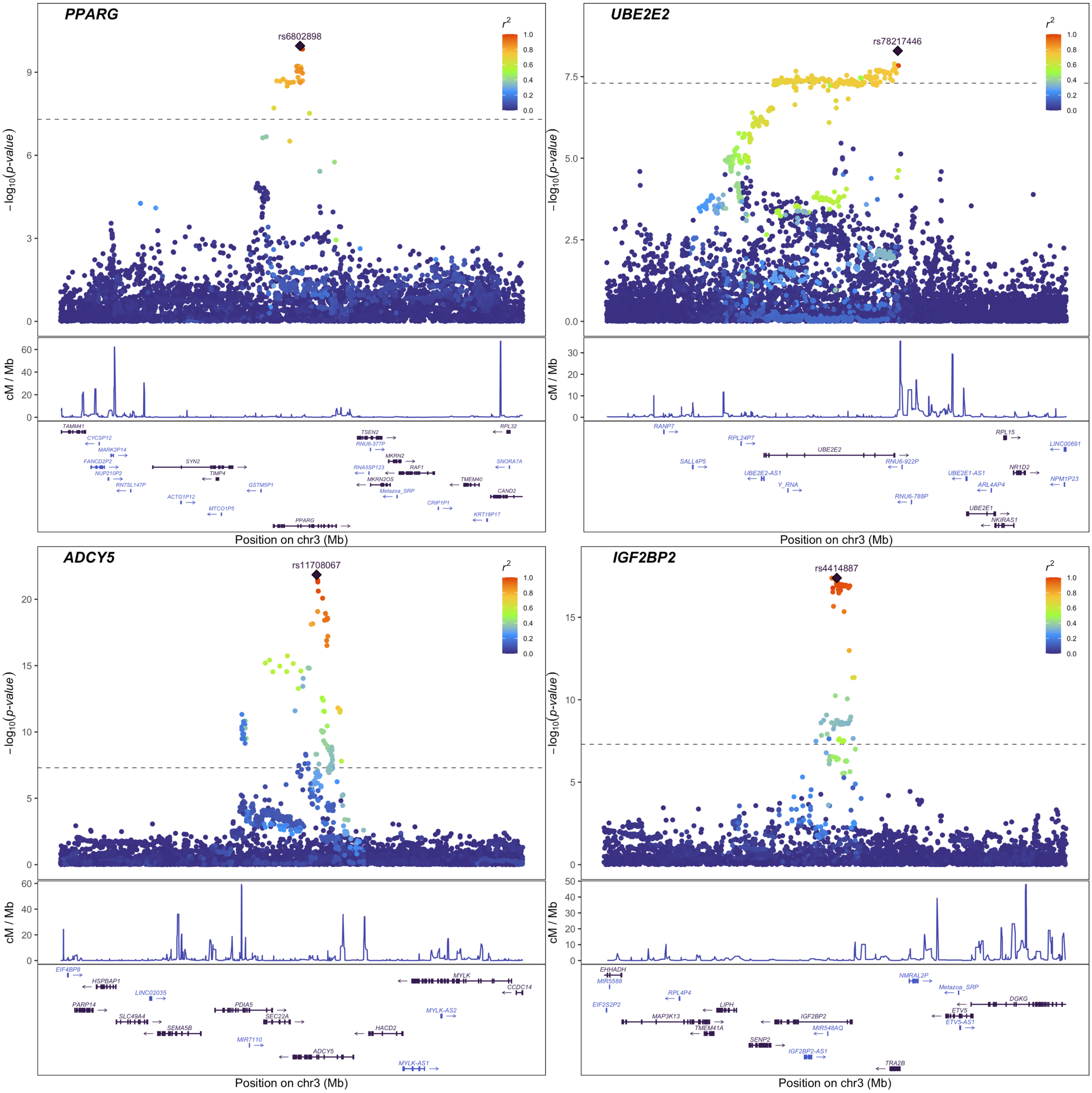


**Supplementary Fig. 1 (continued).**

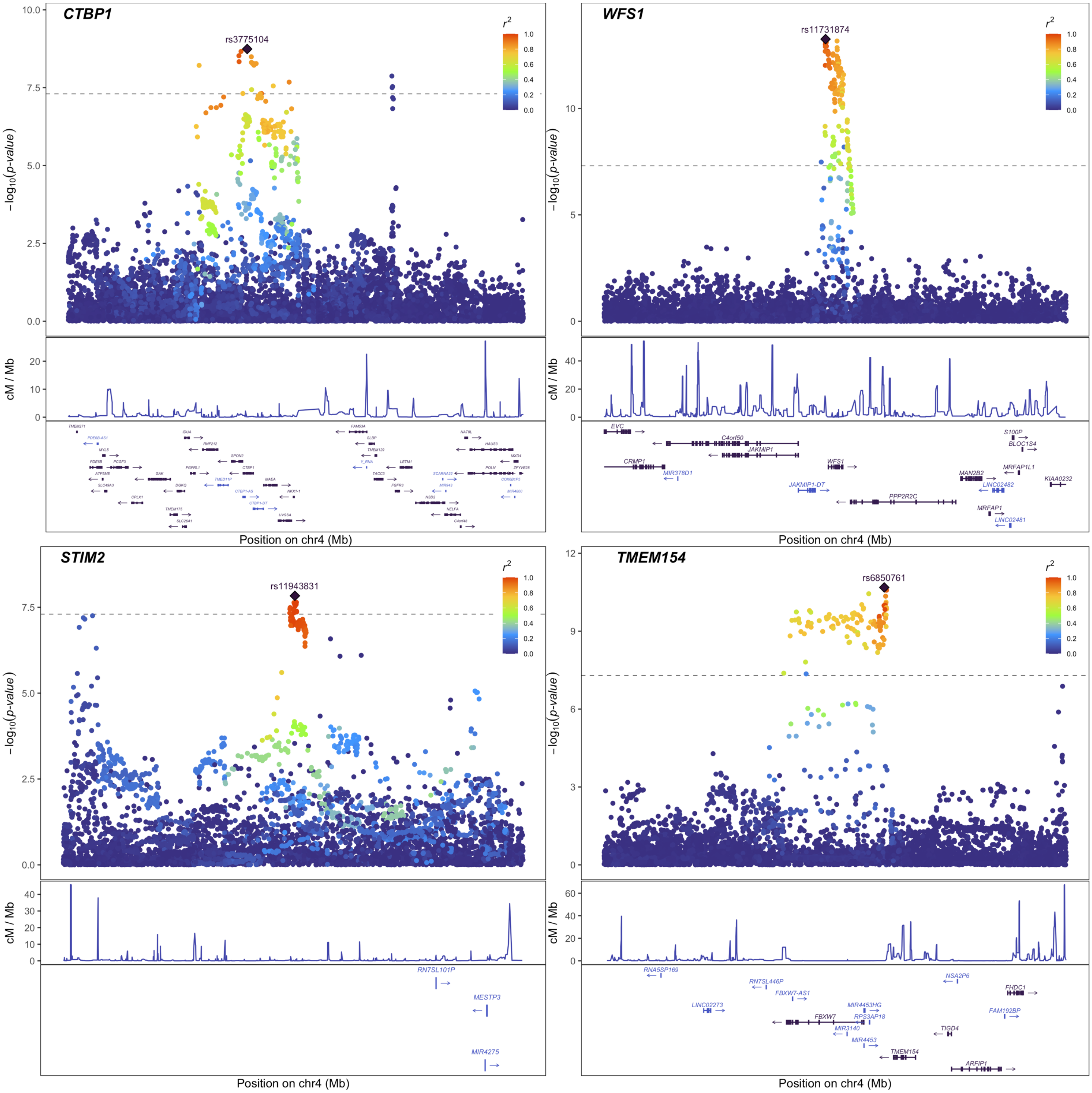


**Supplementary Fig. 1 (continued).**

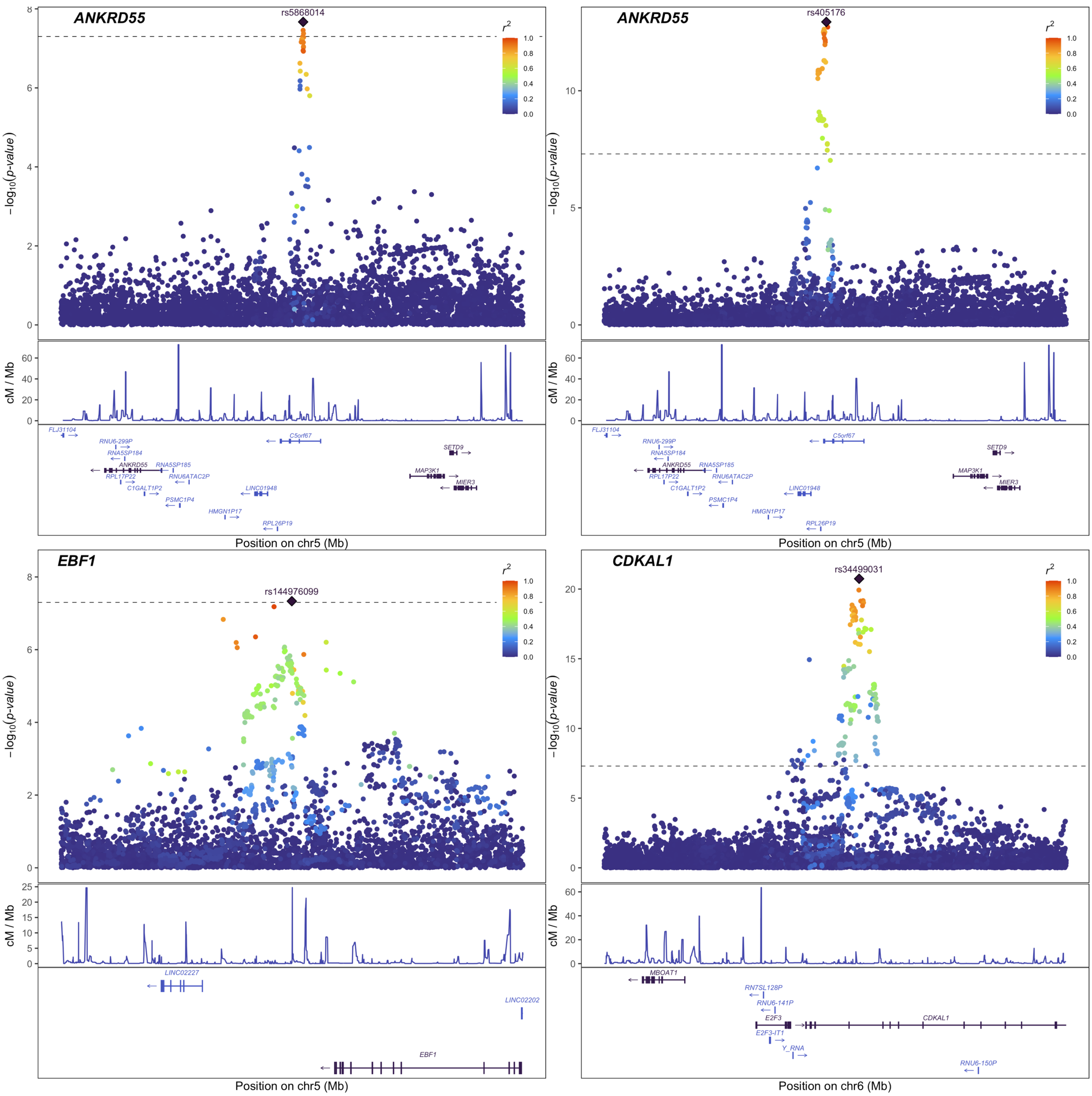


**Supplementary Fig. 1 (continued).**

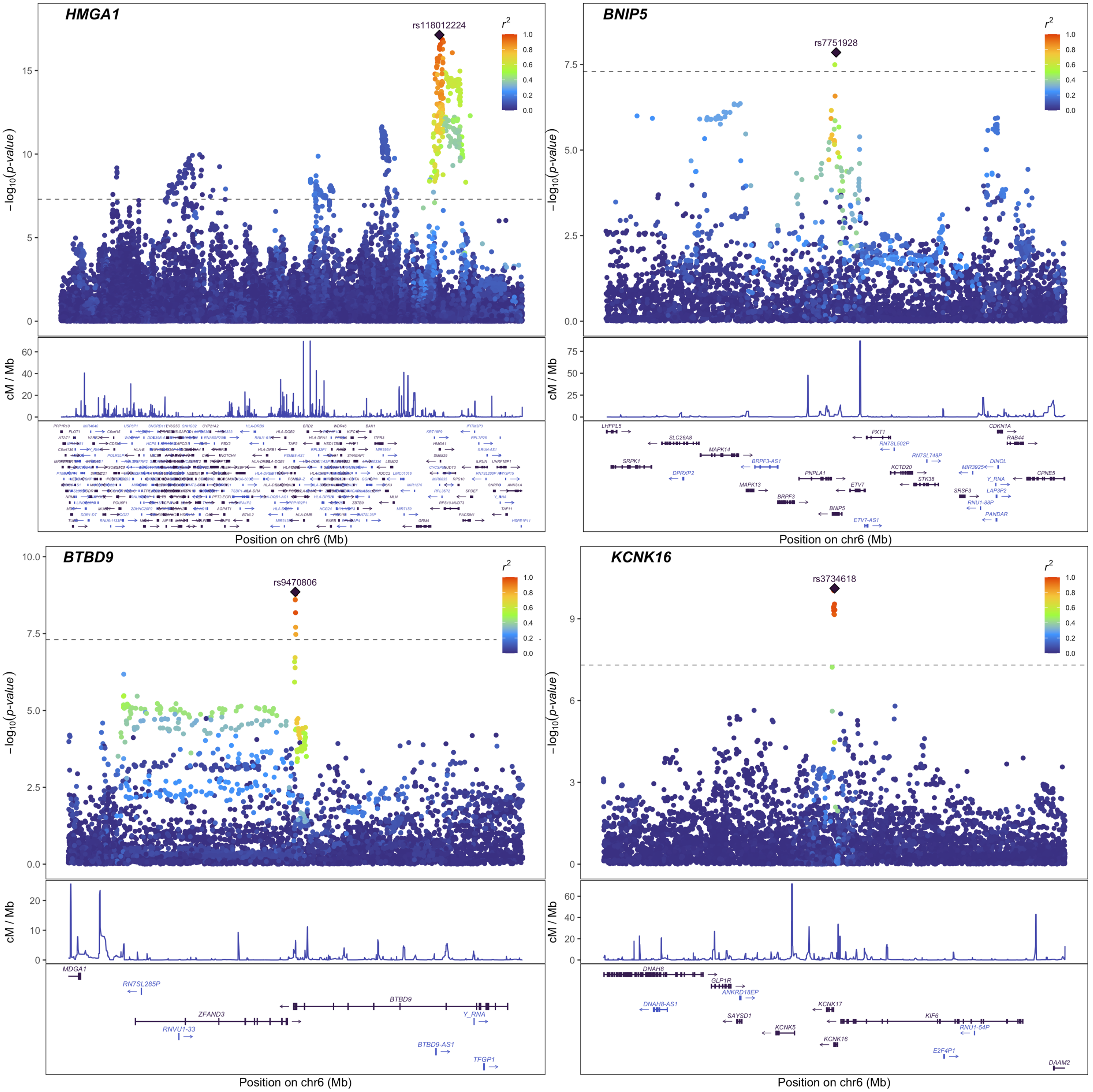


Supplementary Fig. 1 (continued).

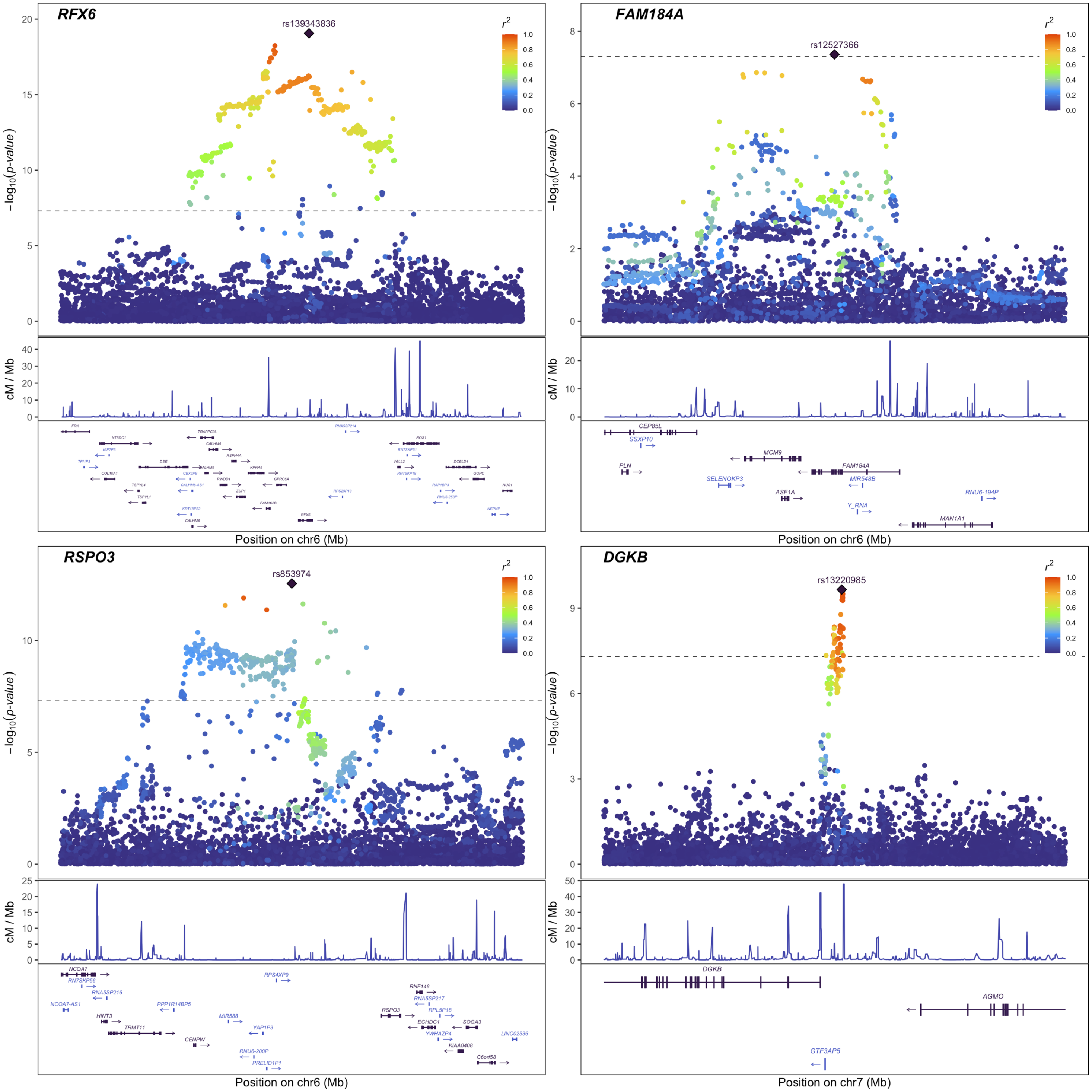


**Supplementary Fig. 1 (continued).**

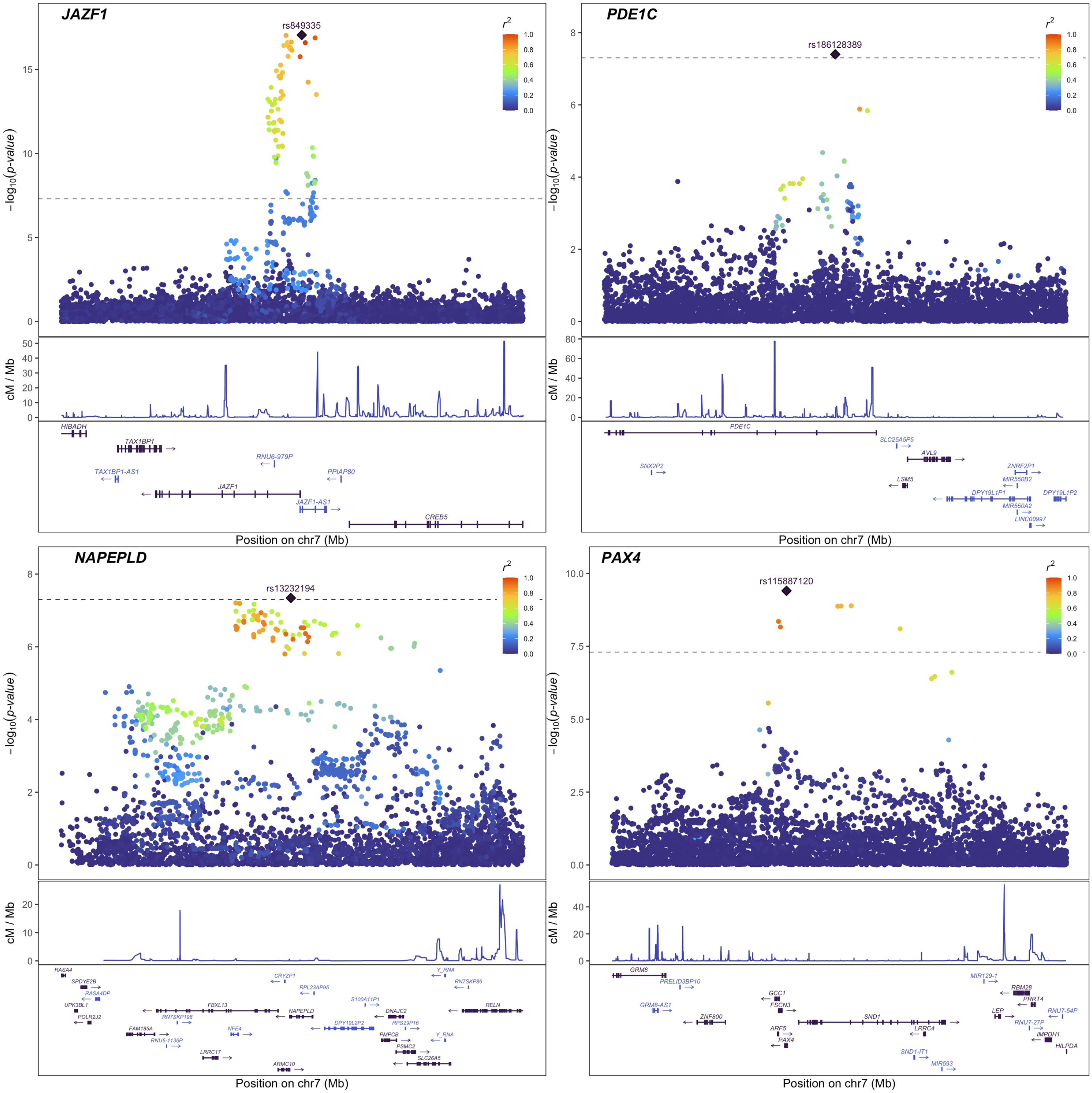


**Supplementary Fig. 1 (continued).**

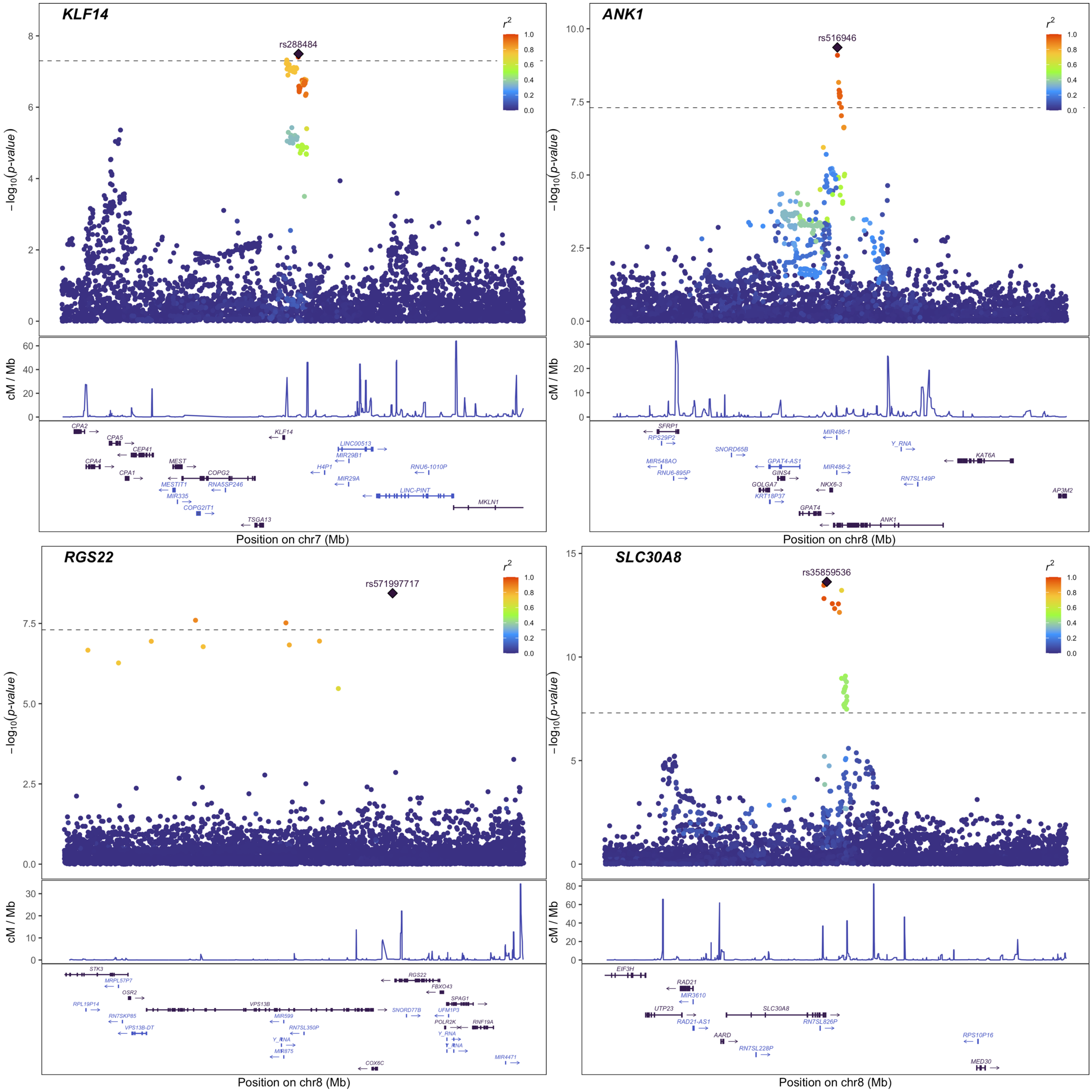


**Supplementary Fig. 1 (continued).**

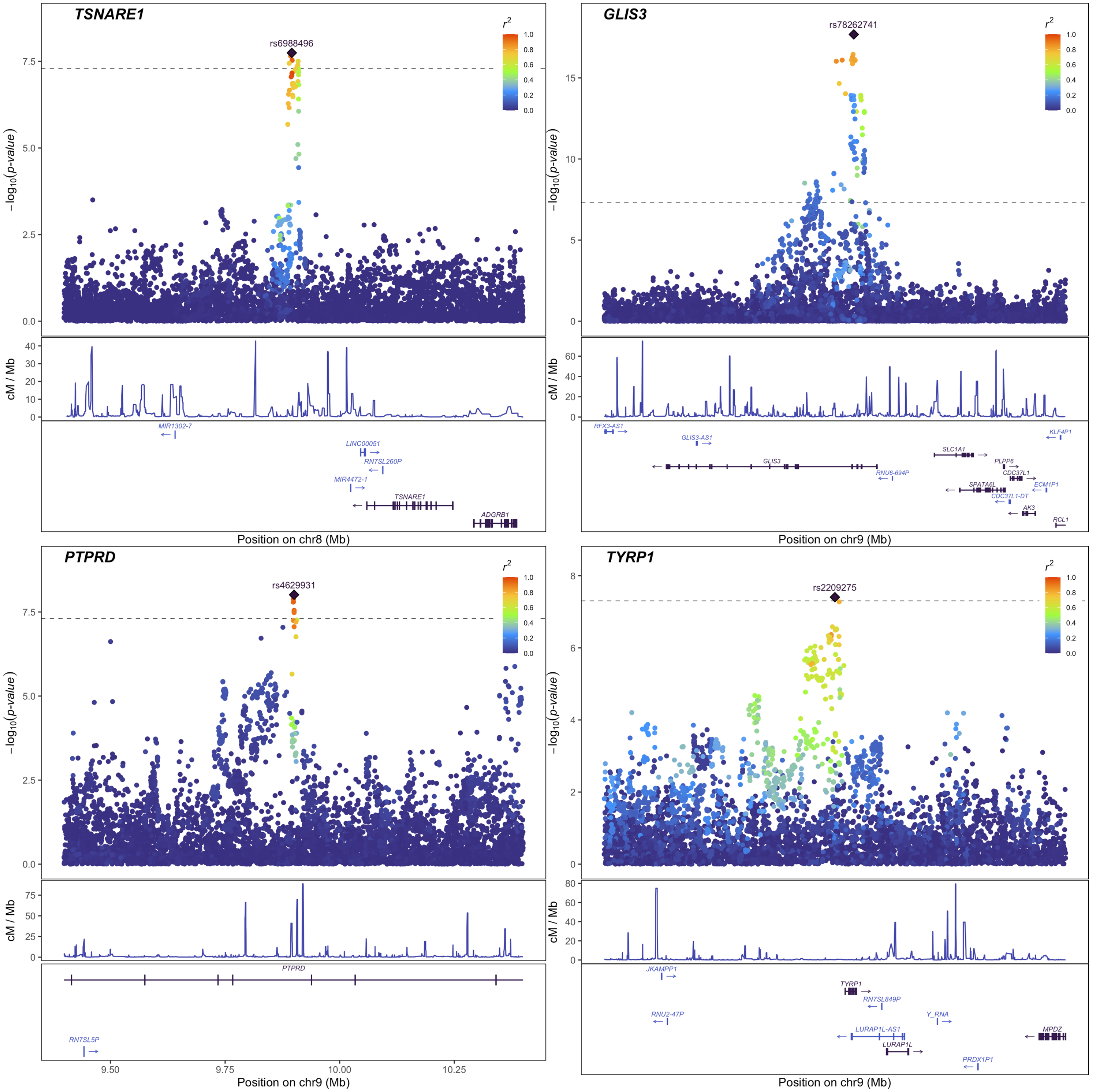


**Supplementary Fig. 1 (continued).**

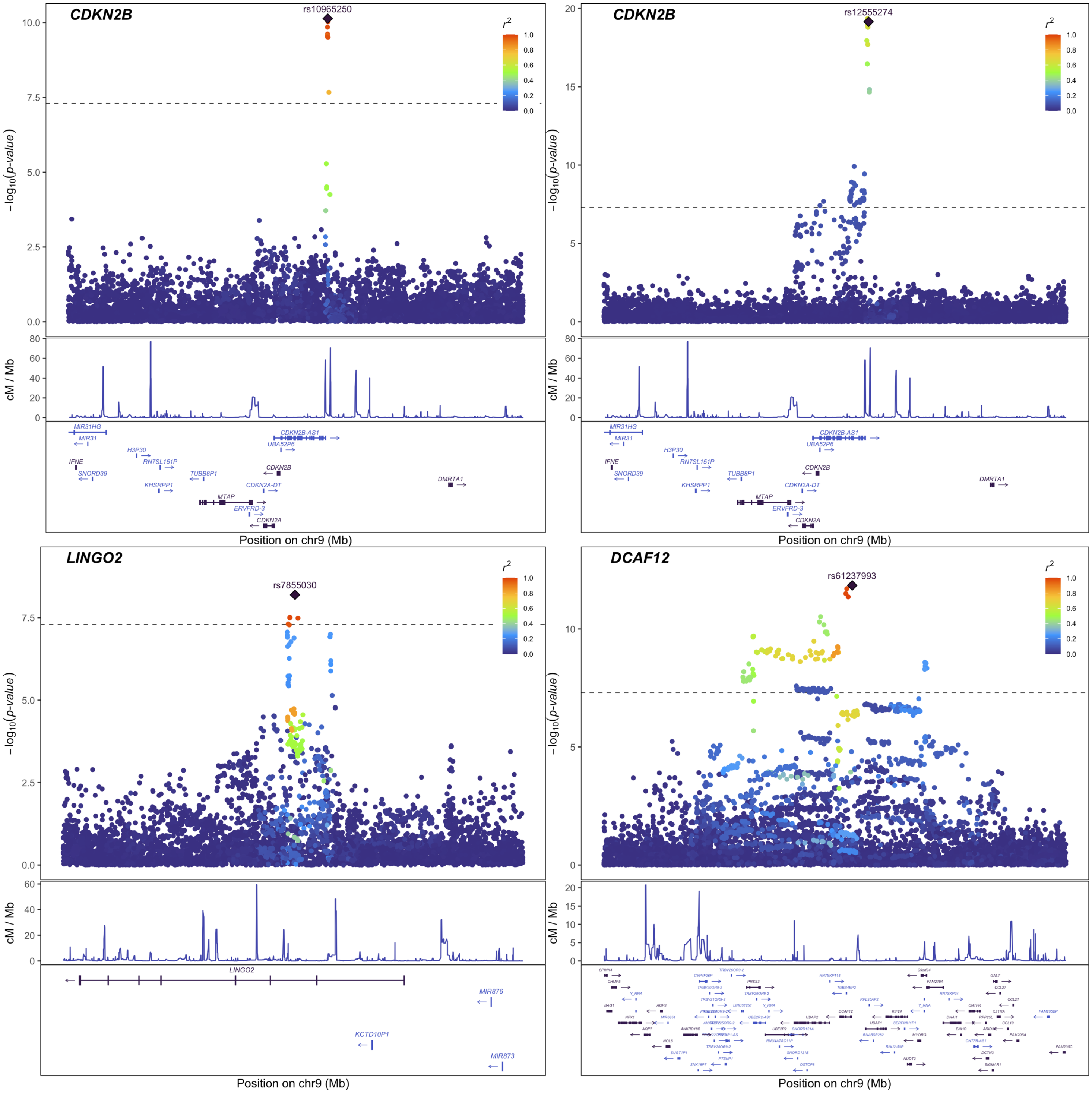


**Supplementary Fig. 1 (continued).**

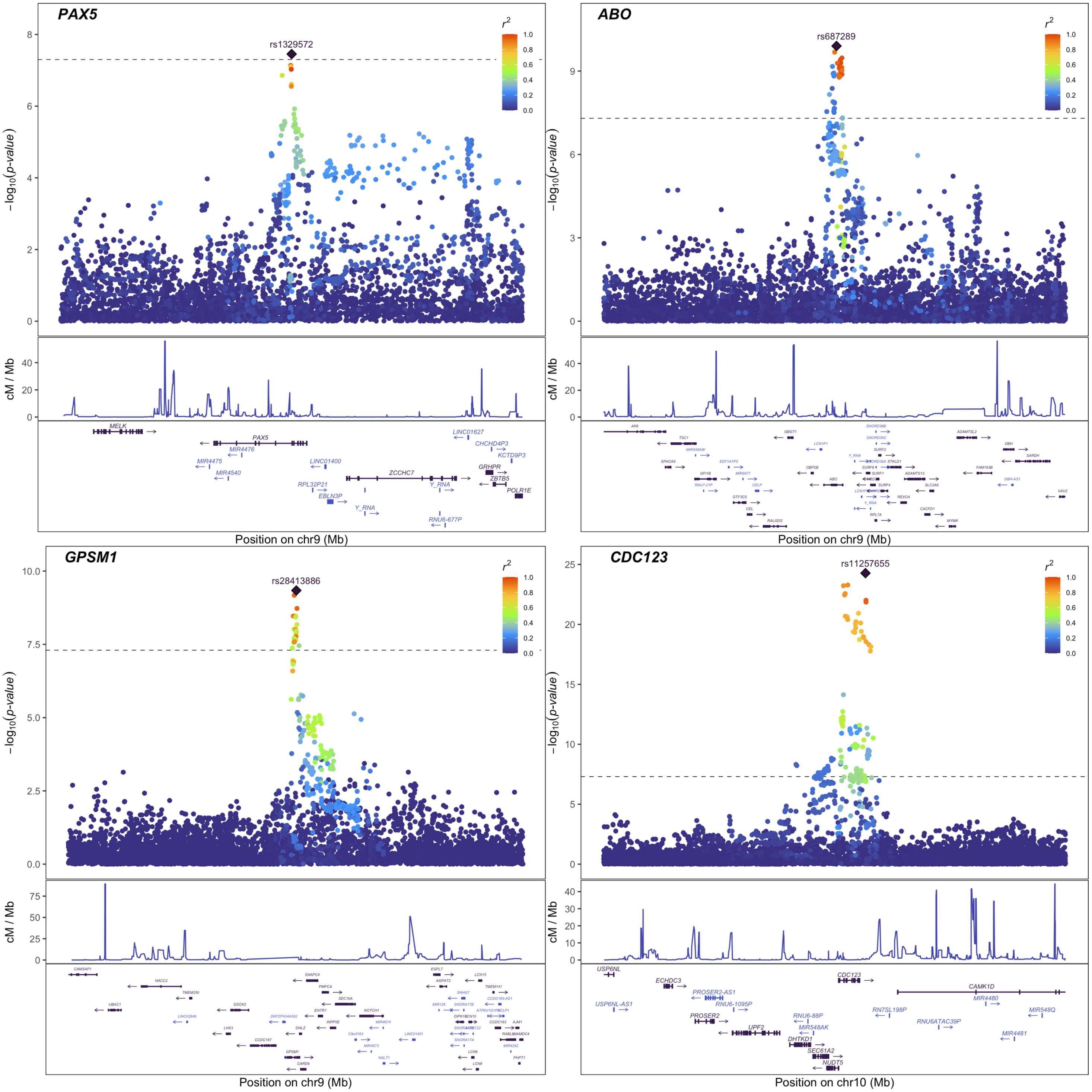


**Supplementary Fig. 1 (continued).**

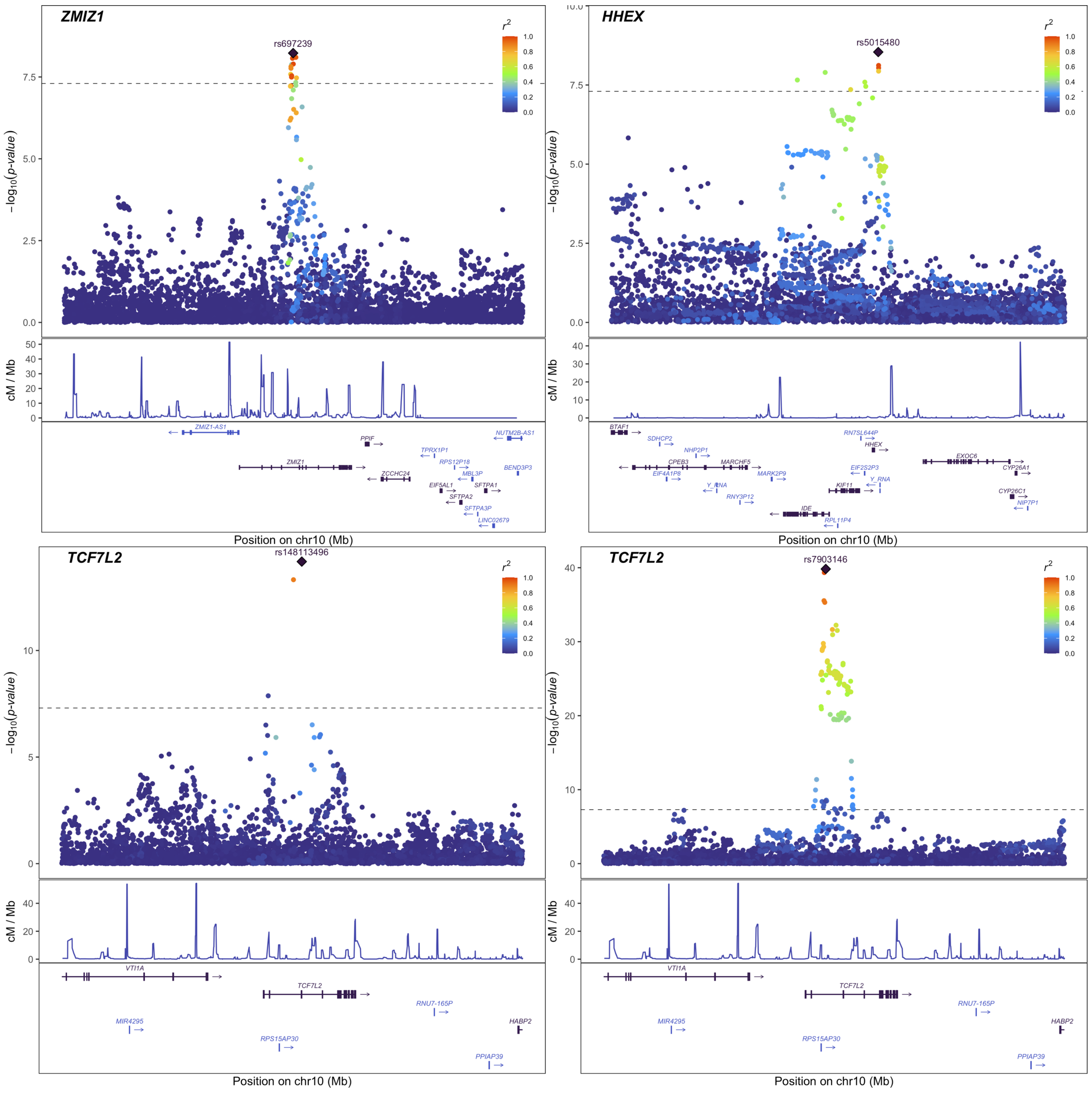


**Supplementary Fig. 1 (continued).**

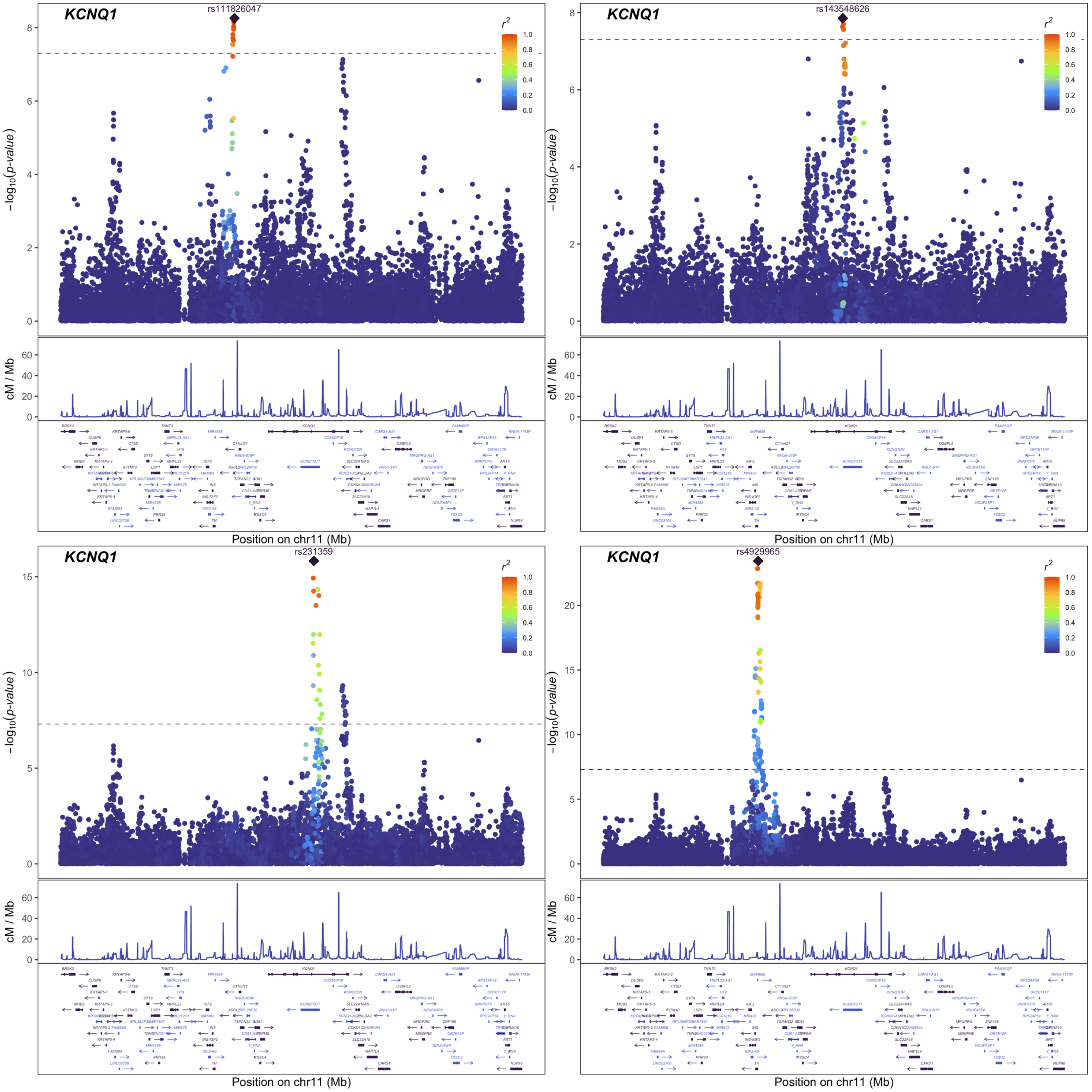


**Supplementary Fig. 1 (continued).**

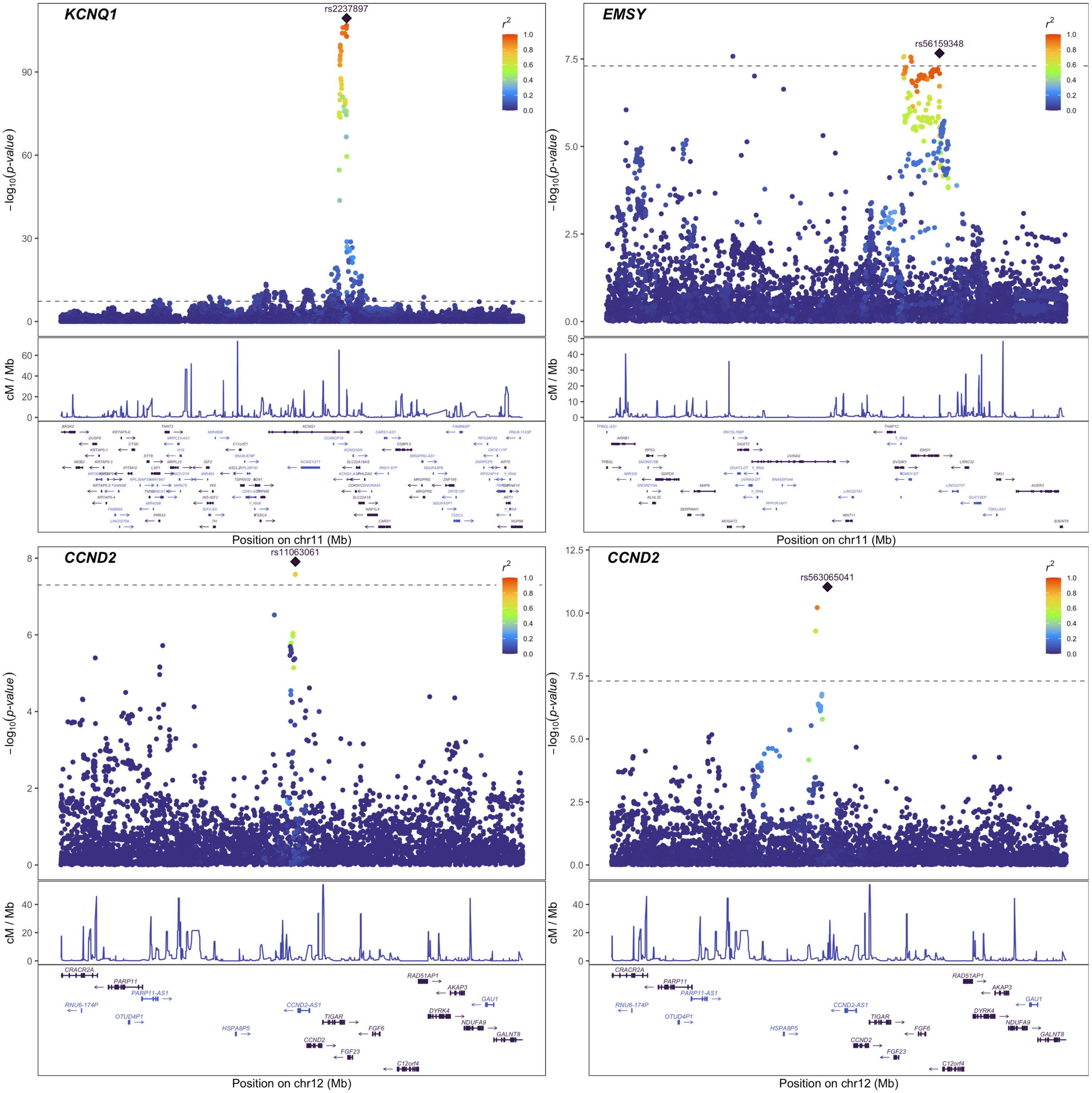


**Supplementary Fig. 1 (continued).**

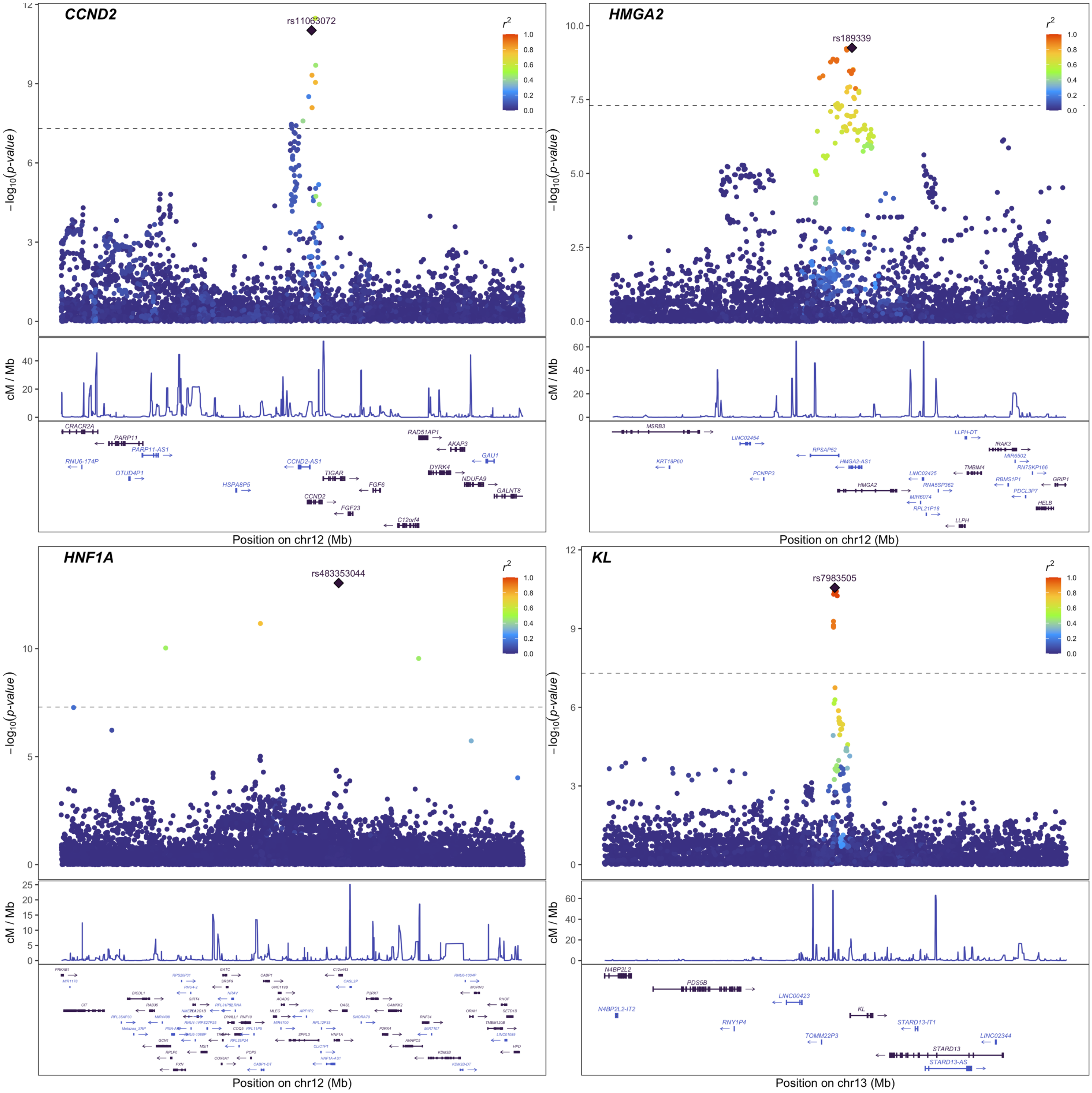


**Supplementary Fig. 1 (continued).**

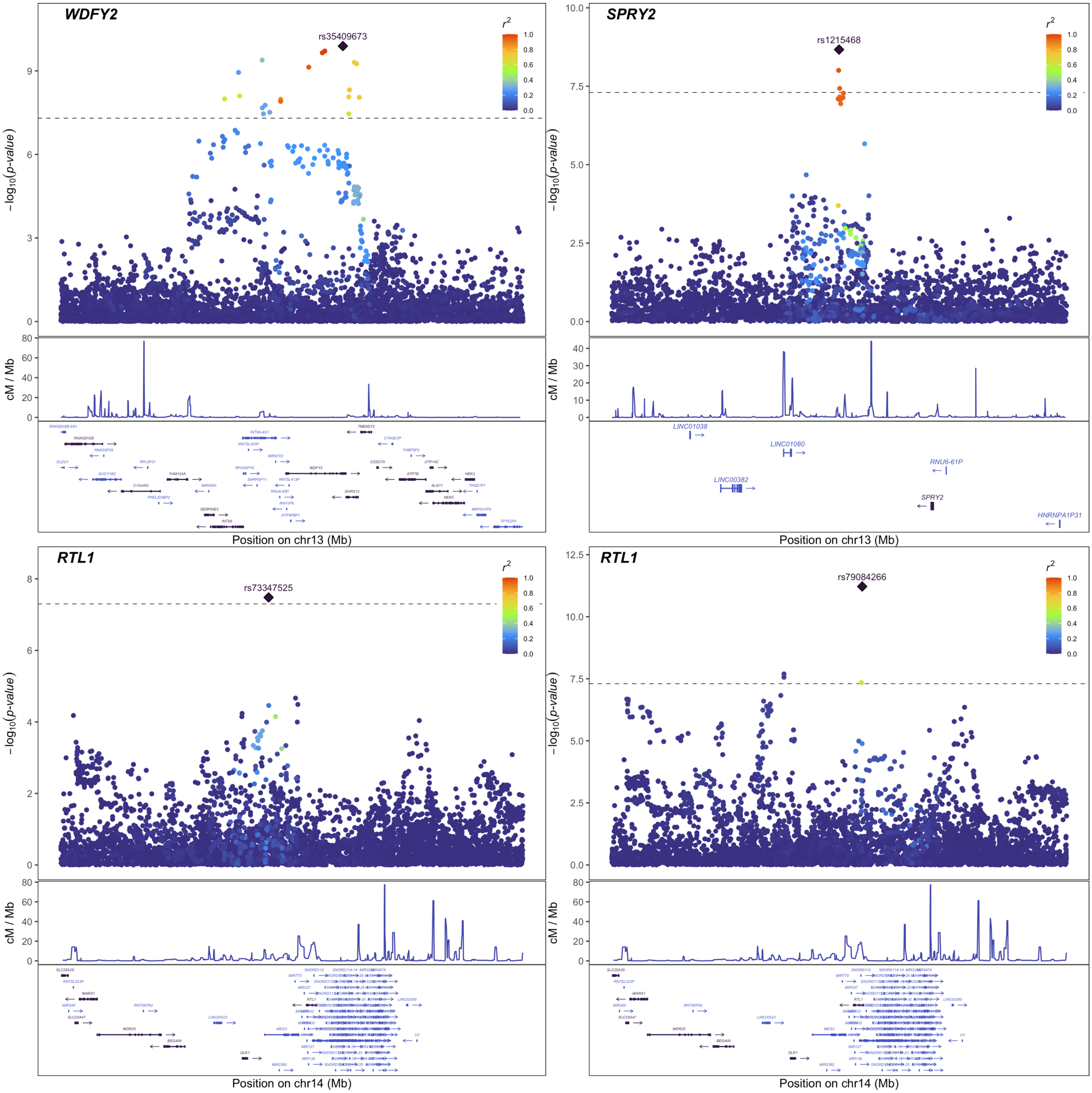


**Supplementary Fig. 1 (continued).**

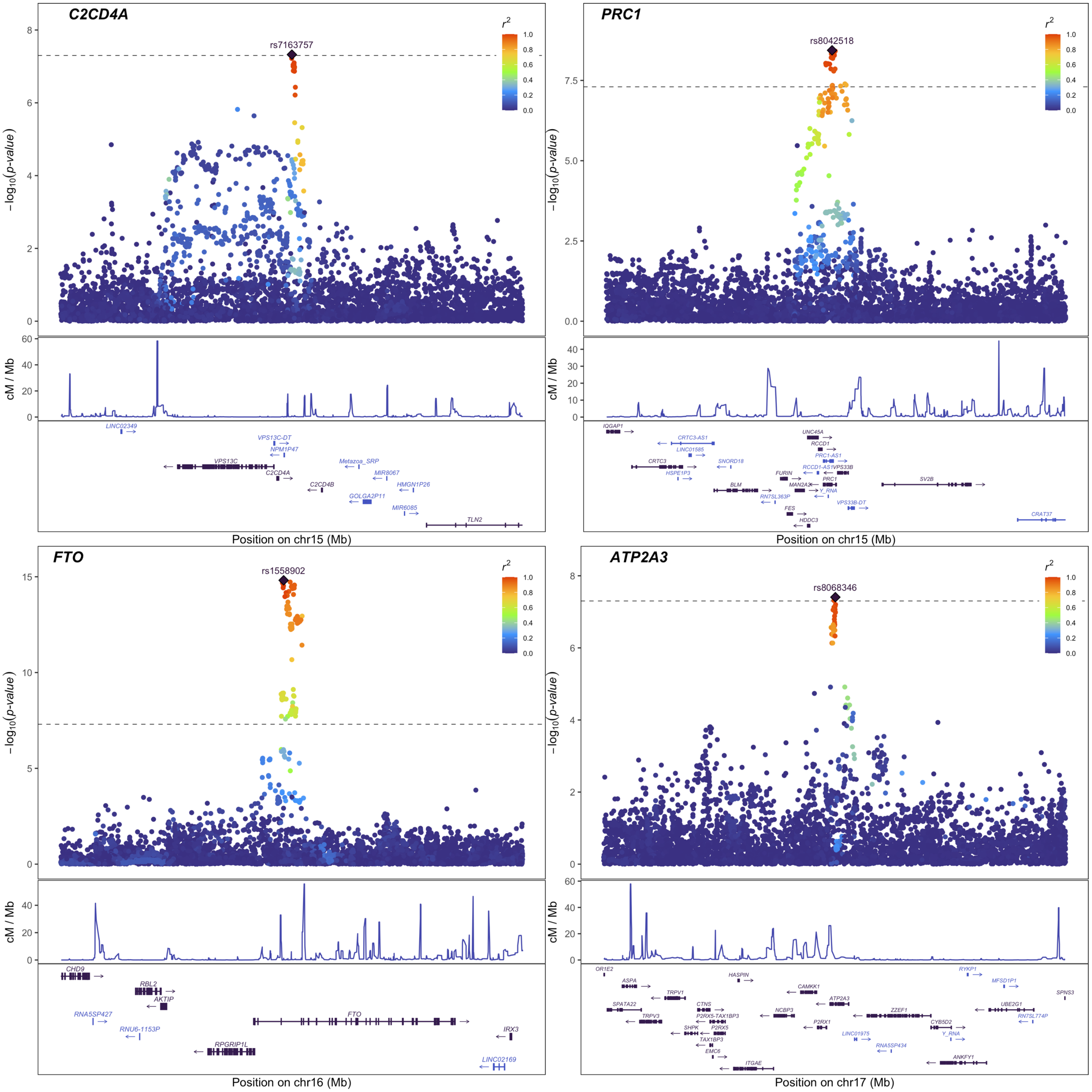


**Supplementary Fig. 1 (continued).**

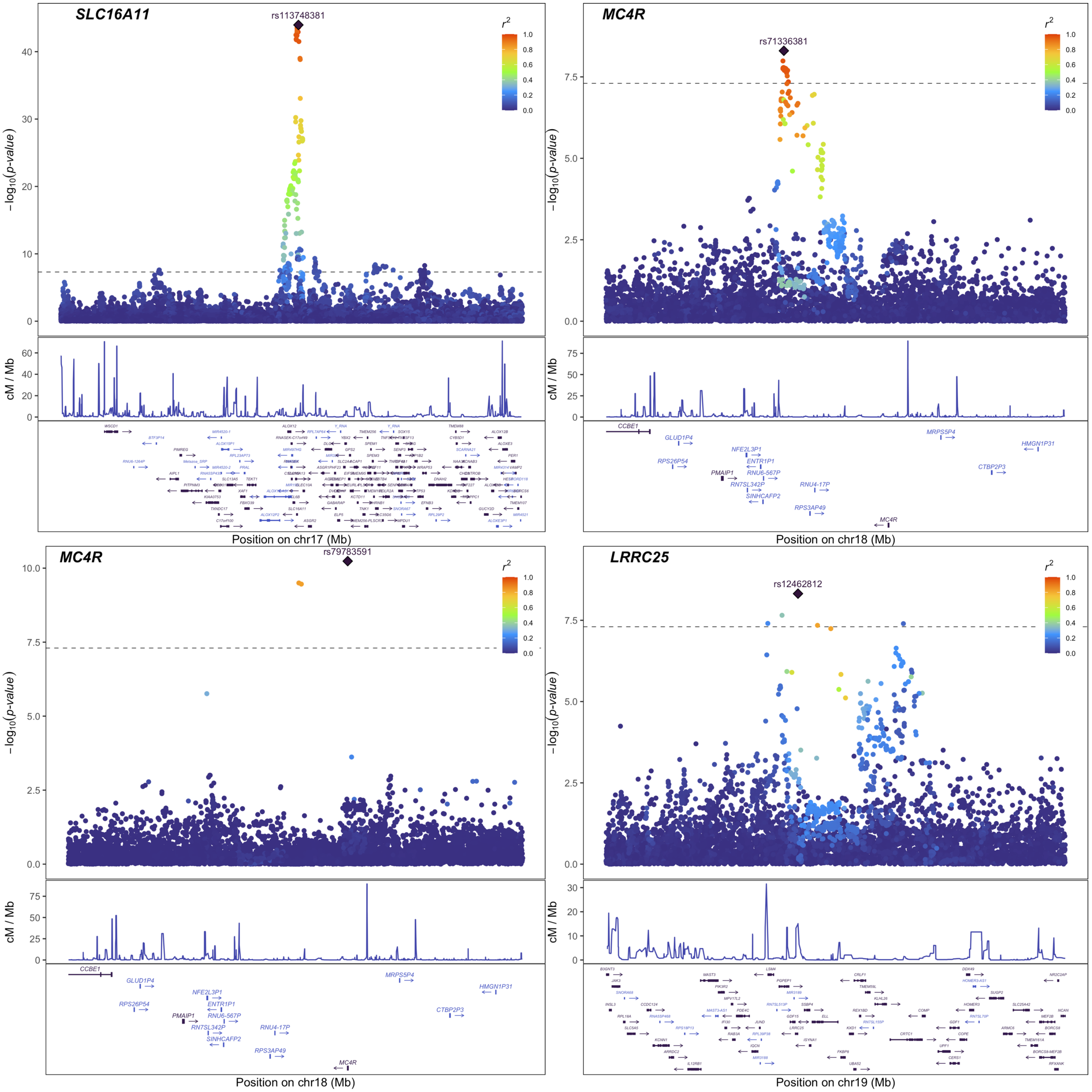


**Supplementary Fig. 1 (continued).**

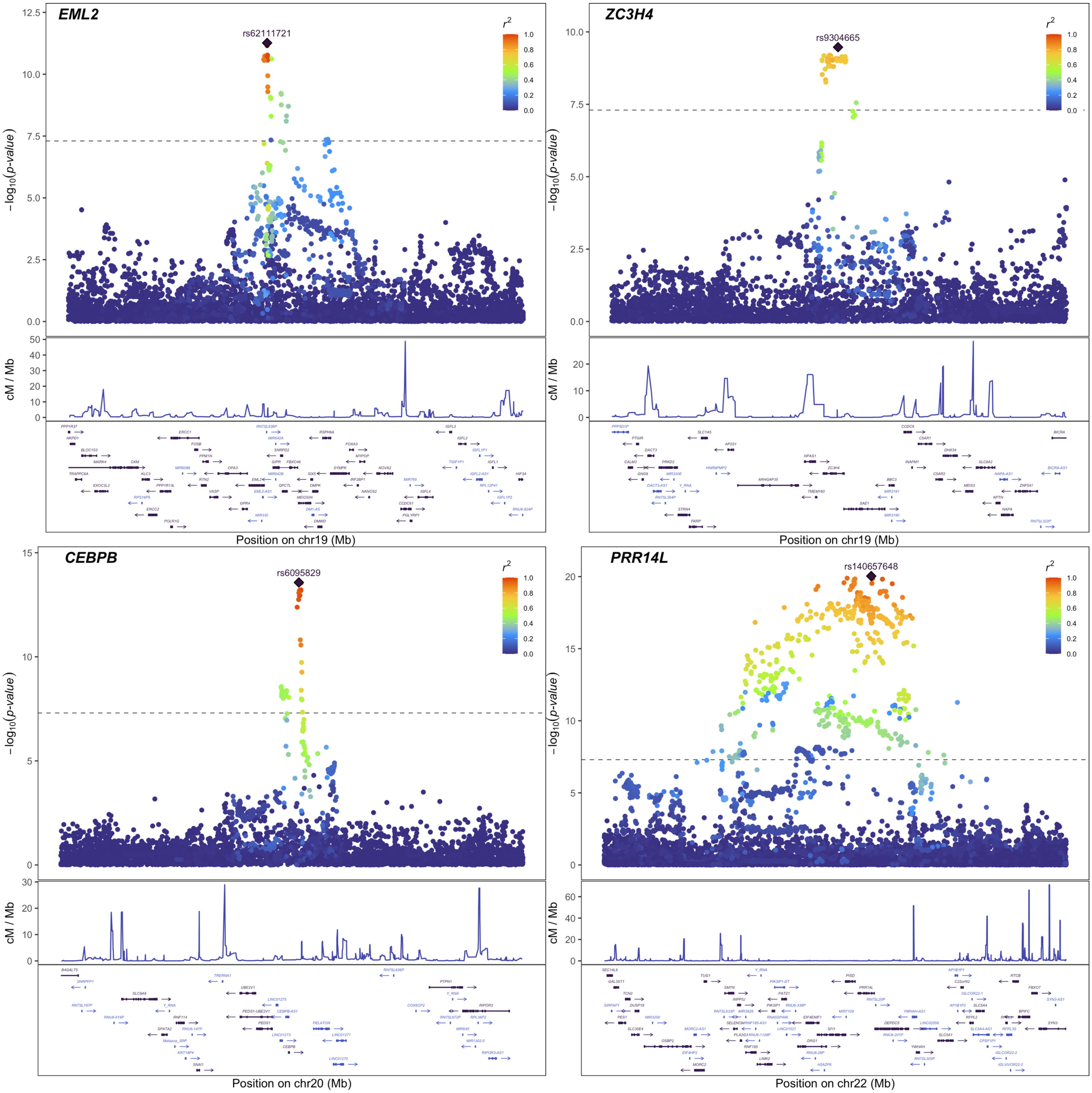


**Supplementary Fig. 1 (continued).**

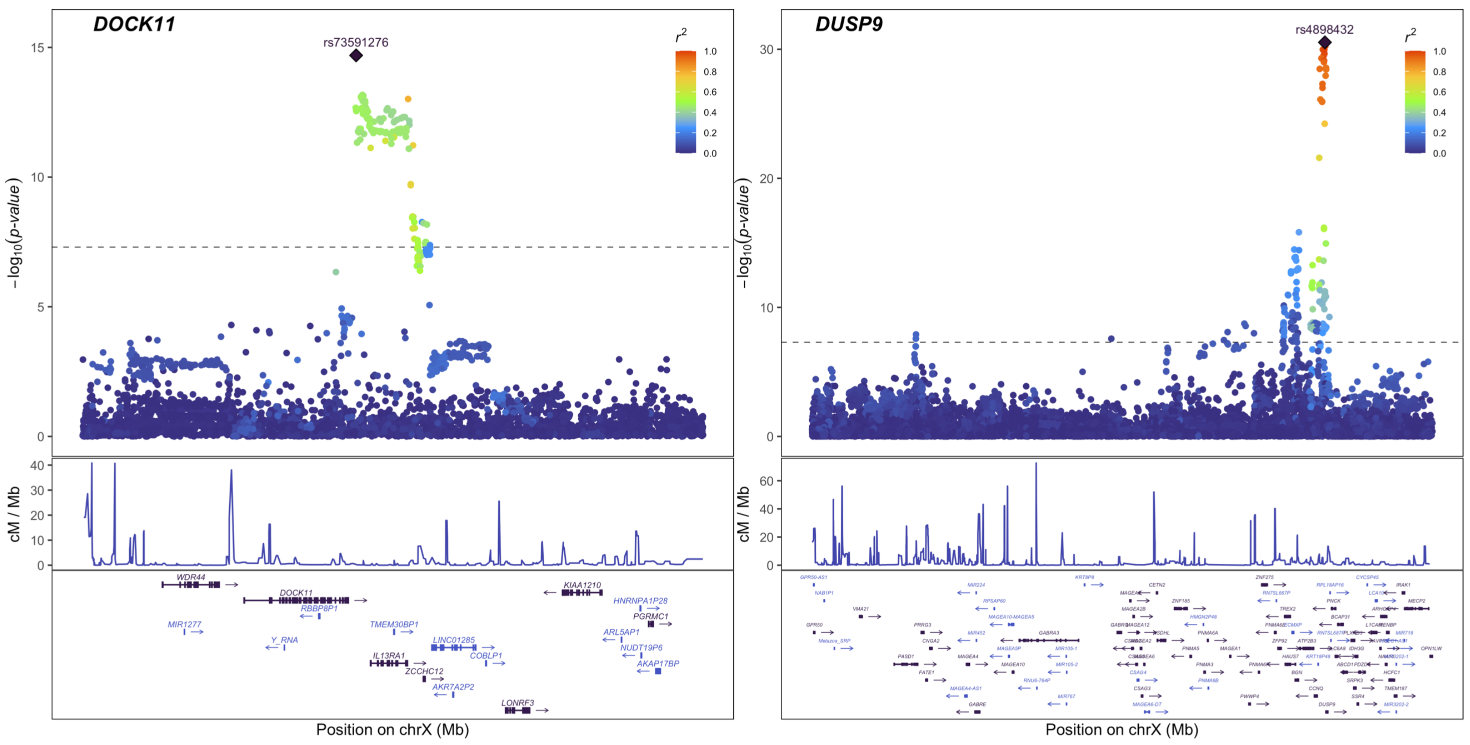


**Supplementary Fig. 1 (continued).**

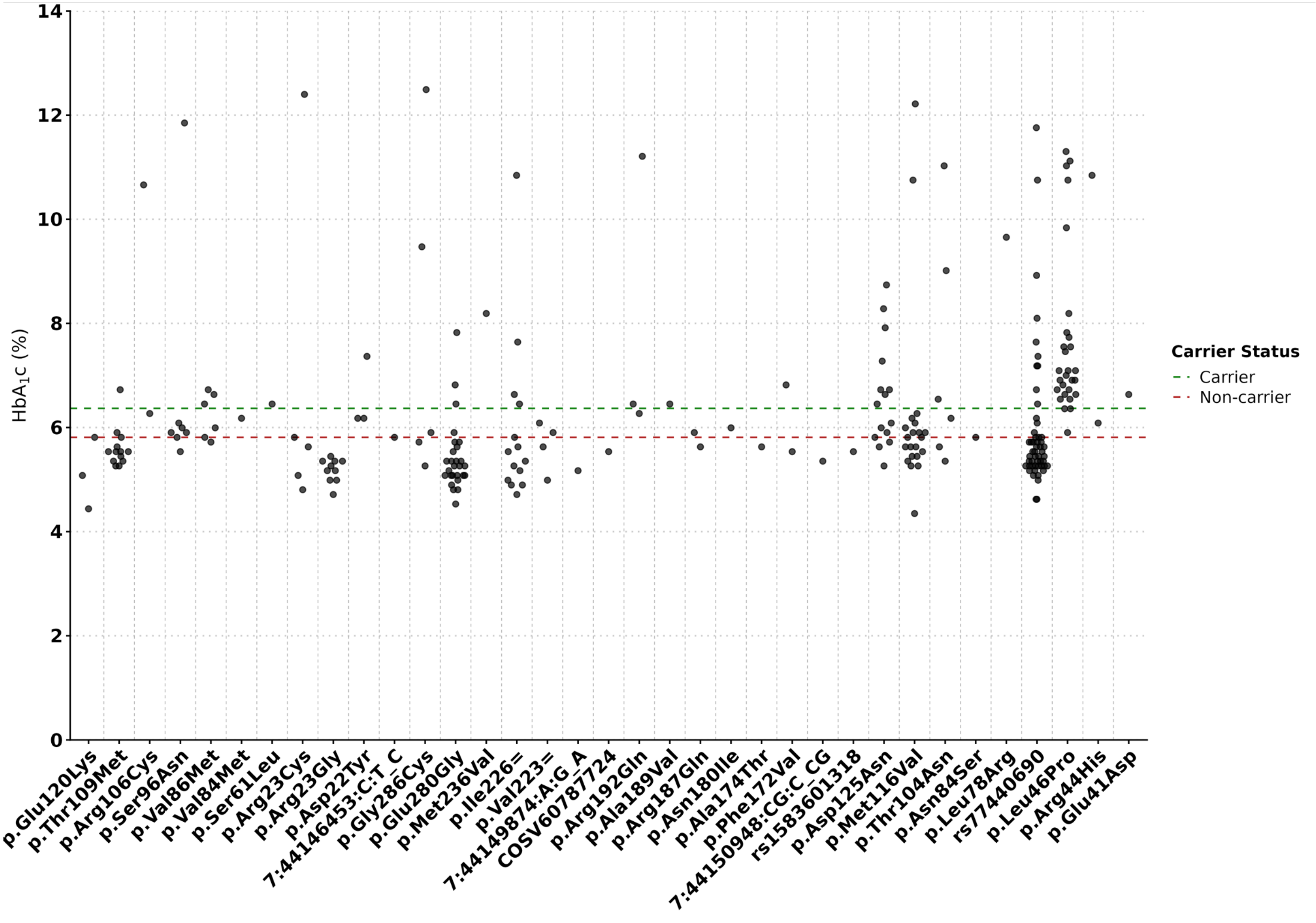


**Supplementary Fig. 2 | HbA1c levels among carriers of damaging missense variants in *GCK*.** Each point represents the HbA1c (%) value for an individual *GCK* missense carrier in MCPS. The green line indicates the mean HbA1c among *GCK* missense carriers, and the red line shows the mean across all non-carriers. rs1131691598 and rs759072800 correspond to p.Leu46Pro (NM_000162.5:c.134T>C) and p.Asp125Asn (NM_000162.5:c.370G>A), respectively. Note that these variant annotations are based on Ensemble VEP (v105), which counts the methionine at position 1, unlike ClinVar.

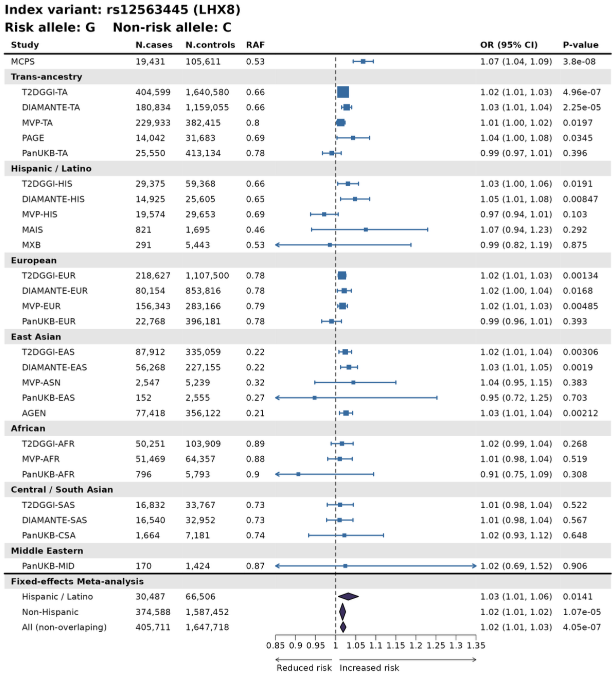


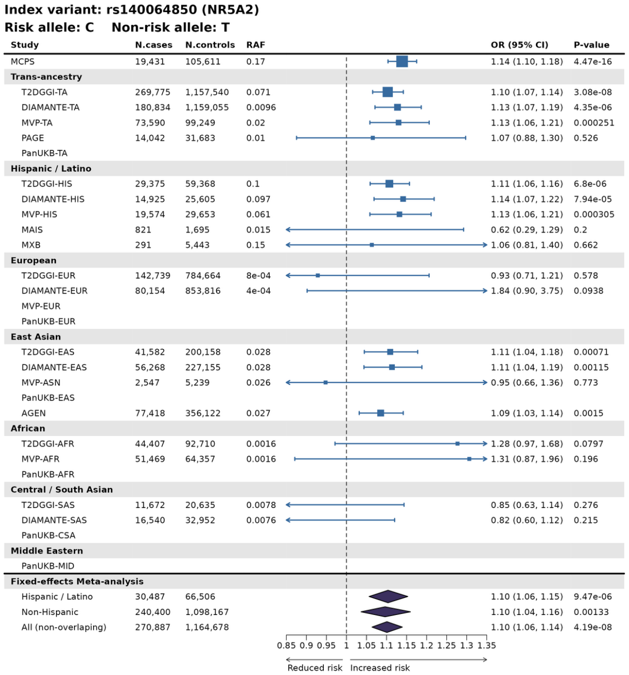


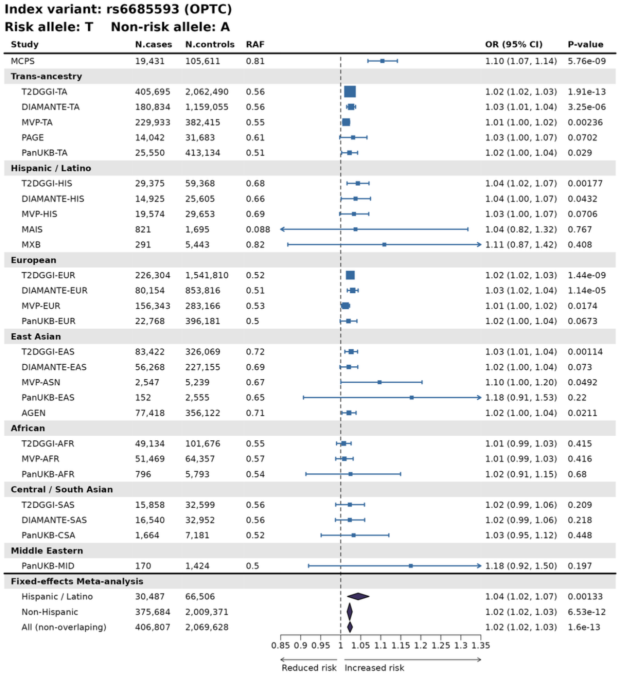


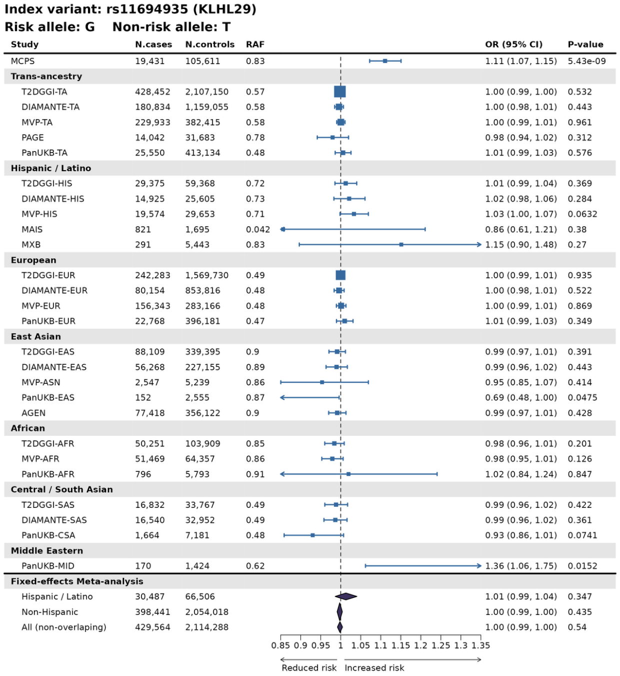


**Supplementary Fig. 3 | Forest plots for 21 novel T2D-associated signals identified in the MCPS GWAS.**

For each index variant, association statistics for the risk allele in MCPS are shown alongside results from T2DGGI, DIAMANTE, Pan-UKB, PAGE, AGEN, MXB, MVP and MAIS studies. Fixed-effects meta-analysis results (inverse-variance weighted) are displayed at the bottom of each plot for Hispanic/Latino, non-Hispanic/Latino, and all non-overlapping cohorts. RAF = risk allele frequency; TA = trans-ancestry; HIS = Hispanic/Latino; EUR = European; EAS/ASN = East Asian; AFR = African/African American; CSA = Central/South Asian; SAS = South Asian; MID = Middle Eastern.

**Index variant: rs113615666 (CLIP4)**
**Risk allele: C    Non-risk allele: CT**

| Study | N.cases | N.controls | RAF | OR (95% CI) | P-value |
|---|---|---|---|---|---|
| MCPS | 19,431 | 105,611 | 0.86 | 1.11 (1.07, 1.14) | 2.77e-08 |
| **Trans-ancestry** | | | | | |
| T2DGGI-TA | | | | | |
| DIAMANTE-TA | | | | | |
| MVP-TA | 229,933 | 382,415 | 0.7 | 1.02 (1.01, 1.03) | 7.89e-06 |
| PAGE | 14,042 | 31,683 | 0.82 | 1.02 (0.98, 1.06) | 0.358 |
| PanUKB-TA | 25,550 | 413,134 | 0.67 | 1.00 (0.98, 1.02) | 0.67 |
| **Hispanic / Latino** | | | | | |
| T2DGGI-HIS | | | | | |
| DIAMANTE-HIS | 14,925 | 25,605 | 0.8 | 1.04 (1.00, 1.08) | 0.0825 |
| MVP-HIS | 19,574 | 29,653 | 0.79 | 1.02 (0.98, 1.06) | 0.245 |
| MAIS | 821 | 1,695 | 0.1 | 1.13 (0.90, 1.40) | 0.287 |
| MXB | 291 | 5,443 | 0.87 | 0.89 (0.68, 1.16) | 0.371 |
| **European** | | | | | |
| T2DGGI-EUR | | | | | |
| DIAMANTE-EUR | | | | | |
| MVP-EUR | 156,343 | 283,166 | 0.66 | 1.02 (1.01, 1.03) | 2.73e-05 |
| PanUKB-EUR | 22,768 | 396,181 | 0.66 | 1.00 (0.98, 1.02) | 0.801 |
| **East Asian** | | | | | |
| T2DGGI-EAS | | | | | |
| DIAMANTE-EAS | | | | | |
| MVP-ASN | 2,547 | 5,239 | 0.96 | 0.86 (0.70, 1.06) | 0.153 |
| PanUKB-EAS | 152 | 2,555 | 0.97 | 0.54 (0.22, 1.34) | 0.182 |
| AGEN | 77,418 | 356,122 | 0.98 | 0.95 (0.91, 1.00) | 0.0396 |
| **African** | | | | | |
| T2DGGI-AFR | | | | | |
| MVP-AFR | 51,469 | 64,357 | 0.8 | 1.02 (0.99, 1.04) | 0.201 |
| PanUKB-AFR | 796 | 5,793 | 0.81 | 1.07 (0.93, 1.23) | 0.377 |
| **Central / South Asian** | | | | | |
| T2DGGI-SAS | | | | | |
| DIAMANTE-SAS | | | | | |
| PanUKB-CSA | 1,664 | 7,181 | 0.74 | 0.95 (0.87, 1.04) | 0.274 |
| **Middle Eastern** | | | | | |
| PanUKB-MID | 170 | 1,424 | 0.7 | 0.94 (0.73, 1.22) | 0.655 |
| **Fixed-effects Meta-analysis** | | | | | |
| Hispanic / Latino | 35,611 | 62,396 | | 1.03 (1.00, 1.06) | 0.04 |
| Non-Hispanic | 313,327 | 1,122,018 | | 1.01 (1.01, 1.02) | 0.00051 |
| All (non-overlaping) | 362,980 | 1,216,097 | | 1.01 (1.01, 1.02) | 6.36e-05 |

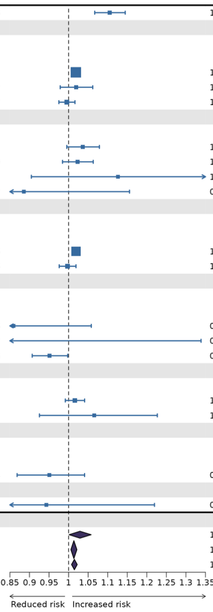


**Index variant: rs11943831 (STIM2)**
**Risk allele: A    Non-risk allele: T**

| Study | N.cases | N.controls | RAF | OR (95% CI) | P-value |
|---|---|---|---|---|---|
| MCPS | 19,431 | 105,611 | 0.18 | 1.10 (1.06, 1.13) | 6.15e-09 |
| **Trans-ancestry** | | | | | |
| T2DGGI-TA | | | | | |
| DIAMANTE-TA | | | | | |
| MVP-TA | 229,933 | 382,415 | 0.54 | 1.01 (1.00, 1.02) | 0.0456 |
| PAGE | | | | | |
| PanUKB-TA | 25,550 | 413,134 | 0.97 | 1.04 (0.98, 1.09) | 0.205 |
| **Hispanic / Latino** | | | | | |
| T2DGGI-HIS | | | | | |
| DIAMANTE-HIS | 14,925 | 25,605 | 0.32 | 1.02 (0.98, 1.07) | 0.351 |
| MVP-HIS | 19,574 | 29,653 | 0.42 | 1.03 (0.99, 1.06) | 0.119 |
| MAIS | 821 | 1,695 | 0.2 | 1.05 (0.89, 1.24) | 0.54 |
| MXB | 291 | 5,443 | 0.18 | 0.99 (0.77, 1.27) | 0.92 |
| **European** | | | | | |
| T2DGGI-EUR | | | | | |
| DIAMANTE-EUR | | | | | |
| MVP-EUR | 156,343 | 283,166 | 0.53 | 1.01 (1.00, 1.02) | 0.187 |
| PanUKB-EUR | | | | | |
| **East Asian** | | | | | |
| T2DGGI-EAS | | | | | |
| DIAMANTE-EAS | | | | | |
| MVP-ASN | 2,547 | 5,239 | 0.59 | 1.00 (0.93, 1.09) | 0.923 |
| PanUKB-EAS | | | | | |
| AGEN | | | | | |
| **African** | | | | | |
| T2DGGI-AFR | | | | | |
| MVP-AFR | 51,469 | 64,357 | 0.62 | 1.01 (0.99, 1.03) | 0.343 |
| PanUKB-AFR | | | | | |
| **Central / South Asian** | | | | | |
| T2DGGI-SAS | | | | | |
| DIAMANTE-SAS | | | | | |
| PanUKB-CSA | | | | | |
| **Middle Eastern** | | | | | |
| PanUKB-MID | | | | | |
| **Fixed-effects Meta-analysis** | | | | | |
| Hispanic / Latino | 35,611 | 62,396 | | 1.03 (1.00, 1.05) | 0.0633 |
| Non-Hispanic | 210,359 | 352,762 | | 1.01 (1.00, 1.02) | 0.106 |
| All (non-overlaping) | 271,520 | 828,292 | | 1.01 (1.00, 1.02) | 0.0179 |

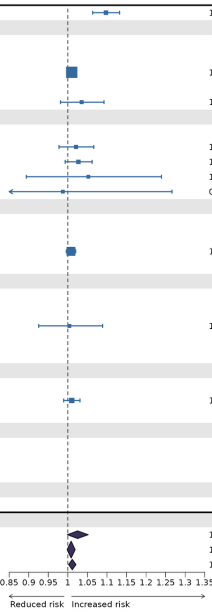


**Index variant: rs7751928 (BNIP5)**
**Risk allele: G    Non-risk allele: A**

| Study | N.cases | N.controls | RAF | OR (95% CI) | P-value |
|---|---|---|---|---|---|
| MCPS | 19,431 | 105,611 | 0.45 | 1.08 (1.05, 1.10) | 1.96e-08 |
| **Trans-ancestry** | | | | | |
| T2DGGI-TA | 428,247 | 2,105,570 | 0.73 | 1.01 (1.00, 1.01) | 0.079 |
| DIAMANTE-TA | 180,834 | 1,159,055 | 0.8 | 1.00 (0.99, 1.02) | 0.54 |
| MVP-TA | 229,933 | 382,415 | 0.83 | 1.00 (0.99, 1.02) | 0.721 |
| PAGE | 14,042 | 31,683 | 0.7 | 1.00 (0.97, 1.04) | 0.865 |
| PanUKB-TA | 25,550 | 413,134 | 0.88 | 0.98 (0.95, 1.01) | 0.216 |
| **Hispanic / Latino** | | | | | |
| T2DGGI-HIS | 29,170 | 57,793 | 0.63 | 1.03 (1.00, 1.06) | 0.0257 |
| DIAMANTE-HIS | 14,925 | 25,605 | 0.62 | 1.01 (0.97, 1.04) | 0.723 |
| MVP-HIS | 19,574 | 29,653 | 0.67 | 1.01 (0.97, 1.05) | 0.546 |
| MAIS | 821 | 1,695 | 0.35 | 1.00 (0.87, 1.15) | 0.983 |
| MXB | 291 | 5,443 | 0.46 | 0.96 (0.79, 1.16) | 0.693 |
| **European** | | | | | |
| T2DGGI-EUR | 242,283 | 1,569,730 | 0.87 | 1.00 (0.99, 1.01) | 0.592 |
| DIAMANTE-EUR | 80,154 | 853,816 | 0.88 | 0.99 (0.98, 1.01) | 0.568 |
| MVP-EUR | 156,343 | 283,166 | 0.88 | 1.00 (0.98, 1.02) | 0.85 |
| PanUKB-EUR | 22,768 | 396,181 | 0.88 | 0.99 (0.96, 1.02) | 0.451 |
| **East Asian** | | | | | |
| T2DGGI-EAS | 88,109 | 339,395 | 0.56 | 1.01 (1.00, 1.02) | 0.124 |
| DIAMANTE-EAS | 56,268 | 227,155 | 0.57 | 1.01 (1.00, 1.03) | 0.143 |
| MVP-ASN | 2,547 | 5,239 | 0.67 | 1.08 (0.99, 1.19) | 0.0954 |
| PanUKB-EAS | 152 | 2,555 | 0.62 | 1.09 (0.83, 1.43) | 0.53 |
| AGEN | 77,418 | 356,122 | 0.56 | 1.01 (1.00, 1.03) | 0.0985 |
| **African** | | | | | |
| T2DGGI-AFR | 50,251 | 103,909 | 0.74 | 1.00 (0.98, 1.02) | 0.805 |
| MVP-AFR | 51,469 | 64,357 | 0.74 | 0.99 (0.97, 1.02) | 0.666 |
| PanUKB-AFR | 796 | 5,793 | 0.74 | 1.05 (0.92, 1.20) | 0.448 |
| **Central / South Asian** | | | | | |
| T2DGGI-SAS | 16,832 | 33,767 | 0.82 | 0.98 (0.95, 1.02) | 0.368 |
| DIAMANTE-SAS | 16,540 | 32,952 | 0.82 | 0.98 (0.94, 1.02) | 0.274 |
| PanUKB-CSA | 1,664 | 7,181 | 0.83 | 0.84 (0.75, 0.94) | 0.00225 |
| **Middle Eastern** | | | | | |
| PanUKB-MID | 170 | 1,424 | 0.84 | 1.05 (0.74, 1.49) | 0.785 |
| **Fixed-effects Meta-analysis** | | | | | |
| Hispanic / Latino | 30,282 | 64,931 | | 1.03 (1.00, 1.05) | 0.034 |
| Non-Hispanic | 398,441 | 2,054,018 | | 1.00 (1.00, 1.01) | 0.217 |
| All (non-overlaping) | 429,359 | 2,112,708 | | 1.01 (1.00, 1.01) | 0.0824 |

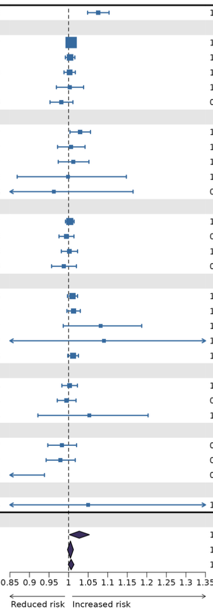


**Index variant: rs9470806 (BTBD9)**
**Risk allele: A    Non-risk allele: G**

| Study | N.cases | N.controls | RAF | OR (95% CI) | P-value |
|---|---|---|---|---|---|
| MCPS | 19,431 | 105,611 | 0.33 | 1.08 (1.06, 1.11) | 4.64e-10 |
| **Trans-ancestry** | | | | | |
| T2DGGI-TA | 428,452 | 2,107,150 | 0.58 | 0.99 (0.99, 1.00) | 0.0104 |
| DIAMANTE-TA | 180,834 | 1,159,055 | 0.59 | 1.00 (0.99, 1.01) | 0.752 |
| MVP-TA | 229,933 | 382,415 | 0.64 | 0.99 (0.98, 1.00) | 0.117 |
| PAGE | 14,042 | 31,683 | 0.45 | 0.98 (0.95, 1.01) | 0.234 |
| PanUKB-TA | 25,550 | 413,134 | 0.73 | 1.01 (0.99, 1.03) | 0.416 |
| **Hispanic / Latino** | | | | | |
| T2DGGI-HIS | 29,375 | 59,368 | 0.49 | 1.01 (0.99, 1.04) | 0.294 |
| DIAMANTE-HIS | 14,925 | 25,605 | 0.48 | 1.01 (0.98, 1.04) | 0.531 |
| MVP-HIS | 19,574 | 29,653 | 0.5 | 1.01 (0.98, 1.04) | 0.621 |
| MAIS | 821 | 1,695 | 0.24 | 0.98 (0.83, 1.15) | 0.77 |
| MXB | 291 | 5,443 | 0.32 | 1.01 (0.82, 1.23) | 0.937 |
| **European** | | | | | |
| T2DGGI-EUR | 242,283 | 1,569,730 | 0.72 | 0.99 (0.98, 1.00) | 0.00503 |
| DIAMANTE-EUR | 80,154 | 853,816 | 0.74 | 0.99 (0.98, 1.01) | 0.317 |
| MVP-EUR | 156,343 | 283,166 | 0.72 | 0.99 (0.98, 1.00) | 0.119 |
| PanUKB-EUR | 22,768 | 396,181 | 0.75 | 1.01 (0.99, 1.04) | 0.254 |
| **East Asian** | | | | | |
| T2DGGI-EAS | 88,109 | 339,395 | 0.15 | 1.00 (0.98, 1.02) | 0.982 |
| DIAMANTE-EAS | 56,268 | 227,155 | 0.14 | 1.01 (0.98, 1.03) | 0.552 |
| MVP-ASN | 2,547 | 5,239 | 0.2 | 1.09 (0.98, 1.20) | 0.104 |
| PanUKB-EAS | 152 | 2,555 | 0.17 | 1.15 (0.83, 1.60) | 0.405 |
| AGEN | 77,418 | 356,122 | 0.15 | 1.00 (0.98, 1.01) | 0.621 |
| **African** | | | | | |
| T2DGGI-AFR | 50,251 | 103,909 | 0.46 | 0.99 (0.97, 1.00) | 0.159 |
| MVP-AFR | 51,469 | 64,357 | 0.44 | 0.99 (0.97, 1.01) | 0.195 |
| PanUKB-AFR | 796 | 5,793 | 0.43 | 1.01 (0.90, 1.13) | 0.852 |
| **Central / South Asian** | | | | | |
| T2DGGI-SAS | 16,832 | 33,767 | 0.47 | 0.99 (0.97, 1.02) | 0.653 |
| DIAMANTE-SAS | 16,540 | 32,952 | 0.47 | 0.99 (0.96, 1.02) | 0.587 |
| PanUKB-CSA | 1,664 | 7,181 | 0.48 | 0.94 (0.86, 1.01) | 0.102 |
| **Middle Eastern** | | | | | |
| PanUKB-MID | 170 | 1,424 | 0.63 | 1.17 (0.91, 1.51) | 0.211 |
| **Fixed-effects Meta-analysis** | | | | | |
| Hispanic / Latino | 30,487 | 66,506 | | 1.01 (0.99, 1.04) | 0.319 |
| Non-Hispanic | 398,441 | 2,054,018 | | 0.99 (0.98, 1.00) | 0.00392 |
| All (non-overlaping) | 429,564 | 2,114,288 | | 0.99 (0.99, 1.00) | 0.0102 |

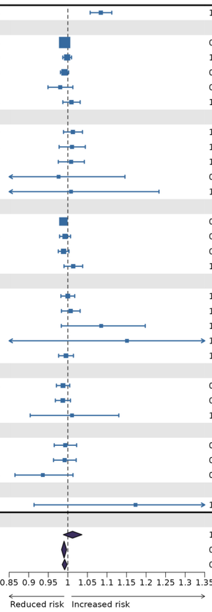


**Supplementary Fig. 3 (continued).**

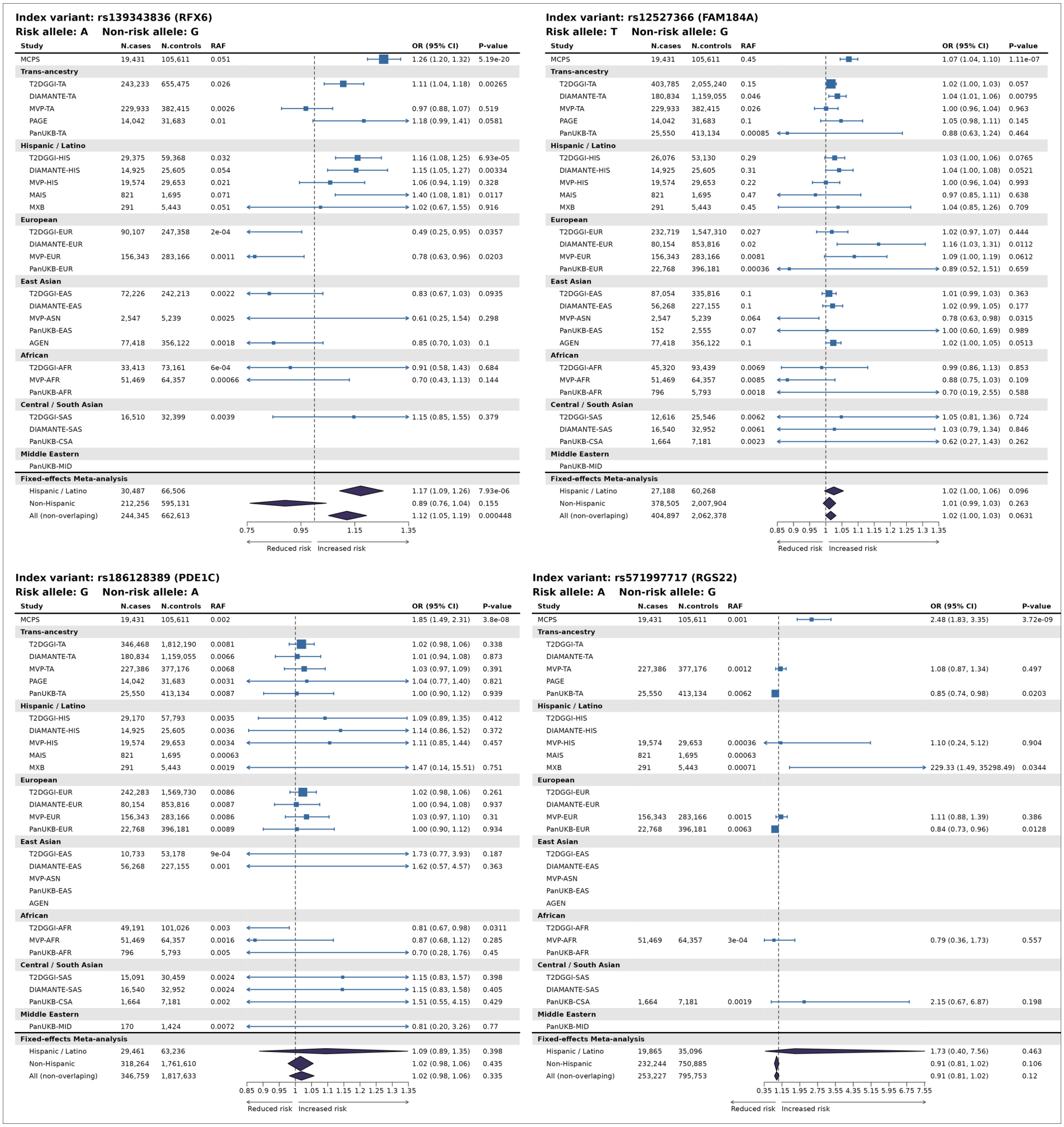


**Supplementary Fig. 3 (continued).**

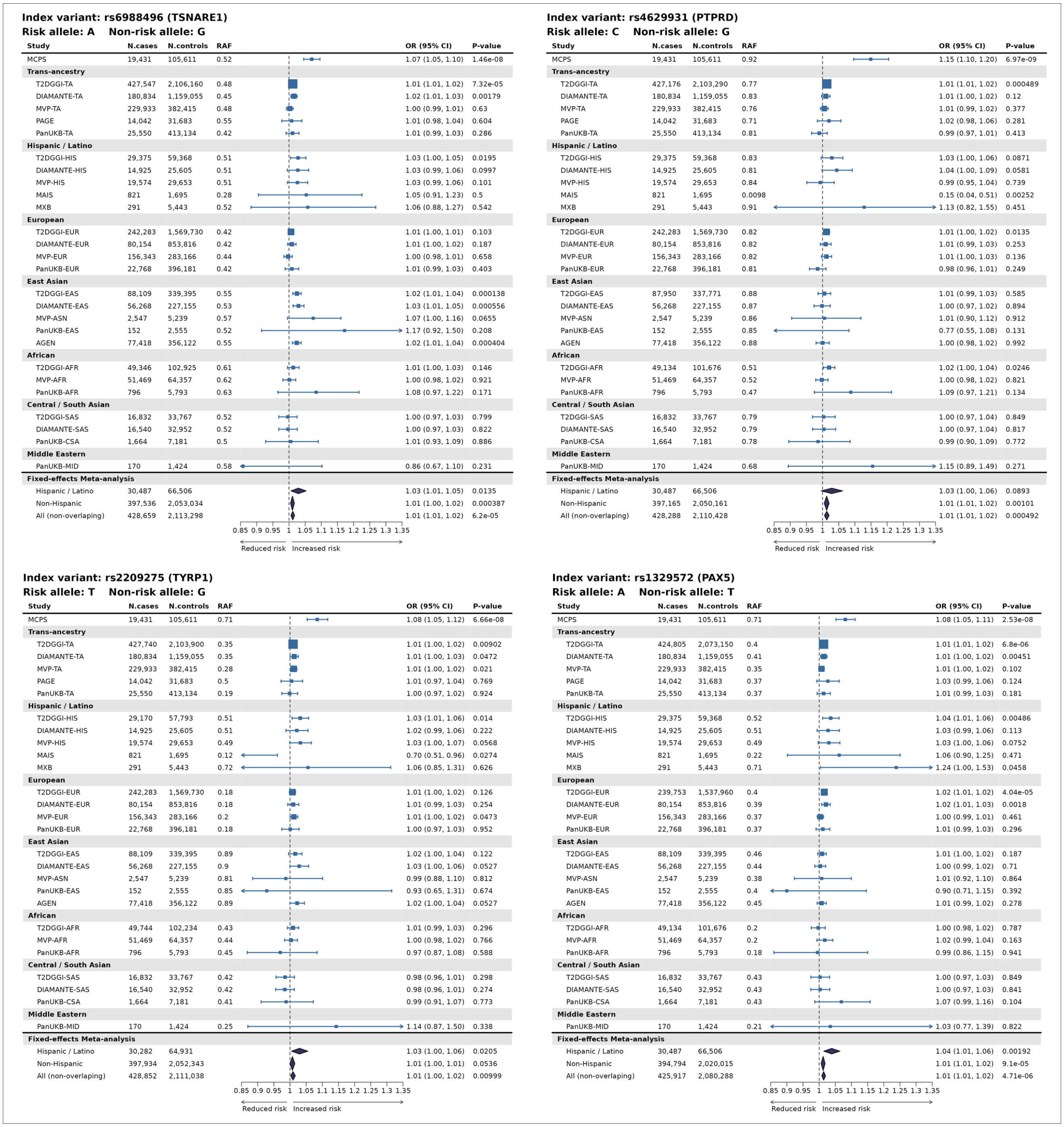

**Index variant: rs6988496 (TSNARE1)**
**Risk allele: A    Non-risk allele: G**

| Study | N.cases | N.controls | RAF | OR (95% CI) | P-value |
|---|---|---|---|---|---|
| MCPS | 19,431 | 105,611 | 0.52 | 1.07 (1.05, 1.10) | 1.46e-08 |
| **Trans-ancestry** | | | | | |
| T2DGGI-TA | 427,547 | 2,106,160 | 0.48 | 1.01 (1.01, 1.02) | 7.32e-05 |
| DIAMANTE-TA | 180,834 | 1,159,055 | 0.45 | 1.02 (1.01, 1.03) | 0.00179 |
| MVP-TA | 229,933 | 382,415 | 0.48 | 1.00 (0.99, 1.01) | 0.63 |
| PAGE | 14,042 | 31,683 | 0.55 | 1.01 (0.98, 1.04) | 0.604 |
| PanUKB-TA | 25,550 | 413,134 | 0.42 | 1.01 (0.99, 1.03) | 0.286 |
| **Hispanic / Latino** | | | | | |
| T2DGGI-HIS | 29,375 | 59,368 | 0.51 | 1.03 (1.00, 1.05) | 0.0195 |
| DIAMANTE-HIS | 14,925 | 25,605 | 0.51 | 1.03 (0.99, 1.06) | 0.0997 |
| MVP-HIS | 19,574 | 29,653 | 0.51 | 1.03 (0.99, 1.06) | 0.101 |
| MAIS | 821 | 1,695 | 0.28 | 1.05 (0.91, 1.23) | 0.5 |
| MXB | 291 | 5,443 | 0.52 | 1.06 (0.88, 1.27) | 0.542 |
| **European** | | | | | |
| T2DGGI-EUR | 242,283 | 1,569,730 | 0.42 | 1.01 (1.00, 1.01) | 0.103 |
| DIAMANTE-EUR | 80,154 | 853,816 | 0.42 | 1.01 (1.00, 1.02) | 0.187 |
| MVP-EUR | 156,343 | 283,166 | 0.44 | 1.00 (0.98, 1.01) | 0.658 |
| PanUKB-EUR | 22,768 | 396,181 | 0.42 | 1.01 (0.99, 1.03) | 0.403 |
| **East Asian** | | | | | |
| T2DGGI-EAS | 88,109 | 339,395 | 0.55 | 1.02 (1.01, 1.04) | 0.000138 |
| DIAMANTE-EAS | 56,268 | 227,155 | 0.53 | 1.03 (1.01, 1.05) | 0.000556 |
| MVP-ASN | 2,547 | 5,239 | 0.57 | 1.07 (1.00, 1.16) | 0.0655 |
| PanUKB-EAS | 152 | 2,555 | 0.52 | 1.17 (0.92, 1.50) | 0.208 |
| AGEN | 77,418 | 356,122 | 0.55 | 1.02 (1.01, 1.04) | 0.000404 |
| **African** | | | | | |
| T2DGGI-AFR | 49,346 | 102,925 | 0.61 | 1.01 (1.00, 1.03) | 0.146 |
| MVP-AFR | 51,469 | 64,357 | 0.62 | 1.00 (0.98, 1.02) | 0.921 |
| PanUKB-AFR | 796 | 5,793 | 0.63 | 1.08 (0.97, 1.22) | 0.171 |
| **Central / South Asian** | | | | | |
| T2DGGI-SAS | 16,832 | 33,767 | 0.52 | 1.00 (0.97, 1.03) | 0.799 |
| DIAMANTE-SAS | 16,540 | 32,952 | 0.52 | 1.00 (0.97, 1.03) | 0.822 |
| PanUKB-CSA | 1,664 | 7,181 | 0.5 | 1.01 (0.93, 1.09) | 0.886 |
| **Middle Eastern** | | | | | |
| PanUKB-MID | 170 | 1,424 | 0.58 | 0.86 (0.67, 1.10) | 0.231 |
| **Fixed-effects Meta-analysis** | | | | | |
| Hispanic / Latino | 30,487 | 66,506 | | 1.03 (1.01, 1.05) | 0.0135 |
| Non-Hispanic | 397,536 | 2,053,034 | | 1.01 (1.00, 1.02) | 0.000387 |
| All (non-overlaping) | 428,659 | 2,113,298 | | 1.01 (1.01, 1.02) | 6.2e-05 |

**Index variant: rs4629931 (PTPRD)**
**Risk allele: C    Non-risk allele: G**

| Study | N.cases | N.controls | RAF | OR (95% CI) | P-value |
|---|---|---|---|---|---|
| MCPS | 19,431 | 105,611 | 0.92 | 1.15 (1.10, 1.20) | 6.97e-09 |
| **Trans-ancestry** | | | | | |
| T2DGGI-TA | 427,176 | 2,103,290 | 0.77 | 1.01 (1.01, 1.02) | 0.000489 |
| DIAMANTE-TA | 180,834 | 1,159,055 | 0.83 | 1.01 (1.00, 1.02) | 0.12 |
| MVP-TA | 229,933 | 382,415 | 0.76 | 1.01 (0.99, 1.02) | 0.377 |
| PAGE | 14,042 | 31,683 | 0.71 | 1.02 (0.98, 1.06) | 0.281 |
| PanUKB-TA | 25,550 | 413,134 | 0.81 | 0.99 (0.97, 1.01) | 0.413 |
| **Hispanic / Latino** | | | | | |
| T2DGGI-HIS | 29,375 | 59,368 | 0.83 | 1.03 (1.00, 1.06) | 0.0871 |
| DIAMANTE-HIS | 14,925 | 25,605 | 0.81 | 1.04 (1.00, 1.09) | 0.0581 |
| MVP-HIS | 19,574 | 29,653 | 0.84 | 0.99 (0.95, 1.04) | 0.739 |
| MAIS | 821 | 1,695 | 0.0098 | 0.15 (0.04, 0.51) | 0.00252 |
| MXB | 291 | 5,443 | 0.91 | 1.13 (0.82, 1.55) | 0.451 |
| **European** | | | | | |
| T2DGGI-EUR | 242,283 | 1,569,730 | 0.82 | 1.01 (1.00, 1.02) | 0.0135 |
| DIAMANTE-EUR | 80,154 | 853,816 | 0.82 | 1.01 (0.99, 1.03) | 0.253 |
| MVP-EUR | 156,343 | 283,166 | 0.82 | 1.01 (1.00, 1.03) | 0.136 |
| PanUKB-EUR | 22,768 | 396,181 | 0.81 | 0.98 (0.96, 1.01) | 0.249 |
| **East Asian** | | | | | |
| T2DGGI-EAS | 87,950 | 337,771 | 0.88 | 1.01 (0.99, 1.03) | 0.585 |
| DIAMANTE-EAS | 56,268 | 227,155 | 0.87 | 1.00 (0.97, 1.02) | 0.894 |
| MVP-ASN | 2,547 | 5,239 | 0.86 | 1.01 (0.90, 1.12) | 0.912 |
| PanUKB-EAS | 152 | 2,555 | 0.85 | 0.77 (0.55, 1.08) | 0.131 |
| AGEN | 77,418 | 356,122 | 0.88 | 1.00 (0.98, 1.02) | 0.992 |
| **African** | | | | | |
| T2DGGI-AFR | 49,134 | 101,676 | 0.51 | 1.02 (1.00, 1.04) | 0.0246 |
| MVP-AFR | 51,469 | 64,357 | 0.52 | 1.00 (0.98, 1.02) | 0.821 |
| PanUKB-AFR | 796 | 5,793 | 0.47 | 1.09 (0.97, 1.21) | 0.134 |
| **Central / South Asian** | | | | | |
| T2DGGI-SAS | 16,832 | 33,767 | 0.79 | 1.00 (0.97, 1.04) | 0.849 |
| DIAMANTE-SAS | 16,540 | 32,952 | 0.79 | 1.00 (0.97, 1.04) | 0.817 |
| PanUKB-CSA | 1,664 | 7,181 | 0.78 | 0.99 (0.90, 1.09) | 0.772 |
| **Middle Eastern** | | | | | |
| PanUKB-MID | 170 | 1,424 | 0.68 | 1.15 (0.89, 1.49) | 0.271 |
| **Fixed-effects Meta-analysis** | | | | | |
| Hispanic / Latino | 30,487 | 66,506 | | 1.03 (1.00, 1.06) | 0.0893 |
| Non-Hispanic | 397,165 | 2,050,161 | | 1.01 (1.01, 1.02) | 0.00101 |
| All (non-overlaping) | 428,288 | 2,110,428 | | 1.01 (1.01, 1.02) | 0.000492 |

**Index variant: rs2209275 (TYRP1)**
**Risk allele: T    Non-risk allele: G**

| Study | N.cases | N.controls | RAF | OR (95% CI) | P-value |
|---|---|---|---|---|---|
| MCPS | 19,431 | 105,611 | 0.71 | 1.08 (1.05, 1.12) | 6.66e-08 |
| **Trans-ancestry** | | | | | |
| T2DGGI-TA | 427,740 | 2,103,900 | 0.35 | 1.01 (1.00, 1.02) | 0.00902 |
| DIAMANTE-TA | 180,834 | 1,159,055 | 0.35 | 1.01 (1.00, 1.03) | 0.0472 |
| MVP-TA | 229,933 | 382,415 | 0.28 | 1.01 (1.00, 1.02) | 0.021 |
| PAGE | 14,042 | 31,683 | 0.5 | 1.01 (0.97, 1.04) | 0.769 |
| PanUKB-TA | 25,550 | 413,134 | 0.19 | 1.00 (0.97, 1.02) | 0.924 |
| **Hispanic / Latino** | | | | | |
| T2DGGI-HIS | 29,170 | 57,793 | 0.51 | 1.03 (1.01, 1.06) | 0.014 |
| DIAMANTE-HIS | 14,925 | 25,605 | 0.51 | 1.02 (0.99, 1.06) | 0.222 |
| MVP-HIS | 19,574 | 29,653 | 0.49 | 1.03 (1.00, 1.07) | 0.0568 |
| MAIS | 821 | 1,695 | 0.12 | 0.70 (0.51, 0.96) | 0.0274 |
| MXB | 291 | 5,443 | 0.72 | 1.06 (0.85, 1.31) | 0.626 |
| **European** | | | | | |
| T2DGGI-EUR | 242,283 | 1,569,730 | 0.18 | 1.01 (1.00, 1.02) | 0.126 |
| DIAMANTE-EUR | 80,154 | 853,816 | 0.18 | 1.01 (0.99, 1.03) | 0.254 |
| MVP-EUR | 156,343 | 283,166 | 0.2 | 1.01 (1.00, 1.02) | 0.0473 |
| PanUKB-EUR | 22,768 | 396,181 | 0.18 | 1.00 (0.97, 1.03) | 0.952 |
| **East Asian** | | | | | |
| T2DGGI-EAS | 88,109 | 339,395 | 0.89 | 1.02 (1.00, 1.04) | 0.122 |
| DIAMANTE-EAS | 56,268 | 227,155 | 0.9 | 1.03 (1.00, 1.06) | 0.0527 |
| MVP-ASN | 2,547 | 5,239 | 0.81 | 0.99 (0.88, 1.10) | 0.812 |
| PanUKB-EAS | 152 | 2,555 | 0.85 | 0.93 (0.65, 1.31) | 0.674 |
| AGEN | 77,418 | 356,122 | 0.89 | 1.02 (1.00, 1.04) | 0.0527 |
| **African** | | | | | |
| T2DGGI-AFR | 49,744 | 102,234 | 0.43 | 1.01 (0.99, 1.03) | 0.296 |
| MVP-AFR | 51,469 | 64,357 | 0.44 | 1.00 (0.98, 1.02) | 0.766 |
| PanUKB-AFR | 796 | 5,793 | 0.45 | 0.97 (0.87, 1.08) | 0.588 |
| **Central / South Asian** | | | | | |
| T2DGGI-SAS | 16,832 | 33,767 | 0.42 | 0.98 (0.96, 1.01) | 0.298 |
| DIAMANTE-SAS | 16,540 | 32,952 | 0.42 | 0.98 (0.96, 1.01) | 0.274 |
| PanUKB-CSA | 1,664 | 7,181 | 0.41 | 0.99 (0.91, 1.07) | 0.773 |
| **Middle Eastern** | | | | | |
| PanUKB-MID | 170 | 1,424 | 0.25 | 1.14 (0.87, 1.50) | 0.338 |
| **Fixed-effects Meta-analysis** | | | | | |
| Hispanic / Latino | 30,282 | 64,931 | | 1.03 (1.00, 1.06) | 0.0205 |
| Non-Hispanic | 397,934 | 2,052,343 | | 1.01 (1.00, 1.01) | 0.0536 |
| All (non-overlaping) | 428,852 | 2,111,038 | | 1.01 (1.00, 1.02) | 0.00999 |

**Index variant: rs1329572 (PAX5)**
**Risk allele: A    Non-risk allele: T**

| Study | N.cases | N.controls | RAF | OR (95% CI) | P-value |
|---|---|---|---|---|---|
| MCPS | 19,431 | 105,611 | 0.71 | 1.08 (1.05, 1.11) | 2.53e-08 |
| **Trans-ancestry** | | | | | |
| T2DGGI-TA | 424,805 | 2,073,150 | 0.4 | 1.01 (1.01, 1.02) | 6.8e-06 |
| DIAMANTE-TA | 180,834 | 1,159,055 | 0.41 | 1.01 (1.00, 1.02) | 0.00451 |
| MVP-TA | 229,933 | 382,415 | 0.35 | 1.01 (1.00, 1.02) | 0.102 |
| PAGE | 14,042 | 31,683 | 0.37 | 1.03 (0.99, 1.06) | 0.124 |
| PanUKB-TA | 25,550 | 413,134 | 0.37 | 1.01 (0.99, 1.03) | 0.181 |
| **Hispanic / Latino** | | | | | |
| T2DGGI-HIS | 29,375 | 59,368 | 0.52 | 1.04 (1.01, 1.06) | 0.00486 |
| DIAMANTE-HIS | 14,925 | 25,605 | 0.51 | 1.03 (0.99, 1.06) | 0.113 |
| MVP-HIS | 19,574 | 29,653 | 0.49 | 1.03 (1.00, 1.06) | 0.0752 |
| MAIS | 821 | 1,695 | 0.22 | 1.06 (0.90, 1.25) | 0.471 |
| MXB | 291 | 5,443 | 0.71 | 1.24 (1.00, 1.53) | 0.0458 |
| **European** | | | | | |
| T2DGGI-EUR | 239,753 | 1,537,960 | 0.4 | 1.02 (1.01, 1.02) | 4.04e-05 |
| DIAMANTE-EUR | 80,154 | 853,816 | 0.39 | 1.02 (1.01, 1.03) | 0.0018 |
| MVP-EUR | 156,343 | 283,166 | 0.37 | 1.00 (0.99, 1.01) | 0.461 |
| PanUKB-EUR | 22,768 | 396,181 | 0.37 | 1.01 (0.99, 1.03) | 0.296 |
| **East Asian** | | | | | |
| T2DGGI-EAS | 88,109 | 339,395 | 0.46 | 1.01 (1.00, 1.02) | 0.187 |
| DIAMANTE-EAS | 56,268 | 227,155 | 0.44 | 1.00 (0.99, 1.02) | 0.71 |
| MVP-ASN | 2,547 | 5,239 | 0.38 | 1.01 (0.92, 1.10) | 0.864 |
| PanUKB-EAS | 152 | 2,555 | 0.4 | 0.90 (0.71, 1.15) | 0.392 |
| AGEN | 77,418 | 356,122 | 0.45 | 1.01 (0.99, 1.02) | 0.278 |
| **African** | | | | | |
| T2DGGI-AFR | 49,134 | 101,676 | 0.2 | 1.00 (0.98, 1.02) | 0.787 |
| MVP-AFR | 51,469 | 64,357 | 0.2 | 1.02 (0.99, 1.04) | 0.163 |
| PanUKB-AFR | 796 | 5,793 | 0.18 | 0.99 (0.86, 1.15) | 0.941 |
| **Central / South Asian** | | | | | |
| T2DGGI-SAS | 16,832 | 33,767 | 0.43 | 1.00 (0.97, 1.03) | 0.849 |
| DIAMANTE-SAS | 16,540 | 32,952 | 0.43 | 1.00 (0.97, 1.03) | 0.841 |
| PanUKB-CSA | 1,664 | 7,181 | 0.43 | 1.07 (0.99, 1.16) | 0.104 |
| **Middle Eastern** | | | | | |
| PanUKB-MID | 170 | 1,424 | 0.21 | 1.03 (0.77, 1.39) | 0.822 |
| **Fixed-effects Meta-analysis** | | | | | |
| Hispanic / Latino | 30,487 | 66,506 | | 1.04 (1.01, 1.06) | 0.00192 |
| Non-Hispanic | 394,794 | 2,020,015 | | 1.01 (1.01, 1.02) | 9.1e-05 |
| All (non-overlaping) | 425,917 | 2,080,288 | | 1.01 (1.01, 1.02) | 4.71e-06 |



#### Supplementary Fig. 3 (continued).

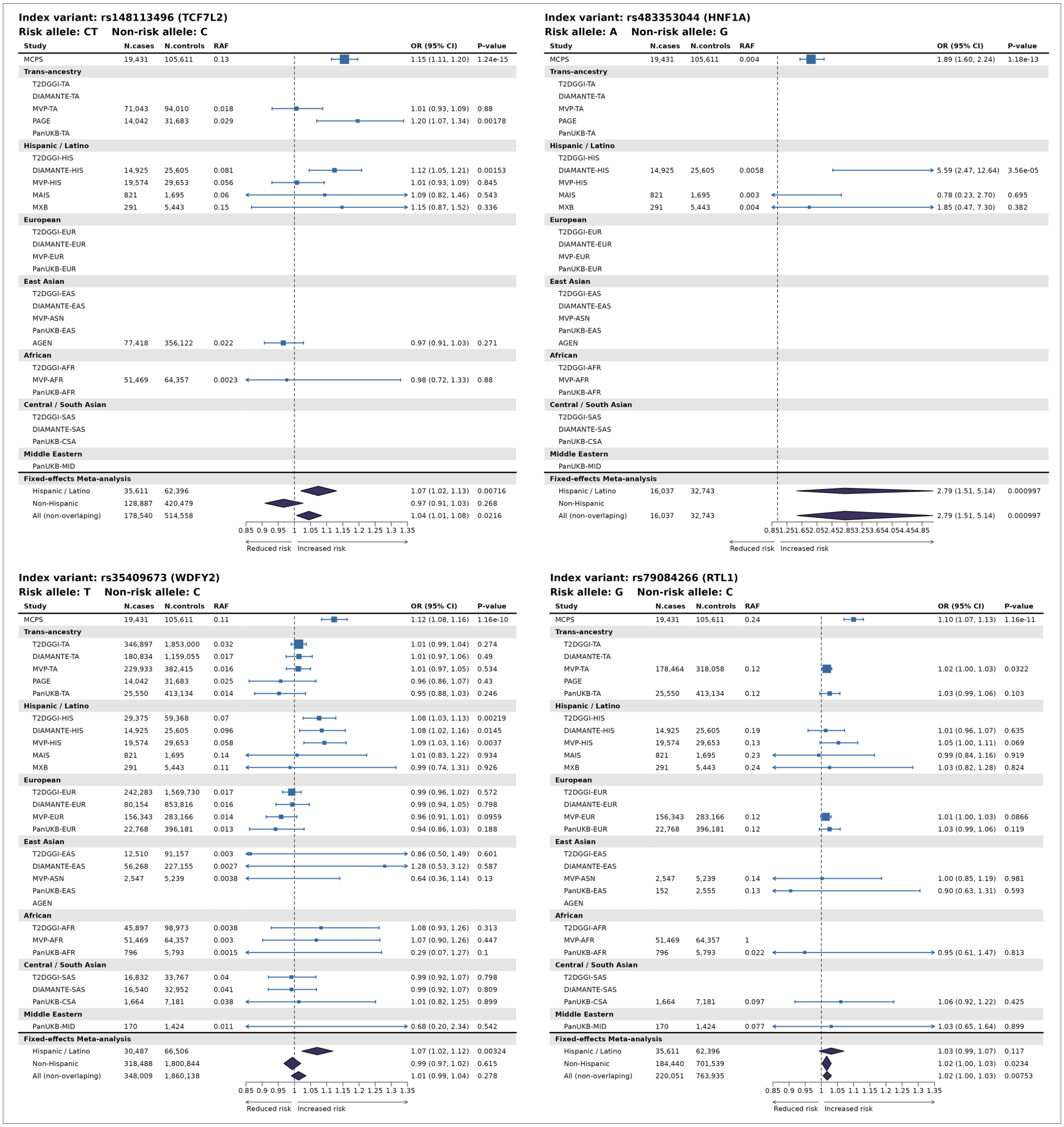


**Supplementary Fig. 3 (continued).**

**Index variant: rs79783591 (MC4R)**
**Risk allele: T    Non-risk allele: A**

| Study | N.cases | N.controls | RAF | OR (95% CI) | P-value |
|---|---|---|---|---|---|
| MCPS | 19,431 | 105,611 | 0.009 | 1.45 (1.30, 1.63) | 4.74e-11 |
| **Trans-ancestry** | | | | | |
| T2DGGI-TA | | | | | |
| DIAMANTE-TA | | | | | |
| MVP-TA | | | | | |
| PAGE | | | | | |
| PanUKB-TA | | | | | |
| **Hispanic / Latino** | | | | | |
| T2DGGI-HIS | | | | | |
| DIAMANTE-HIS | 14,925 | 25,605 | 0.005 | 1.47 (0.98, 2.20) | 0.0606 |
| MVP-HIS | 19,574 | 29,653 | 0.003 | 1.26 (0.96, 1.66) | 0.097 |
| MAIS | 821 | 1,695 | 0.0022 | 2.76 (0.67, 11.35) | 0.158 |
| MXB | 291 | 5,443 | 0.0071 | 1.03 (0.29, 3.59) | 0.964 |
| **European** | | | | | |
| T2DGGI-EUR | | | | | |
| DIAMANTE-EUR | | | | | |
| MVP-EUR | | | | | |
| PanUKB-EUR | | | | | |
| **East Asian** | | | | | |
| T2DGGI-EAS | | | | | |
| DIAMANTE-EAS | | | | | |
| MVP-ASN | | | | | |
| PanUKB-EAS | | | | | |
| AGEN | | | | | |
| **African** | | | | | |
| T2DGGI-AFR | | | | | |
| MVP-AFR | | | | | |
| PanUKB-AFR | | | | | |
| **Central / South Asian** | | | | | |
| T2DGGI-SAS | | | | | |
| DIAMANTE-SAS | | | | | |
| PanUKB-CSA | | | | | |
| **Middle Eastern** | | | | | |
| PanUKB-MID | | | | | |
| **Fixed-effects Meta-analysis** | | | | | |
| Hispanic / Latino | 35,611 | 62,396 | | 1.34 (1.07, 1.67) | 0.00965 |
| Non-Hispanic | | | | | |
| All (non-overlaping) | 35,611 | 62,396 | | 1.34 (1.07, 1.67) | 0.00965 |

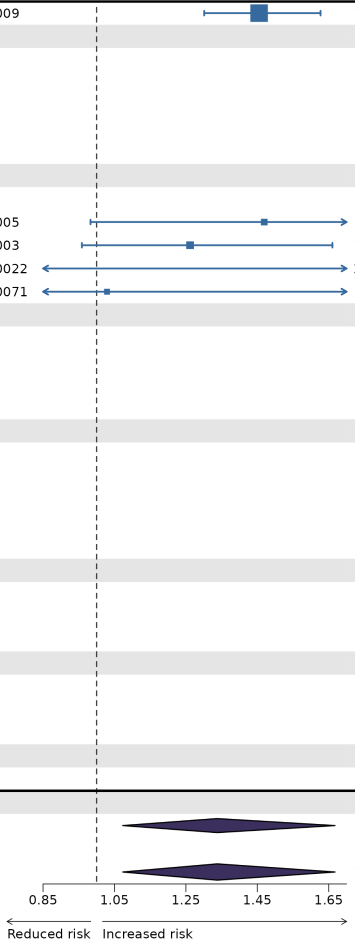


#### Supplementary Fig. 3 (continued).

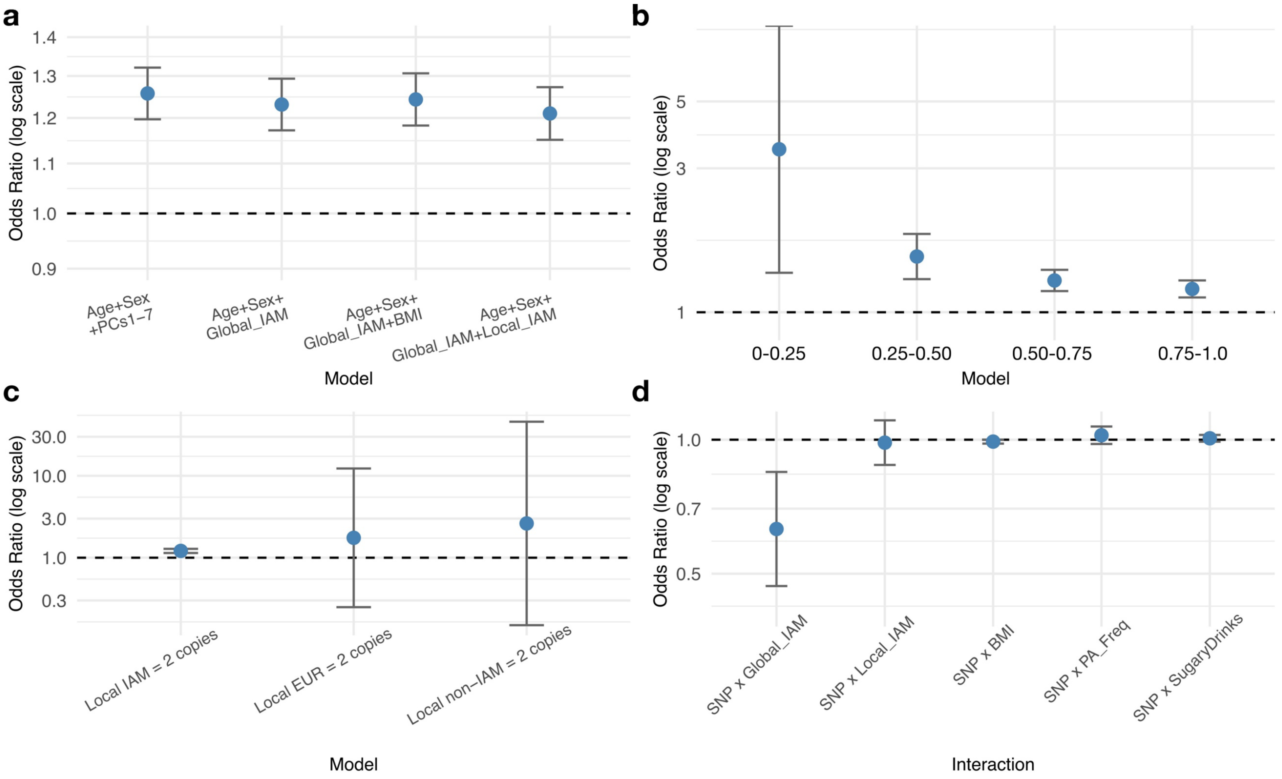


**Supplementary Fig. 4 | Sensitivity and interaction analyses for rs139343836 at the *RFX6* locus.**

**a**, Effect estimates from models with differing adjustment for age, sex, BMI, genetic principal components (PCs), global Indigenous American (IAM) ancestry proportion and local IAM ancestry. **b**, Effect estimates stratified by global IAM ancestry proportion, adjusted for age and sex. **c**, Effect estimates stratified by local ancestry, restricted to individuals carrying two copies of IAM, European or non-IAM ancestry at the locus. Models were adjusted for age, sex and global IAM ancestry proportion. **d**, Interaction analyses between rs139343836 and global IAM ancestry, local IAM ancestry, BMI, physical activity frequency (PA_Freq) and sugary drink consumption frequency. All models were adjusted for age, sex and global IAM ancestry proportion. Effect estimates in **a–c** were obtained using SUGEN, whereas interaction analyses in **d** were performed using REGENIE v3.

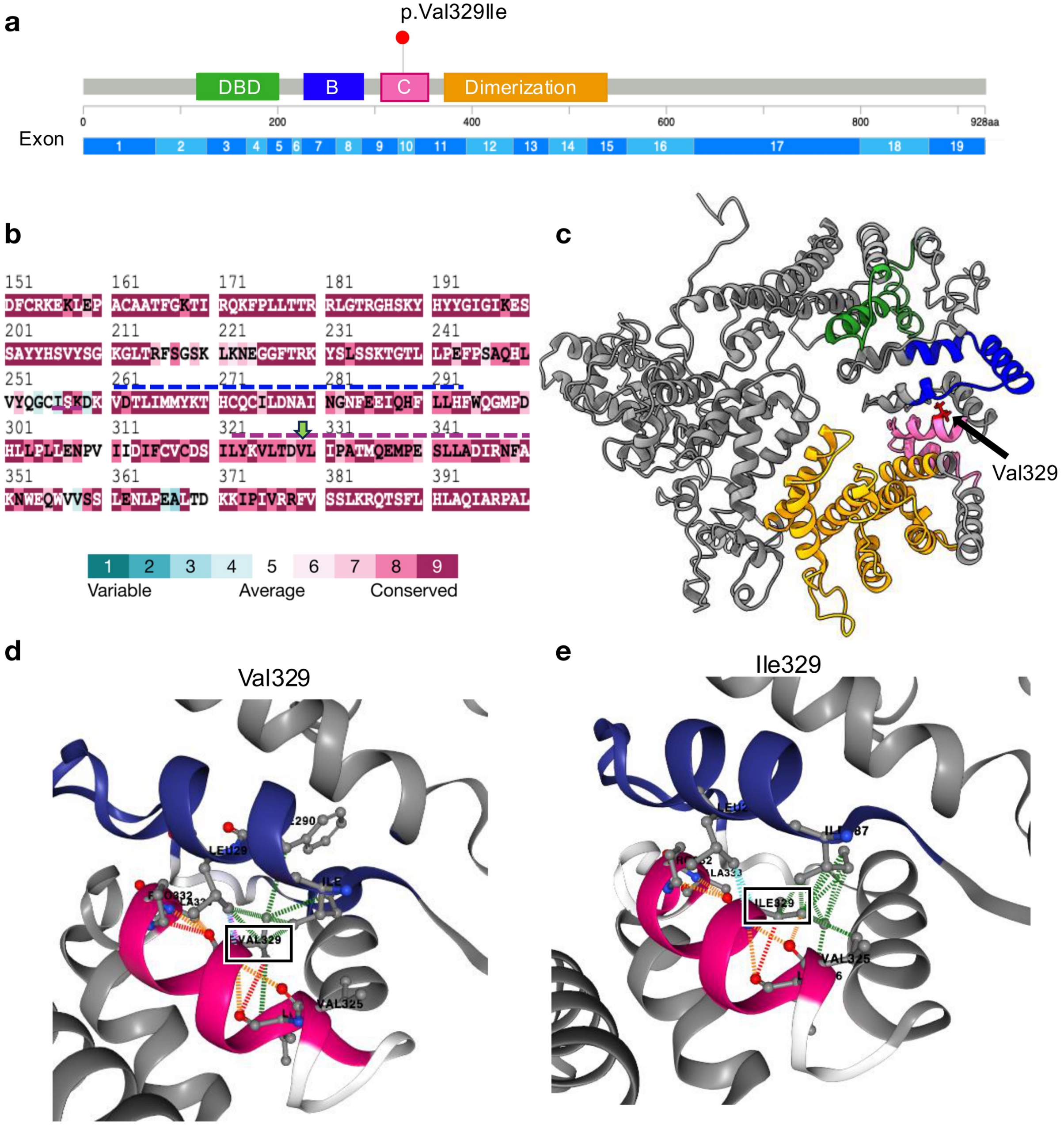


**Supplementary Fig. 5 | Protein modelling analysis of the p.Val329Ile substitution in *RFX6*.**
**a**, Domain structure of RFX6 showing the location of the p.Val329Ile variant within the C region of the protein. **b**, Amino acid conservation across RFX6. The green arrow indicates residue 329, and dotted blue and magenta lines denote the B and C regions, respectively. **c**, Predicted protein structure of RFX6 generated using I-TASSER. Colours correspond to the domain structure shown in **a**. **d**,**e**, Close-up views of the C region containing either valine (**d**) or isoleucine (**e**) at residue 329. Dotted lines indicate predicted intra-protein interactions, including hydrophobic (green), polar (orange), ionic (yellow), van der Waals (blue) and clash (pink) interactions.

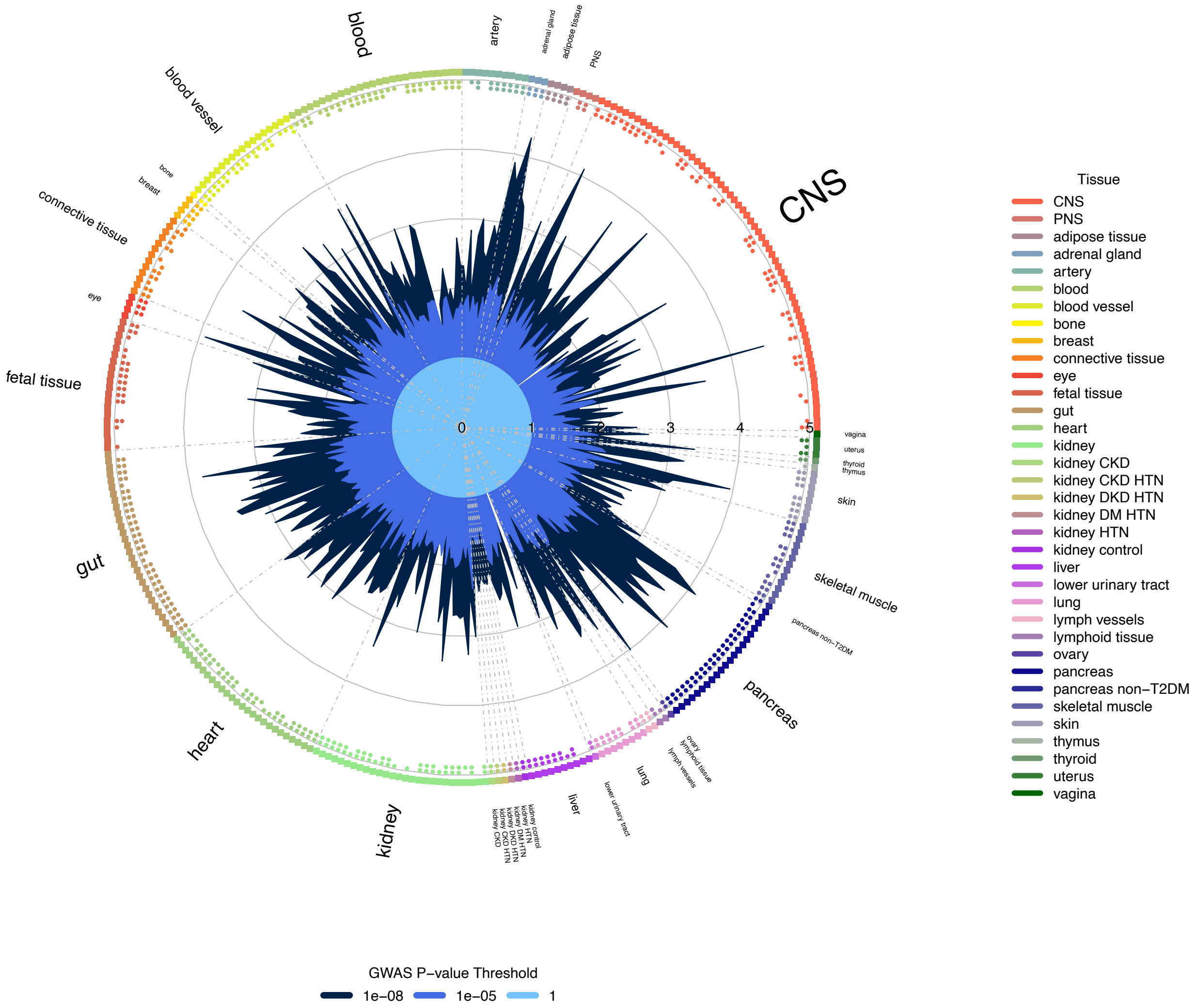


**Supplementary Fig. 6 | Genome-wide enrichment of chromatin accessibility annotations.** GARFIELD was used to test for enrichment of DNase I hypersensitive sites (DHS), ATAC-seq peaks and single-nucleus ATAC-seq (snATAC-seq) peaks among T2D-associated variants in MCPS. Annotations correspond to bulk tissue and single-cell chromatin accessibility maps obtained from the Common Metabolic Diseases Genome Atlas. Radial axis values indicate odds ratios from the GARFIELD logistic regression model, which quantify enrichment of trait-associated variants within each annotation.

Abbreviations: CNS, central nervous system; PNS, peripheral nervous system; CKD, chronic kidney disease; HTN, hypertension; DKD, diabetic kidney disease; DM, diabetes mellitus.

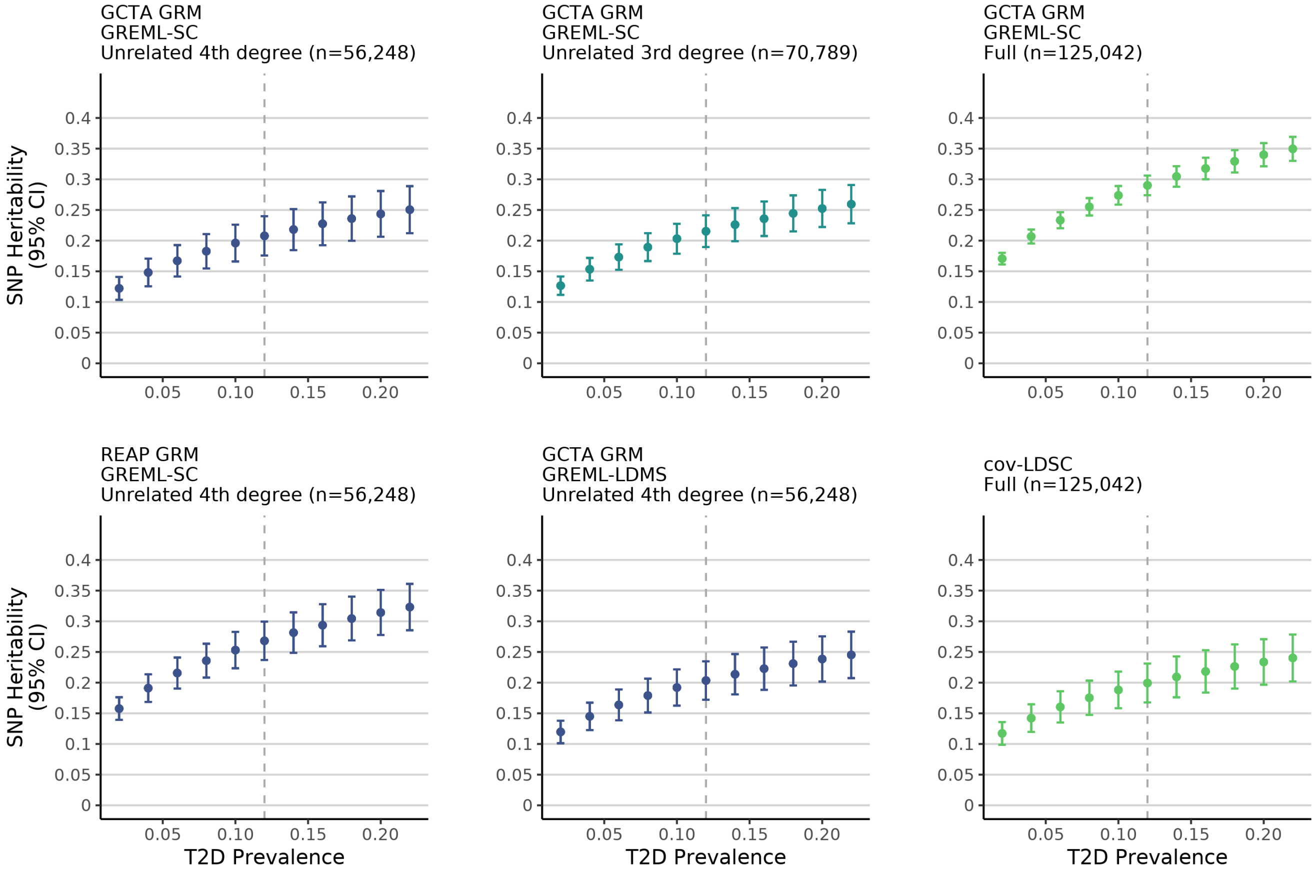


**Supplementary Fig. 7 | Sensitivity of liability-scale SNP heritability estimates to disease prevalence.** Liability-scale SNP heritability ($h^2_{SNP}$) estimates are shown across a range of assumed T2D prevalence values for the four heritability estimation methods evaluated in MCPS. The vertical dashed line indicates the prevalence estimate used in the primary analyses ($K$ = 0.12). Error bars indicate 95% confidence intervals where applicable.

Abbreviations: cov-LDSC, covariate-adjusted LD score regression; GREML-SC, single-component GREML; GREML-LDMS, LD- and MAF-stratified GREML; REAP, relatedness estimation in admixed populations.

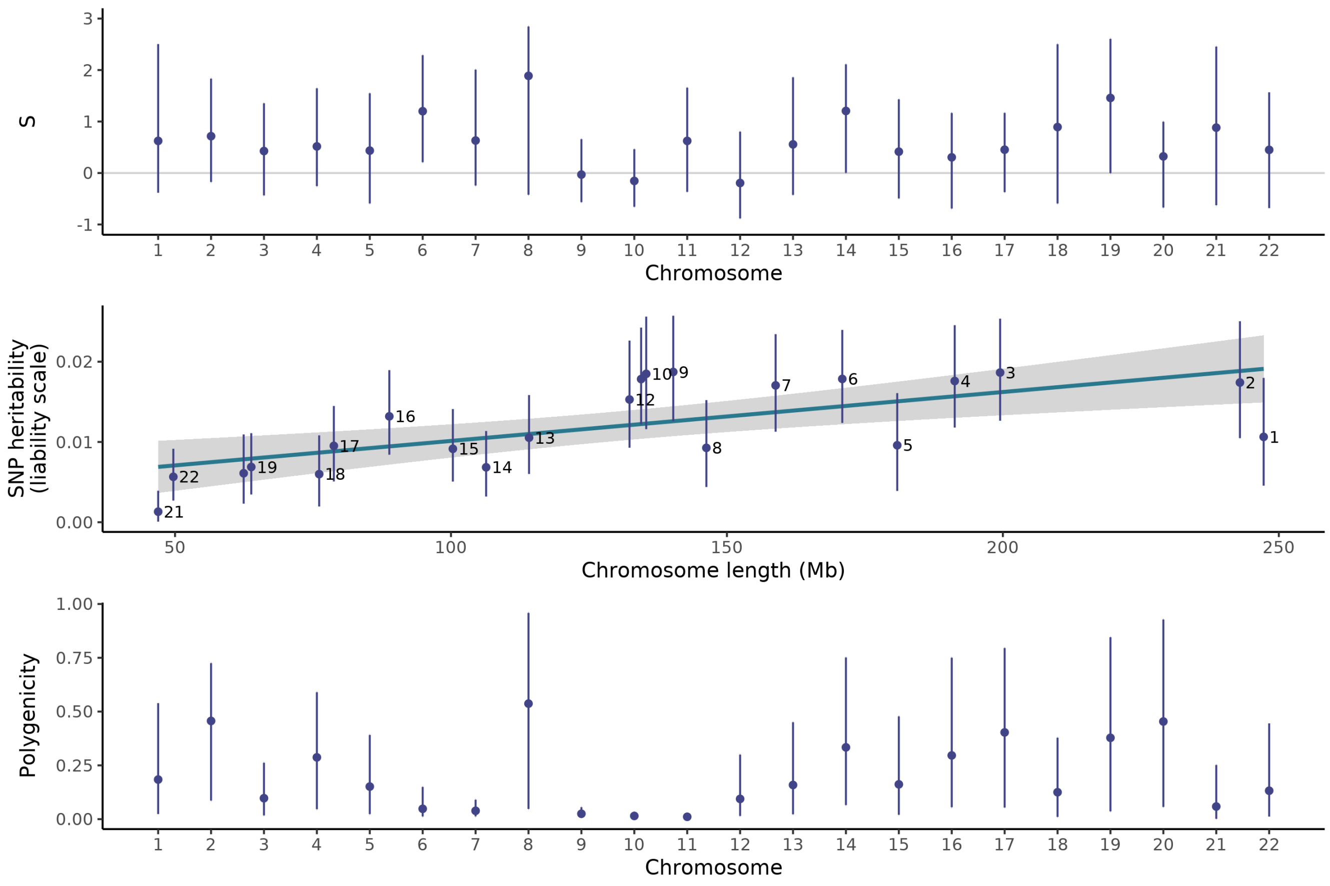


**Supplementary Fig. 8 | Per-chromosome genetic architecture parameters for T2D from GCTB-BayesS.**

Points and bars represent posterior parameters with 95% credible intervals. The sample size is 56,248 participants maximally unrelated to the fourth degree. Analyses used 333,195 genotyped variants after removing those with MAF <1%, Hardy-Weinberg equilibrium test $p < 10^{-6}$, and genotype missing rate >5% and were adjusted for age, sex, and the first seven genetic PCs. Per-chromosome $h^2_{SNP}$ scaled linearly with chromosome length.

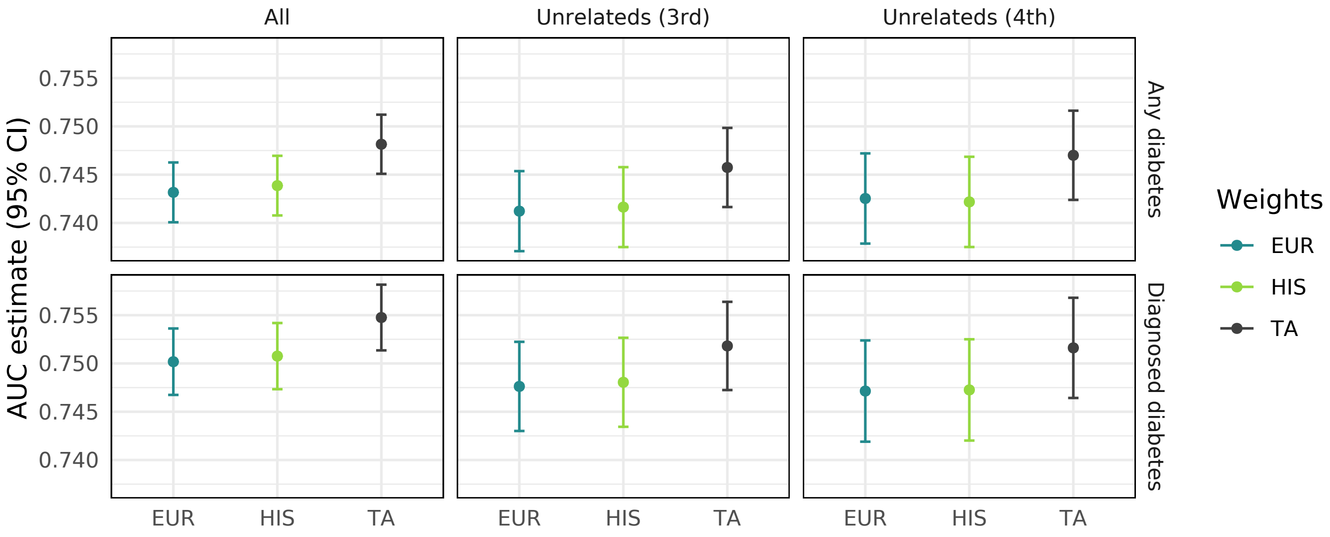


**Supplementary Fig. 9 | Predictive performance of T2D polygenic risk scores in MCPS.** Polygenic risk scores (PRSs) were calculated using trans-ancestry (TA), European (EUR) and Hispanic (HIS) effect estimates from the T2DGGI study (Suzuki et al. 2025). Predictive performance was evaluated separately among all participants and subsets of participants with increasing levels of relatedness. Area under the receiver operating characteristic curve (AUC) estimates were obtained from logistic regression models adjusted for age, sex, district of residence and body mass index, with each PRS modelled as a continuous variable (per one standard deviation increase). The designation of 'diagnosed diabetes' is consistent with the type 2 diabetes case definition used in the GWAS, whereas 'any diabetes' additionally includes participants with undiagnosed type 2 diabetes, defined by HbA1c levels ≥ 6.5%.

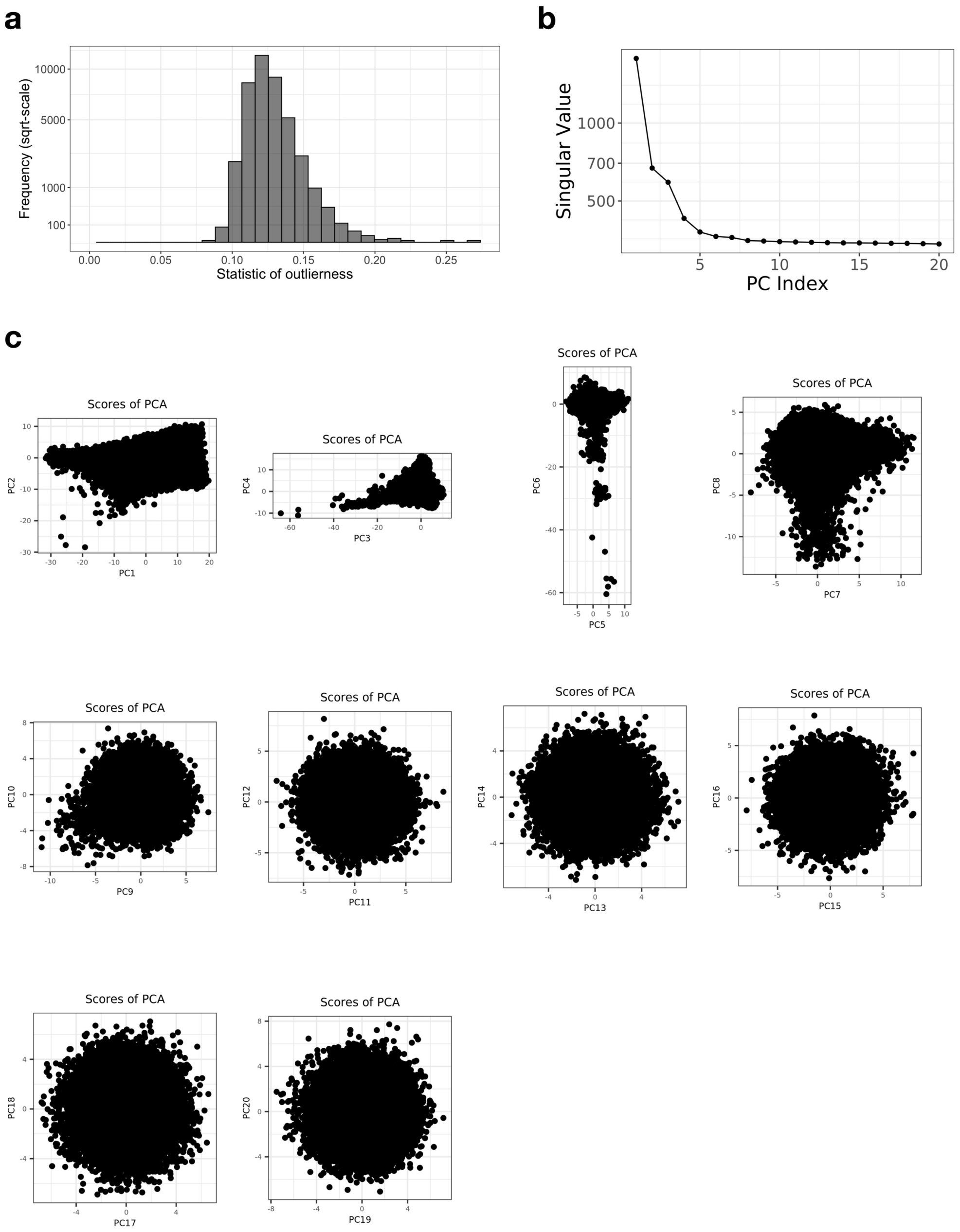


**Supplementary Fig. 10 | Principal component analysis of the MCPS cohort.**
PCA was performed using 539,448 autosomal variants genotyped in 140,829 MCPS participants. **a**, Distribution of the per-individual outlier statistic used to identify sample outliers. **b**, Scree plot of principal component singular values. **c**, Pairwise plots of principal component scores. PCA was performed using the workflow of Privé et al.

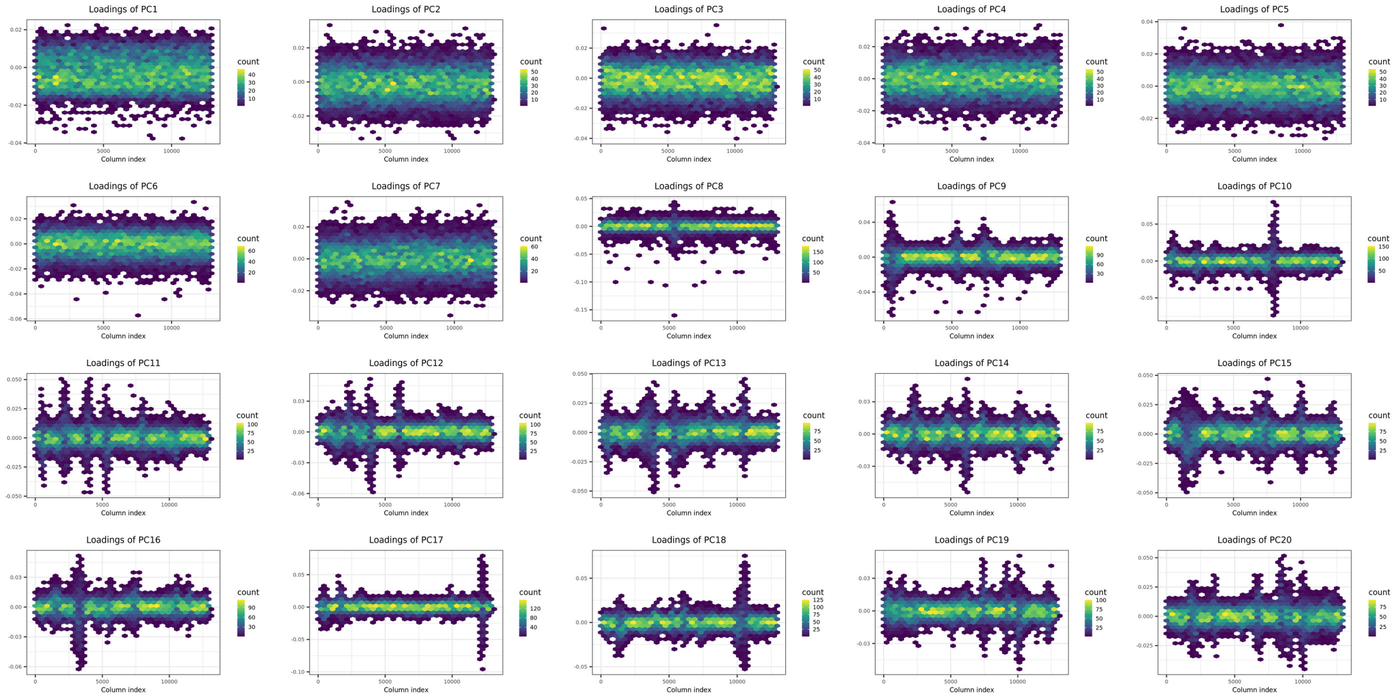


**Supplementary Fig. 11 | Distribution of principal component loadings in MCPS.**
Principal component loadings are shown across the genome for the first 20 principal components from the PCA of 140,829 MCPS participants. Colours indicate the density of loadings along the genome. Principal components exhibiting approximately normal loading distributions are consistent with population structure, whereas components containing localized peaks are suggestive of local LD structure.

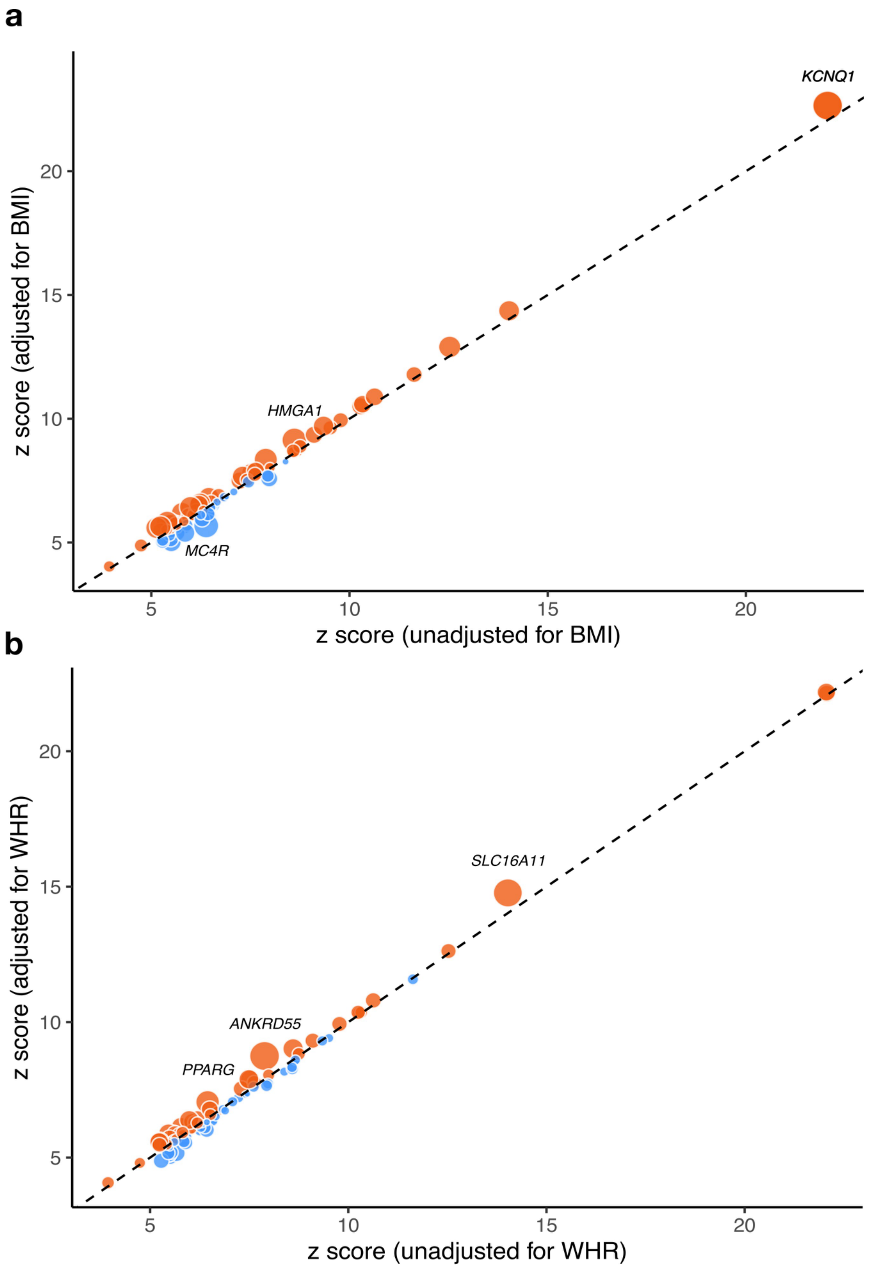


**Supplementary Fig. 12 | Impact of adiposity adjustment on GWAS signals.**
*Z* scores are shown for GWAS index variants with and without adjustment for **a**, BMI or **b**, waist-to-hip ratio (WHR). Red indicates signals for which adiposity adjustment increased the strength of association, whereas blue indicates signals for which adjustment attenuated the association. Point size corresponds to –log10(*p*) for the test of difference in effect size between adjusted and unadjusted models ($P_{diff}$). Gene names are shown for signals with FDR-adjusted $P_{diff} < 0.05$.

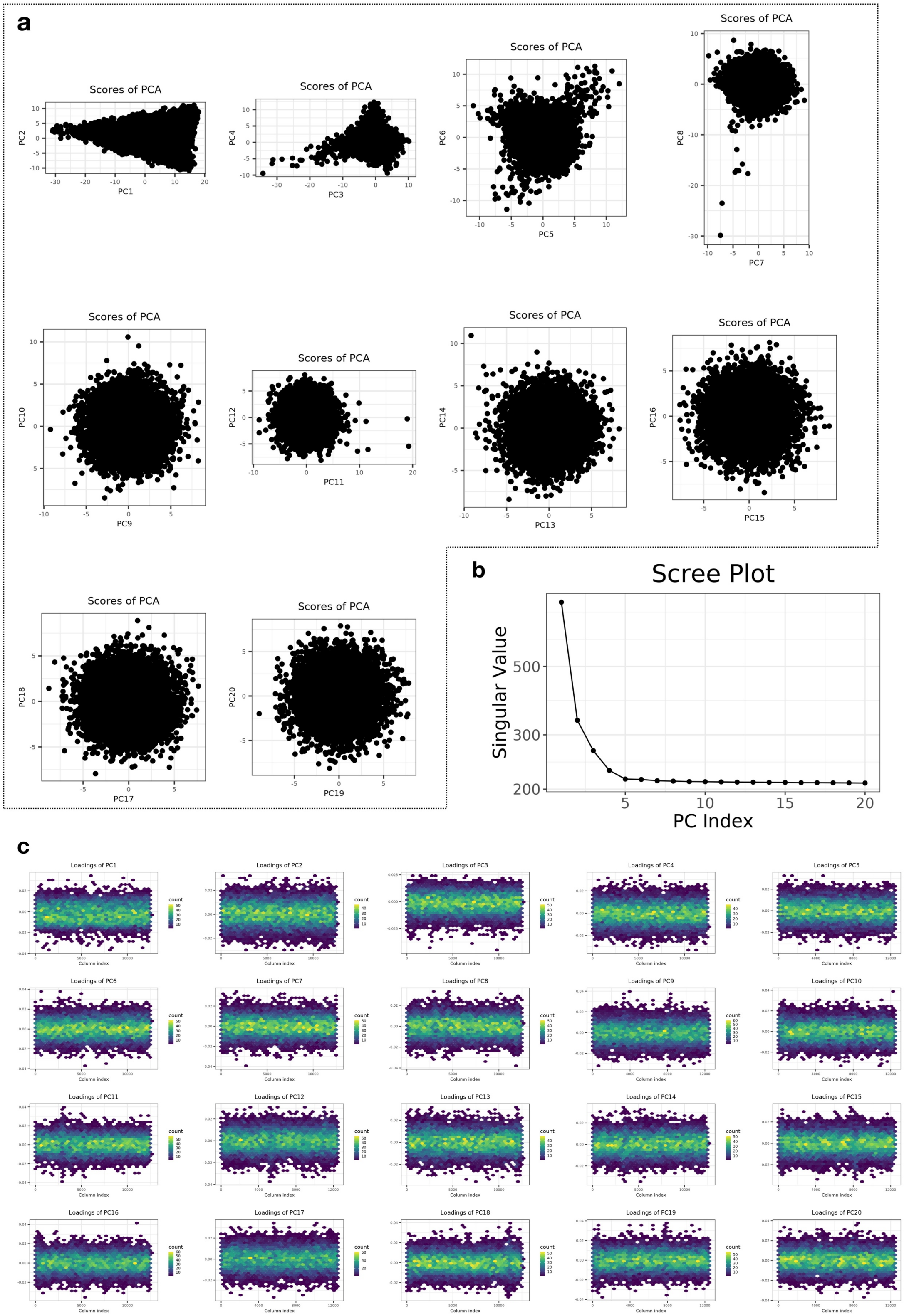


**Supplementary Fig. 13 | Principal component analysis of 9,764 whole-genome sequenced MCPS participants.**

PCA was performed using the workflow of Privé et al. with variants filtered at MAF ≥ 0.05 and LD-pruned at $r^2$ = 0.005. **a**, Pairwise plots of principal component scores. **b**, Scree plot of principal component singular values. **c**, Genomic distribution of principal component loadings.
